# Safety and immunogenicity of a SARS-CoV-2 recombinant protein nanoparticle vaccine (GBP510) adjuvanted with AS03: a phase 1/2, randomized, placebo-controlled, observer-blinded trial

**DOI:** 10.1101/2022.03.30.22273143

**Authors:** Joon Young Song, Won Suk Choi, Jung Yeon Heo, Jin Soo Lee, Dong Sik Jung, Shin-Woo Kim, Kyung-Hwa Park, Joong Sik Eom, Su Jin Jeong, Jacob Lee, Ki Tae Kwon, Hee Jung Choi, Jang Wook Sohn, Young Keun Kim, Ji Yun Noh, Woo Joo Kim, François Roman, Maria Angeles Ceregido, Francesca Solmi, Agathe Philippot, Lauren Carter, David Veesler, Neil King, Hun Kim, Ji Hwa Ryu, Su Jeen Lee, Yong Wook Park, Ho Keun Park, Hee Jin Cheong

## Abstract

**Background:** Vaccination has helped to mitigate the COVID-19 pandemic. Ten traditional and novel vaccines have been listed by the World Health Organization for emergency use. Additional alternative approaches may better address ongoing vaccination globally, where there remains an inequity in vaccine distribution. GBP510 is a recombinant protein vaccine, which consists of self-assembling, two-component nanoparticles displaying the receptor-binding domain (RBD) in a highly immunogenic array.

**Methods:** We conducted a randomized, placebo-controlled, observer-blinded, phase 1/2 trial to evaluate the safety and immunogenicity of GBP510 (2-doses at a 28-day interval) adjuvanted with or without AS03 in adults aged 19–85 years. The main outcomes included solicited and unsolicited adverse events; anti-SARS-CoV-2 RBD IgG antibody and neutralizing antibody responses; T-cell immune responses.

**Findings:** Of 328 participants who underwent randomization, 327 participants received at least 1 dose of vaccine. Each received either 10 μg GBP510 adjuvanted with AS03 (n = 101), 10 μg unadjuvanted GBP510 (n = 10), 25 μg GBP510 adjuvanted with AS03 (n = 104), 25 μg unadjuvanted GBP510 (n = 51), or placebo (n = 61). Most solicited adverse events were mild-to-moderate in severity and transient. Higher reactogenicity was observed in the GBP510 adjuvanted with AS03 groups compared to the non-adjuvanted and placebo groups. Reactogenicity was higher post-dose 2 compared to post-dose 1, particularly for systemic adverse events. The geometric mean concentrations of anti-SARS-CoV-2-RBD IgG antibody reached 2163.6/2599.2 BAU/mL in GBP510 adjuvanted with AS03 recipients (10 μg/25 μg) by 14 days after the second dose. Two-dose vaccination with 10 μg or 25 μg GBP510 adjuvanted with AS03 induced high titers of neutralizing antibody via pseudovirus (1369.0/1431.5 IU/mL) and wild-type virus (949.8/861.0 IU/mL) assays.

**Interpretation:** GBP510 adjuvanted with AS03 was well tolerated and highly immunogenic. These results support further development of the vaccine candidate, which is currently being evaluated in Phase 3.

**Funding:** Coalition for Epidemic Preparedness Innovations

**RESEARCH IN CONTEXT:** *Evidence before this study:* We searched PubMed for research articles published by December 31, 2021, using the terms “COVID-19” or “SARS-CoV-2,” “vaccine,” and “clinical trial.” In previously reported randomized clinical trials, we found that mRNA vaccines were more immunogenic than adenovirus-vectored vaccines. Solicited adverse events were more frequent and more severe in intensity after the first dose compared to the second dose for adenovirus-vectored vaccines, whereas they increased after the second dose of mRNA or recombinant spike-protein nanoparticle vaccines.

*Added value of this study:* This is the first human study evaluating the immunogenicity and safety of recombinant SARS-CoV-2 protein nanoparticle with and without adjuvant AS03, designed to elicit functional cross-protective responses via receptor-binding domain (RBD). Both 10 and 25 μg of GBP510 with AS03 formulations were well tolerated with an acceptable safety profile. Potent humoral immune responses against the SARS-CoV-2 RBD were induced and peaked by day 42 (14 days after the second dose). In addition, GBP510 adjuvanted with AS03 elicited a noticeable Th1 response, with production of IFN-γ, TNF-α, and IL-2. IL-4 was inconsistent and IL-5 nearly inexistent response across all groups.

*Implications of the available evidence:* The results from this phase 1/2 trial indicate that GBP510 adjuvanted with AS03 has an acceptable safety profile with no vaccine-related serious adverse events. Two-dose immunization with GBP510 adjuvanted with AS03 induced potent humoral and cellular immune responses against SARS-CoV-2.

## INTRODUCTION

Since December 2019, more than 349 million individuals worldwide have been diagnosed with severe acute respiratory syndrome coronavirus 2 (SARS-CoV-2) infection, resulting in 5,592,266 deaths as of January 25, 2022.^1^ With global effort to control pandemics, several SARS-CoV-2 vaccines based on different technologies have been approved and play key roles to reduce this ongoing health crisis. While these vaccines demonstrated remarkable reduction in disease, there still remain urgent and unmet needs for the vaccine worldwide and development of new platform vaccines is needed to resolve cold chain supply issues, disparity of vaccination rate between countries, and emergence of variants.^2^ In response to these challenges, SK Bioscience Co., Ltd. (Korea) has been focusing on developing synthetic protein-based vaccine candidates especially targeting RBD region as an antigen which has potential advantages on high production yield, thermostability, and focusing immunity on key protective determinants, anticipating that would help increase population immunity.^2^

GBP510 is a recombinant protein vaccine that consists of self-assembling, two-component nanoparticles displaying the receptor-binding domain (RBD) of the SARS-CoV-2 Spike protein in a highly immunogenic array. Unlike traditional protein subunit vaccines, which are easily degraded by various proteolytic enzymes in the blood, GBP510 is designed to display SARS-CoV-2 RBD on nanoparticle scaffolds, hence it could induce strong antibody responses with delayed degradation.^3,4^ In addition, GBP510 can provide protective immunity against diverse escape mutants by inducing antibodies targeting multiple, conserved, non-overlapping RBD epitopes.^3,4^ Thus, in a mouse model, vaccination with RBD nanoparticles elicited 10-fold higher neutralizing antibody titers compared to the prefusion-stabilized spike ectodomain trimer, even at a 5-fold lower dose, and protected against mouse-adapted SARS-CoV-2 challenge.^3^ Moreover, a multivalent RBD nanoparticle vaccine elicited diverse polyclonal antibody responses, thereby providing protection against heterotypic SARS-CoV-2.^4^

Here, we report the safety and immunogenicity data until 4 weeks after the second dose from a randomized, placebo-controlled, observer-blinded, phase 1/2 trial of GBP510 adjuvanted with or without AS03 in healthy younger and older adults.

## METHODS

### Study design and participants

This phase 1/2, randomized, placebo-controlled, observer-blinded, dose-escalating trial was conducted at 14 centers in South Korea. Healthy adults aged 19–85 years were eligible for the study. Participants were excluded from the study if they met any of the following criteria: any clinically significant respiratory symptoms, febrile illness, or acute illness within 72 hours; previous diagnosis of laboratory-confirmed COVID-19; previous diagnosis of laboratory-confirmed SARS or middle east respiratory syndrome (MERS); exposure to person with SARS-CoV-2 infection; receipt of any medications or vaccinations intended to prevent COVID-19; any positive test results for hepatitis B virus, hepatitis C virus, or human immunodeficiency virus at screening; an immunocompromised condition; a history of autoimmune diseases; a history of malignancy within 5 years; a history of bleeding disorder or thrombocytopenia; receipt of immunoglobulins and/or any blood products within the previous 12 weeks; pregnancy or breast feeding.

This study consisted of two stages, and a stepwise approach was adopted as a safety precaution (figure S1). In the first stage, 40 participants aged 19 to 55 years in a low-dose cohort were initially block-randomized in a 2:1:1 ratio to the 10 μg GBP510 adjuvanted with AS03 (group 1), 10 μg unadjuvanted GBP510 (group 2), or placebo on a two-dose schedule. Subsequently, an additional 40 participants aged 19 to 55 years in a high-dose cohort were randomly assigned in a 2:1:1 ratio to the 25 μg GBP510 adjuvanted with AS03 (group 3), 25 μg unadjuvanted GBP510 (group 4), or placebo. In each dose level cohort, post-vaccination 7-day safety data of the first sentinel eight participants were reviewed by the independent data safety monitoring board (DSMB) before vaccination of the remaining 32 participants. In the second stage, 240 healthy younger and older adults aged 19 to 85 years were enrolled, and they were randomly assigned in a 2:2:1:1 ratio to 10 μg GBP510 adjuvanted with AS03 (group 1), 25 μg GBP510 adjuvanted with AS03 (group 3), 25 μg unadjuvanted GBP510 (group 4), or placebo on a two-dose schedule. At each time point before dose escalation, second-dose vaccination, or advancement to the next stage, post-vaccination 7-day safety data were reviewed by the DSMB.

The trial was designed by SK Bioscience with support from the Coalition for Epidemic Preparedness Innovations (CEPI). The trial protocol was approved by the Institutional Review Board (IRB) of each participating hospital and was performed in accordance with the International Council for Harmonization Good Clinical Practice guidelines. Written informed consent was obtained from all participants. This study was registered with ClinicalTrials.gov (NCT04750343).

### Study vaccine, adjuvant, and procedures

The study antigen GBP510 is a novel vaccine candidate, which contains self-assembling, two-component nanoparticles (RBD-16GS-I53-50) displaying the RBD of the SARS-CoV-2 spike protein. It was developed by the Institute for Protein Design at the University of Washington, and advanced by SK bioscience using structure-based vaccine design techniques. AS03 (α-tocopherol-containing oil-in-water emulsion manufactured by GlaxoSmithKline) was used as an adjuvant.^5^ GBP510 is expected to have an enhanced ability to induce an immune response with adjuvant, owing to its molecular structure enabling multivalent antigen presentation.^3,4^

For each participant, two-doses of GBP510 or placebo (0.5 mL injection volume) were injected into the deltoid muscle of the upper arm at a 28-day interval: SARS-CoV-2 RBD nanoparticle 10 μg or 25 μg per 0.5 mL for unadjuvanted GBP510; SARSCoV-2 RBD nanoparticle 10 μg or 25 μg per 0.25 mL plus 0.25 mL of AS03 for adjuvanted GBP510. Blood samples were drawn from participants in all groups, and clinical assessments for safety were performed (Figure 1).

**Figure 1.**
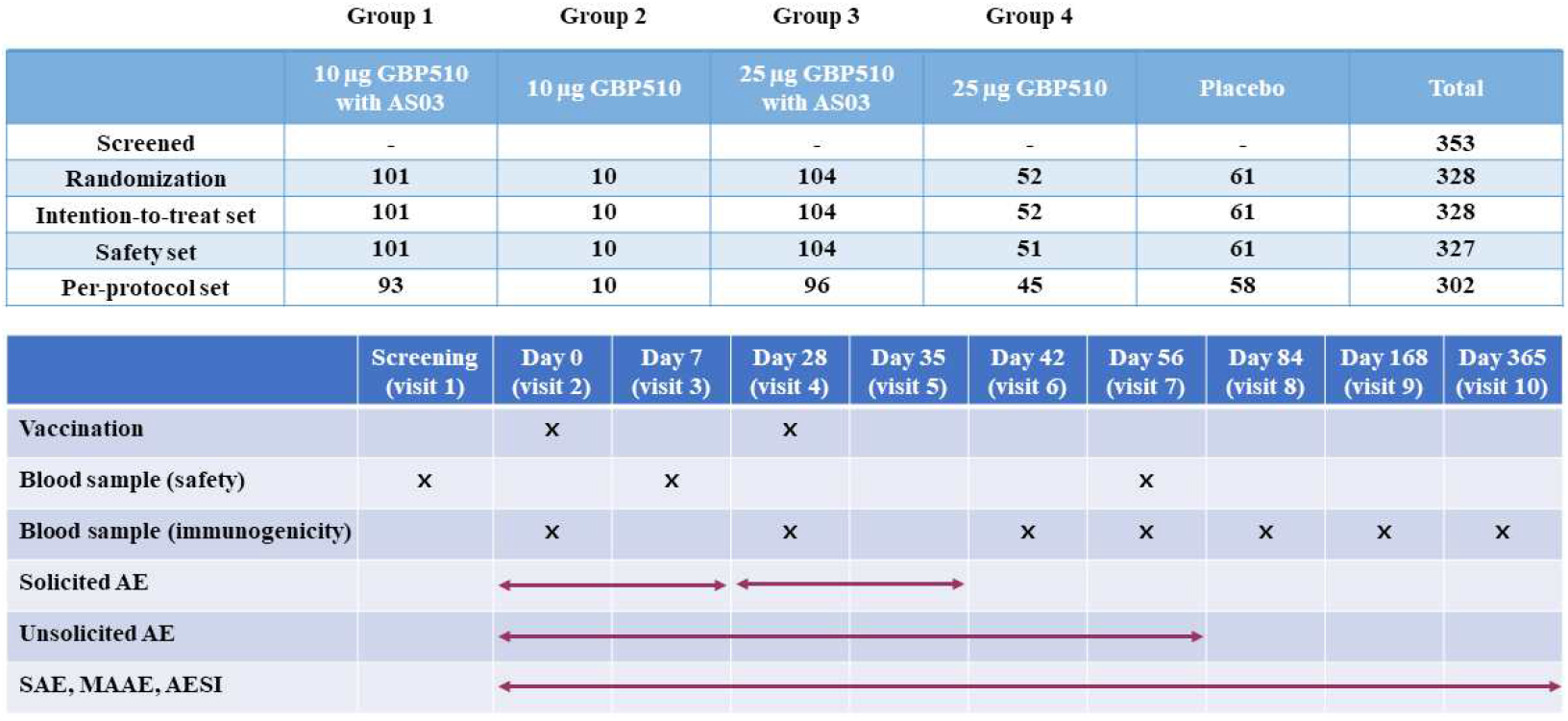
Vaccine trial groups and timeline.

### Safety assessment

Participants from each group were instructed to complete a paper or electronic diary to record solicited local and systemic adverse events (AEs) for 7 days after each dose. Laboratory values (blood chemistry and hematology tests) were monitored on baseline (day 0), and 28th day after the second vaccination. Sentinel groups were additionally evaluated on the 7th day after the first vaccination for the laboratory values. In addition, participants were instructed to record all unsolicited AEs that occurred during 28 days after each vaccination, using the paper/electronic diary or telephone contact. Serious AEs (SAEs), medically attended AEs (MAAEs), AEs of special interest (AESIs) including potential immune-mediated diseases (pIMDs)^6^ are collected throughout the study, from the first dose vaccination (day 0) until 12 months after the after the second dose (Table S1 and S2). The results of the interim analysis are presented here including the safety data until 28 days after the second dose (Table S3 - S7). The severity of solicited local and systemic AEs was assessed using the US Food and Drug Administration (FDA) toxicity grading scale.^7^

### Immunogenicity assessment

The IgG antibody response to the SARS-CoV-2 RBD was measured via enzyme-linked immunosorbent assay (ELISA) before the administration (day 0) of vaccine or placebo, at 28 days after the first dose (day 28), at 14 days after the second dose (day 42), and 28 days after the second dose (day 56). The neutralizing antibody responses to SARS-CoV-2 were measured by pseudovirus (at days 0, 28, 42 and 56) and wild-type virus neutralization assays (at days 0 and 42). Additionally, cell-mediated responses were assessed at days 0, 28 and 42. The titers of ELISA and neutralization assay were converted to binding antibody unit (BAU)/mL and international unit (IU)/mL using WHO international standard (NIBSC 20/136)^8^ and compared with the HCS of WHO representative panel.

The pseudovirus-based neutralization assay (PBNA) is a lentiviral vector-based assay used to measure neutralizing antibody titers in polyclonal sera. Pseudovirus containing the luciferase reporter gene was made with spike envelope protein, which is the surface proteins of the target virus. After pseudovirus infection, the neutralizing antibody titer was calculated by measuring the 50% inhibition level of reporter enzyme from infected cells. In addition, a plaque reduction neutralization test (PRNT) was performed, in a subset of participants, using wild-type SARS-CoV-2 (BetaCoV/Korea/KCDC03/2020) to measure the 50% reduction plaque level of neutralizing antibodies against SARS-CoV-2 as described previously.^9^

Cell-mediated immune responses were measured by intracellular cytokine staining using SARS-CoV-2 RBD to restimulate T cells. Type 1 helper T (Th1) cell responses are characterized by the expression of interferon (IFN)-γ, tumor necrosis factor (TNF)-α, and interleukin (IL)-2, while type 2 helper T (Th2) cell responses are characterized by the expression of IL-4 and IL-5.

### Statistical analysis

This study was not designed to test any hypothesis. Thus, sample size was calculated based on probability of detecting rare AEs with pre-determined sample size. Each test group in this study was planned to comprise 10, 50, or 100 participants, depending on the dose level and presence of adjuvant. Assuming a dropout rate of approximately 10%, at least 9, 45, or 90 evaluable participants were anticipated for each test group. Safety outcomes were analyzed for participants in the safety set who received at least one dose of the study intervention. Safety outcomes were described as frequencies (%) with 95% confidence intervals (CIs). In contrast, immunogenicity analyses were performed using the per-protocol set. SARS-CoV-2 RBD IgG concentrations as geometric mean concentrations (GMCs), and SARS-CoV-2 neutralizing antibody titers as geometric mean titers (GMTs), are presented along with standard deviations. The GMCs and GMTs were calculated as the mean of the assay results after the logarithmic transformation was made and exponentiated to the original scale. In addition, geometric mean fold rise (GMFR) and seroconversion rate (percentage of participants with ≥ 4-fold rise from baseline) were calculated at each time point (at 28 days after the first dose, and at 14 days and 28 days after the second dose) compared to baseline. For cell-mediated immunity, the percentage of CD4+ T cells expressing each cytokine, median, and quartiles were plotted. Two sample t-test were performed to compare the GMTs and Chi-square test or Fisher’s exact test were performed to compare the seroconversion rates between groups using SAS version 9.4.

## RESULTS

### Characteristics of study participants

Between February 2 and July 28, 2021, a total of 353 subjects were screened at 14 clinical trial sites in South Korea. Among them, 328 participants were randomly assigned to group 1 (n = 101), group 2 (n = 10), group 3 (n = 104), group 4 (n = 52), and placebo group (n = 61) (figure 1). Among 328 randomized participants, 320 participants (97.56%) completed the 28-day follow-up after the second dose vaccination, excluding eight participants: due to AE (breast cancer diagnosed after enrollment not related to vaccine, one in group 4), lost to follow-up (one in group 1), discretion by the investigator because of poor compliance (one in group 1), request for early termination by the participants (one in groups 1, 3, and placebo group; two in group 4). Among 328 randomized participants in the intention-to-treat set (ITT set), one was excluded and 327 received at least 1-dose vaccination, and those were included in the safety set. Of the 327 participants in the safety set, 302 completed the vaccination schedule without major protocol deviations, and these were included in the per-protocol set (PP set).

The demographic characteristics are presented in table 1. The median age of the ITT set was 42 years. 89.3% (293/328) of participants were aged 19–64 years and 10.7% (35/328) were elderly (≥ 65 years). Among the 328 participants, 45.1% (148) were men and 54.9% (180) were women. The mean body mass index (BMI) at screening was 23.9 (± 2.7) kg/m^2^.

**Table 1.** Demographic characteristics of participants at enrollment.

|  | **GBP510 10 µg**<br>**with AS03**<br>**(N = 101)** | **GBP510 10 µg**<br>**(N = 10)** | **GBP510 25 µg**<br>**with AS03**<br>**(N = 104)** | **GBP510 25 µg**<br>**(N = 52)** | **Placebo**<br>**(N = 61)** | **Total**<br>**(N = 328)** |
| --- | --- | --- | --- | --- | --- | --- |
| Age (years) |  |  |  |  |  |  |
| Mean ± SD | 45.0 ± 14.9 | 38.7 ± 8.7 | 43.9 ± 14.2 | 43.2 ± 15.3 | 43.3 ± 14.7 | 43.9 ± 14.5 |
| Median (range) | 44.0<br>(21.0–76.0) | 40.1<br>(24.0–50.0) | 42.0<br>(19.0–74.0) | 42.0<br>(19.0–73.0) | 42.0<br>(19.0–80.0) | 42.0<br>(19.0–80.0) |
| Age, n (%) |  |  |  |  |  |  |
| 19–64 years | 88 (87.1) | 10 (100) | 92 (88.5) | 47 (90.4) | 56 (91.8) | 293 (89.3) |
| 65–85 years | 13 (12.9) | 0 (0) | 12 (11.5) | 5 (9.6) | 5 (8.2) | 35 (10.7) |
| Sex, n (%) |  |  |  |  |  |  |
| Male | 51 (50.5) | 3 (30.0) | 45 (43.3) | 23 (44.2) | 26 (42.6) | 148 (45.1) |
| Female | 50 (49.5) | 7 (70.0) | 59 (56.7) | 29 (55.8) | 35 (57.4) | 180 (54.9) |
| BMI (kg/m<sup>2</sup>) |  |  |  |  |  |  |
| Mean ± SD | 24.0 ± 2.8 | 23.3 ± 2.3 | 24.1 ± 2.6 | 23.8 ± 2.9 | 23.8 ± 2.6 | 23.9 ± 2.7 |
| Median (range) | 24.2<br>(18.4–29.9) | 23.0<br>(20.2–26.6) | 23.8<br>(18.0–29.6) | 24.2<br>(18.4–29.9) | 24.0<br>(18.0–28.2) | 24.1<br>(18.0–29.9) |

SD, standard deviation; BMI, body mass index

### Safety outcomes

Of the 327 participants in the safety set, solicited local AEs within 7 days after any vaccination occurred in 89.1% (90/101 participants) in group 1, 50.0% (5/10 participants) in group 2, 92.3% (96/104 participants) in group 3, 66.7% (34/51 participants) in group 4, and 21.3% (13/61 participants) in the placebo group. Solicited systemic AEs within 7 days after any vaccination occurred in 85.2% (86/101 participants) of group 1, 70.0% (7/10 participants) of group 2, 85.6% (89/104 participants) of group 3, 70.6% (36/51 participants) of group 4, and 57.4% (35/61 participants) of the placebo group. Both local and systemic AEs were more common in the groups receiving the AS03-adjuvanted formulation (groups 1 and 3). Most local and systemic AEs were mild (grade 1) to moderate (grade 2) in severity (Figure 2 and Table 2) and transient. For solicited local reactions, the frequency of pain at injection site was similar after the first and second dose vaccination irrespective of vaccine composition (Table 2), while redness tended to increase post-dose 2 in groups 1 and 3. The ratio of Grade 3 solicited local AEs were 1.9% (2/103 events) only in group 3 after dose 1, but those accounted for 2.1% (2/95 events) and 1.9% (2/107 events) for group 1 and 3 after dose 2. In participants vaccinated with GBP510 adjuvanted with AS03 (groups 1 and 3), the frequency and intensity of systemic AEs was higher after the second dose as compared to the first dose vaccination. No grade 3 solicited systemic AEs were reported after the first dose, while grade 3 systemic events were reported after the second dose: The ratio of Grade 3 systemic AE were 2.1% (6/293 events) for group 1, 4.7% (16/339 events) for group 3 and 1.4% (1/71 events) for placebo. No grade 4 (potentially life-threatening) AE was observed.

**Figure 2.**
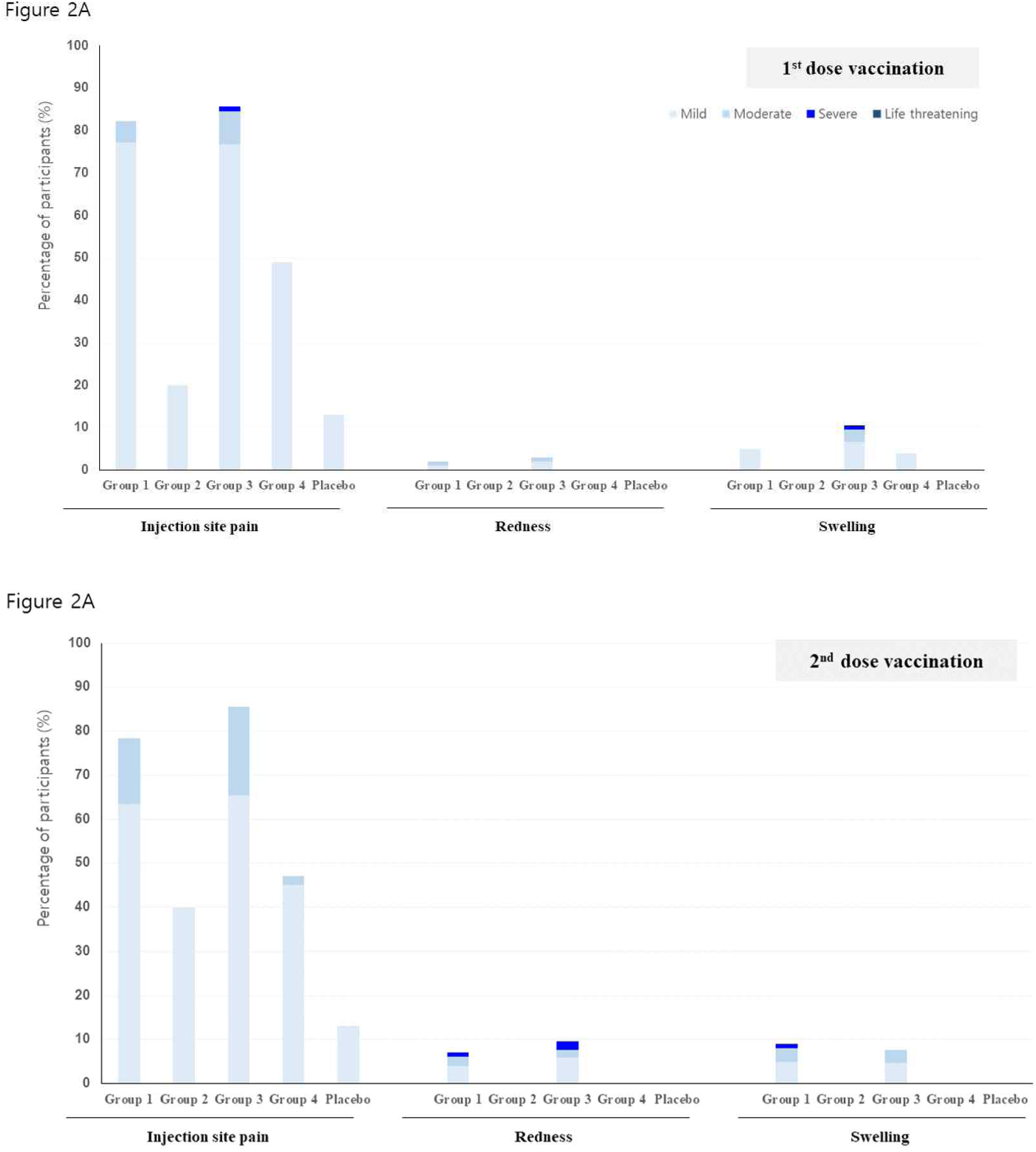
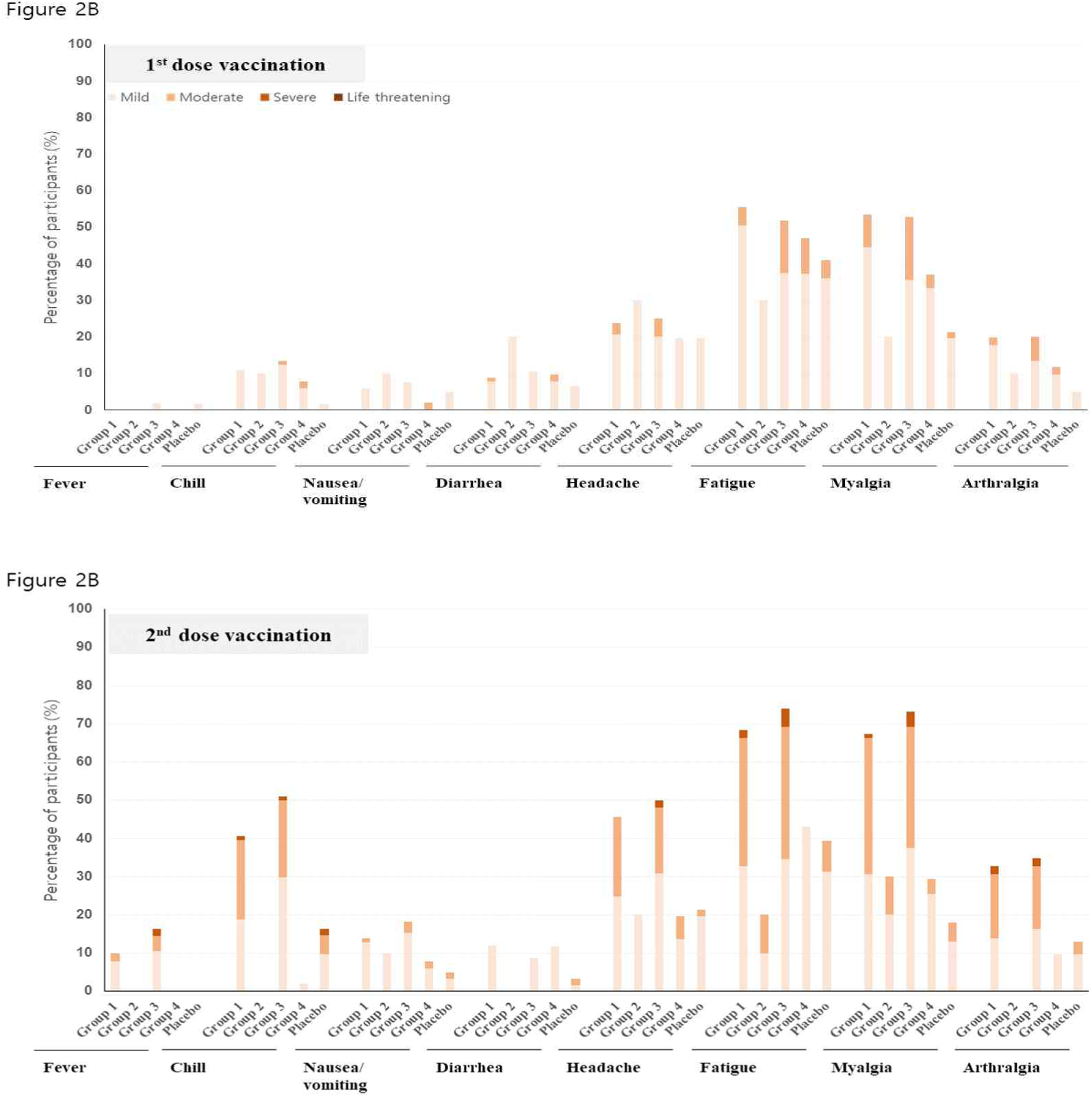
Solicited local (A) and systemic (B) adverse events within 7 days after first-dose and second-dose vaccination. Data on adverse events were recorded in a paper or an electronic diary for 7 days after each injection. Group 1, 10 μg GBP510 adjuvanted with AS03; group 2, 10 μg unadjuvanted GBP510; group 3, 25 μg GBP510 adjuvanted with AS03; group 4, 25 μg unadjuvanted GBP510. Each adverse event was graded as mild (grade 1, does not interfere with activity), moderate (grade 2, interferes with activity), severe (grade 3, prevents daily activity), or potentially life-threatening (grade 4, led to an emergency department visit or hospitalization). Redness and swelling were graded as mild (2.0 to 5.0 cm in diameter), moderate (> 5.0 to 10.0 cm in diameter), severe (> 10.0 cm in diameter), or potentially life-threatening (necrosis or exfoliative dermatitis for redness and necrosis for swelling). Fever was graded as mild (38.0–38.4°C), moderate (38.5–38.9°C), severe (39.0–40.0°C), or potentially life-threatening (> 40.0°C).

**Table 2.** Solicited local and systemic adverse events by severity (safety set)

|  | GBP510 10 µg<br>with AS03<br>(N = 101) | GBP510 10 µg<br>(N = 10) | GBP510 25 µg with<br>AS03<br>(N = 104) | GBP510 25 µg<br>(N = 51) | Placebo<br>(N = 61) |
| --- | --- | --- | --- | --- | --- |
| Total No. of local AEs after any vaccine injection | 185 | 6 | 210 | 51 | 16 |
| Maximum severity, No. of AEs, n (%) |  |  |  |  |  |
| Grade 1 (mild) | 157 (84.9) | 6 (100) | 168 (80.0) | 50 (98.0) | 16 (100) |
| Grade 2 (moderate) | 26 (14.1) | 0 (0) | 38 (18.1) | 1 (2.0) | 0 (0) |
| Grade 3 (severe) | 2 (1.1) | 0 (0) | 4 (1.9) | 0 (0) | 0 (0) |
| Grade 4 (life threatening) | 0 (0) | 0 (0) | 0 (0) | 0 (0) | 0 (0) |
| Total No. of local AEs after the 1st vaccination | 90 | 2 | 103 | 27 | 8 |
| Maximum severity, No. of AEs, n (%) |  |  |  |  |  |
| Grade 1 (mild) | 84 (93.3) | 2 (100) | 89 (86.4) | 27 (100) | 8 (100) |
| Grade 2 (moderate) | 6 (6.7) | 0 (0) | 12 (11.7) | 0 (0) | 0 (0) |
| Grade 3 (severe) | 0 (0) | 0 (0) | 2 (1.9) | 0 (0) | 0 (0) |
| Grade 4 (life threatening) | 0 (0) | 0 (0) | 0 (0) | 0 (0) | 0 (0) |
| Total No. of local AEs after the 2nd vaccination | 95 | 4 | 107 | 24 | 8 |
| Maximum severity, No. of AEs, n (%) |  |  |  |  |  |
| Grade 1 (mild) | 73 (76.8) | 4 (100) | 79 (73.8) | 23 (95.8) | 8 (100) |
| Grade 2 (moderate) | 20 (21.1) | 0 (0) | 26 (24.3) | 1 (4.2) | 0 (0) |
| Grade 3 (severe) | 2 (2.1) | 0 (0) | 2 (1.9) | 0 (0) | 0 (0) |
| Grade 4 (life threatening) | 0 (0) | 0 (0) | 0 (0) | 0 (0) | 0 (0) |
| Total No. of systemic<br>AEs after any vaccine injection | 473 | 21 | 531 | 131 | 133 |
| Maximum severity, No. of AEs, n (%) |  |  |  |  |  |
| Grade 1 (mild) | 315 (66.6) | 19 (90.5) | 336 (63.3) | 115 (87.8) | 112 (84.2) |
| Grade 2 (moderate) | 152 (32.1) | 2 (9.5) | 179 (33.7) | 16 (12.2) | 20 (15.0) |
| Grade 3 (severe) | 6 (1.3) | 0 (0) | 16 (3.0) | 0 (0) | 1 (0.8) |
| Grade 4 (life threatening) | 0 (0) | 0 (0) | 0 (0) | 0 (0) | 0 (0) |
| Total No. of systemic AEs after the 1st vaccination | 180 | 13 | 192 | 69 | 62 |
| Maximum severity, No. of AEs, n (%) |  |  |  |  |  |
| Grade 1 (mild) | 160 (88.9) | 13 (100) | 145 (75.5) | 58 (84.1) | 58 (93.6) |
| Grade 2 (moderate) | 20 (11.1) | 0 (0) | 47 (24.5) | 11 (15.9) | 4 (6.5) |
| Grade 3 (severe) | 0 (0) | 0 (0) | 0 (0) | 0 (0) | 0 (0) |
| Grade 4 (life threatening) | 0 (0) | 0 (0) | 0 (0) | 0 (0) | 0 (0) |
| Total No. of systemic AEs after the 2nd<br>vaccination | 293 | 8 | 339 | 62 | 71 |
| Maximum severity, No. of AEs, n (%) |  |  |  |  |  |
| Grade 1 (mild) | 155 (52.9) | 6 (75.0) | 191 (56.3) | 57 (91.9) | 54 (76.1) |
| Grade 2 (moderate) | 132 (45.1) | 2 (25.0) | 132 (38.9) | 5 (8.1) | 16 (22.5) |
| Grade 3 (severe) | 6 (2.1) | 0 (0) | 16 (4.7) | 0 (0) | 1 (1.4) |
| Grade 4 (life threatening) | 0 (0) | 0 (0) | 0 (0) | 0 (0) | 0 (0) |

AE, adverse event
%: n / (No. of AEs in Safety Set by group) \* 100
n = number of adverse events

As for each local AE, injection site pain was the most frequently reported solicited local AE after any vaccination, but was mostly mild in severity, lasting for a mean of less than 3 days across groups. It was more frequent in GBP510 adjuvanted with AS03 recipients (88.1% of participants in group 1 and 92.3% of participants in group 3) than in the unadjuvanted vaccine recipients (50.0% of participants in group 2 and 66.7% of participants in group 4) and one case was reported as Grade 3 in group 3 after the first dose only. Injection site redness and swelling were rarely reported (less than 15% of participants across groups after any vaccination), which persisted for a mean of 1.4–2.7 days after the first dose and a mean of 2.9–3.7 days after the second dose vaccination, respectively. (Figure 2 and Table S3). For the solicited systemic AEs, fatigue was the most common, followed by myalgia and headache (Figure 2 and Table S4). Fatigue and myalgia were two most frequently reported systemic AEs and more frequently reported in GBP510 adjuvanted with AS03 recipients (79.2% and 78.2% of participants in group 1; 75.0% and 79.8% of participants in group 3, respectively) than in the unadjuvanted vaccine recipients (40.0% and of 40.0% of participants in group 2 and 60.8% and 47.1% of participants in group 4) after any vaccination. Headache was also more common in GBP510 adjuvanted with AS03 group however, it was less frequent than fatigue and myalgia (53.47% in group1 and 56.73% in group 3 after any vaccination). Of note, most solicited systemic AEs were mild and moderate in severity after the first dose, while more moderate and severe solicited AEs occurred after the second dose (Table S4).

As for the laboratory values, there was no clinically significant change until 7 days after the first dose vaccination in sentinel group participants. Through 28 days after the second dose vaccination, clinically significant abnormalities occurred in two participants (one in group 1 and one in the placebo group) with hematologic test and nine participants (one in group 1, three in group 3, one in group 4, and four in placebo group) with blood biochemistry test among all study participants (Tables S5 and S6). Four abnormal laboratory values showed causal relation with GBP510 administration (mild leukopenia, 1 case; change in triglyceride level, 3 cases). Abnormal laboratory values were not associated with any clinical manifestations.

Unsolicited AEs within 28 days after any vaccination were reported in 19.8% (20/101, 26 events) in group 1, 20.0% (2/10, 3 events) in group 2, 18.3% (19/104, 31 events) in group 3, 19.6% (10/51, 16 events) in group 4, and 16.4% (10/61, 16 events) in the placebo group (table S7). Most of them were mild and few were related to GBP510 vaccination: 6.93% (7/101 participants) in group 1, 0% (0/10 participants) in group 2, 4.81% (5/104 participants) in group 3, 1.96% (1/51 participants) in group 4, and 3.28% (2/51 participants) in the placebo group. MAAEs within 28 days after any vaccination were reported in 8.9% (9/101 participants, 12 events) in group 1, 20.0% (2/10 participants, 2 events) in group 2, 8.7% (9/104 participants, 9 events) in group 3, 9.8% (5/51 participants, 8 events) in group 4, 6.6% (4/61 participants, 5 events) in the Placebo group. SAEs within 28 days after vaccination were reported in 1.0% (1/104 participants) in group 3 and 3.9% (2/51 participants) in group 4; they were not considered related to the vaccine. No AESI, including pIMDs, was reported within 28 days after any vaccination. Compared with the whole safety set, reactogenicity profile was higher in younger adults aged 19 - 64 years than in elderly aged 65 years or older. (Figure S3).

### Immunogenicity outcomes

Immunogenicity was analyzed in 302 participants in the PP group. GMCs of anti-SARS-CoV-2-RBD IgG antibody ranged from 11.0 to 16.3 BAU/mL at baseline (day 0), and those increased to 111.9/129.0 BAU/mL in GBP510 adjuvanted with AS03 recipients (group 1/group 3) and 37.7/28.6 BAU/mL in unadjuvanted GBP510 recipients (group 2/group 4) by 28 days after the first dose, respectively (figure 3 and table S8). By day 42 (14 days after second dose), GMCs of anti-SARS-CoV-2-RBD IgG antibody had further increased to 2163.6/2599.2 BAU/mL in GBP510 adjuvanted with AS03 recipients (group 1/group 3) and 155.3/112.4 BAU/mL in unadjuvanted GBP510 recipients (group 2/group 4). GMCs of GBP510 adjuvanted with AS03 recipients (group 1 and group 3) were 3.0–4.5 times higher than those of unadjuvanted GBP510 recipients (group2 and 4) at days 28 and these ratios increased to 13.9 and 23.1 times at 42, respectively (P <0.0001 for group 1 vs 2 and group 3 vs 4 at days 28 and 42). Seroconversion rates of IgG antibody were also higher in GBP510 adjuvanted with AS03 recipients than in the non-adjuvanted GBP510 recipients at day 28 and reached 100% (group 1), 100% (group 2), 100% (group 3), and 64.4% (group 4) on day 42.

**Figure 3.**
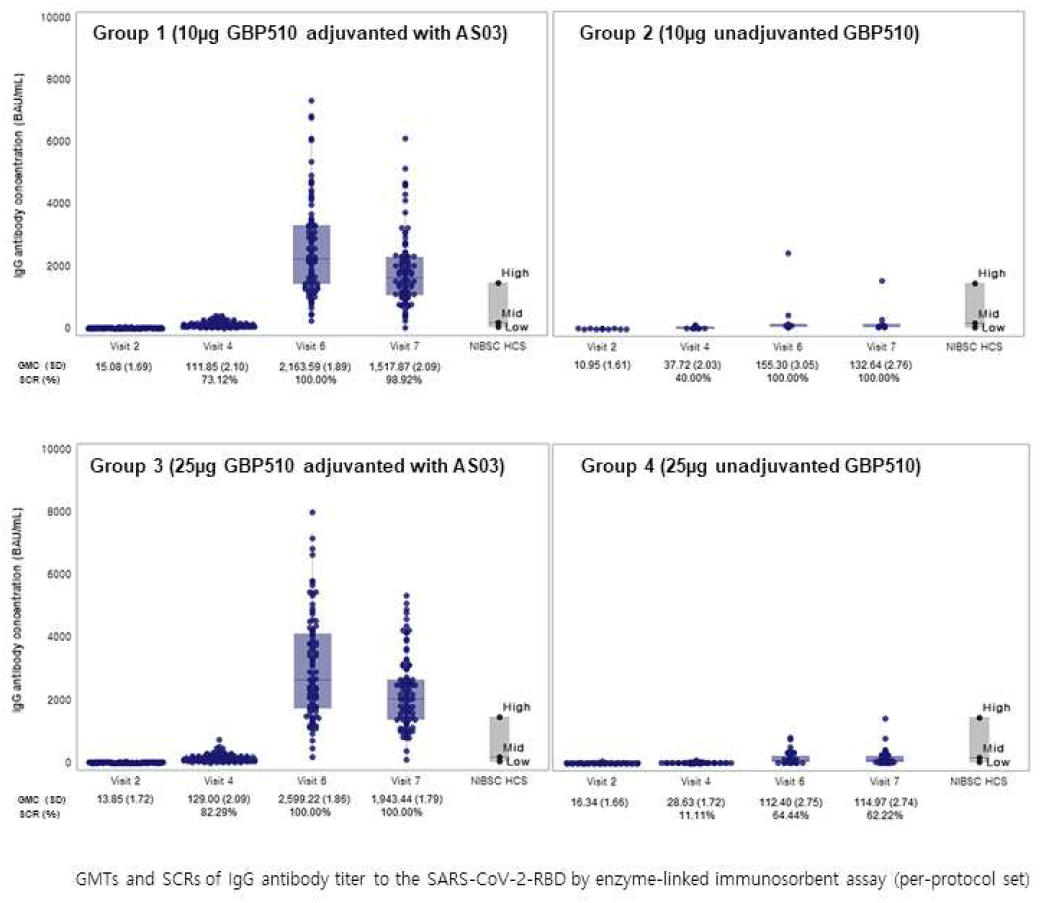
Anti-SARS-CoV-2 receptor-binding domain (RBD) IgG antibody response. Geometric mean concentrations of anti-SARS-CoV-2 RBD IgG were compared from baseline (day 0) to day 56 (28 days after the second dose). BAU, binding antibody unit; GMC, Geometric mean concentration; SD, standard deviation; SCR, seroconversion rate; NIBSC, National Institute for Biological Standards and Control; HCS, human convalescent serum

As for the neutralizing antibody titers by PBNA, GMTs ranged from 18.1 to 21.0 IU/mL at baseline (day 0), and those increased to 40.0/40.9 IU/mL in GBP510 adjuvanted with AS03 recipients (group 1/group 3) and 32.8/21.2 IU/mL in unadjuvanted GBP510 recipients (group 2/group 4) by 28 days after the first dose, respectively (P<0.0001 for group 3 vs. group 4; figure 4 and table S9). At day 42 (14 days after the second dose vaccination), the GMTs were more than 10 times higher in GBP510 adjuvanted with AS03 recipients (group 1/group 3) compared to unadjuvanted GBP510 recipients (group 2/group 4): 1369.0/1431.5 IU/mL versus 83.5/63.9 IU/mL (P <0.0002 for group 1 vs. group 2 and P<0.0001 for group 3 vs. group 4). Two-dose vaccination with 10 μg or 25 μg GBP510 adjuvanted with AS03 induced similar magnitudes of neutralizing antibody response and seroconversion rates (100% in group 1; 99.0% in group 3).

**Figure 4.**
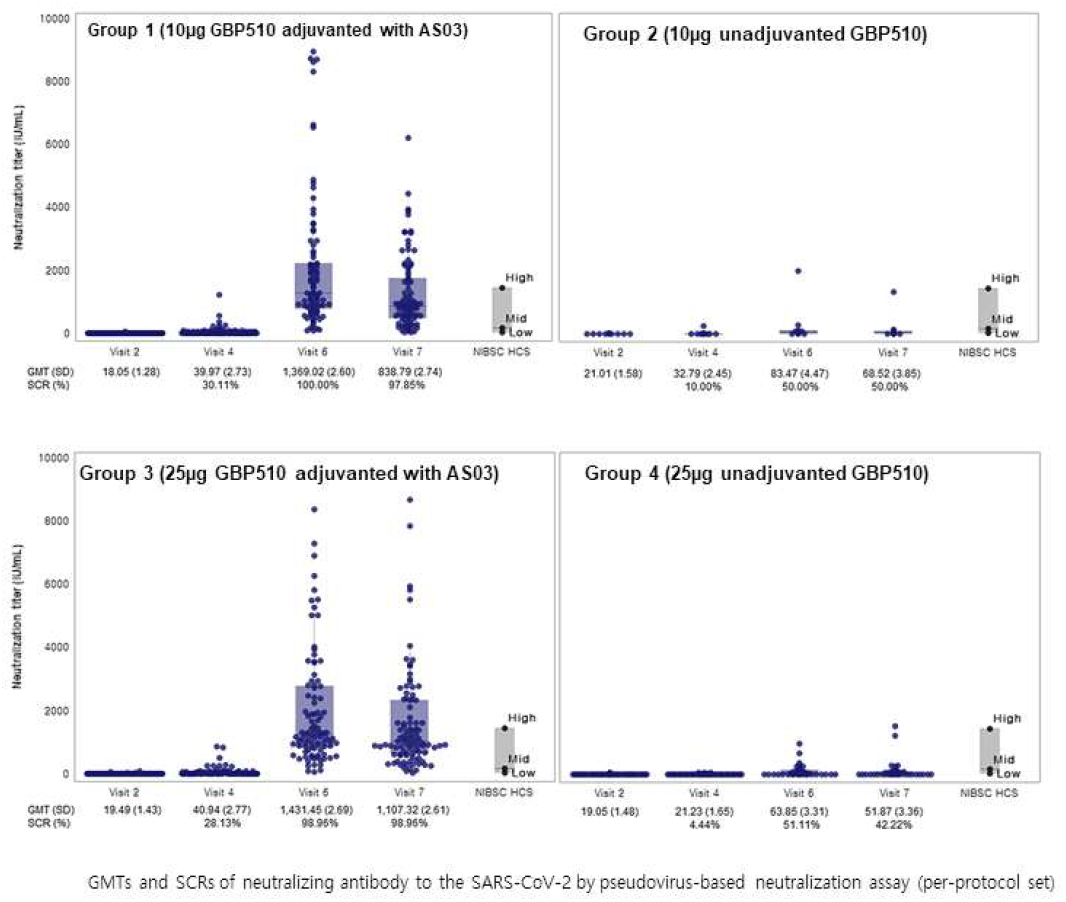
Neutralizing antibody response measured by pseudovirus-based neutralization assay (PBNA). Geometric mean neutralizing antibody titers were compared from baseline (day 0) to day 56 (28 days after the second dose). The serum neutralization titers were defined as the 50% maximal inhibitory concentration (IC_50_). GMT, Geometric mean titer; SD, standard deviation; SCR, seroconversion rate; NIBSC, National Institute for Biological Standards and Control; HCS, human convalescent serum

In addition to PBNA, PRNT was performed for selected 76 participants, including 23 participants in group 1, 4 participants in group 2, 21 participants in group 3, 9 participants in group 4, and 19 participants in the placebo group. GMTs ranged from 4.3 to 9.0 IU/mL at baseline (day 0), and those increased to 949.8/861.0 IU/mL in GBP510 adjuvanted with AS03 recipients (group 1/group 3), 34.1/58.1 IU/mL in unadjuvanted GBP510 recipients (group 2/group 4), and 4.8 IU/mL in the placebo group by day 42, respectively (Figure 5 and Table S10). Two-dose vaccination with GBP510 adjuvanted with AS03 induced higher magnitudes of neutralizing antibody response and seroconversion rates compared to unadjuvanted GBP510 (100% in group 1, 50.0% in group 2, 100% in group 3, and 77.8% in group 4).

**Figure 5.**
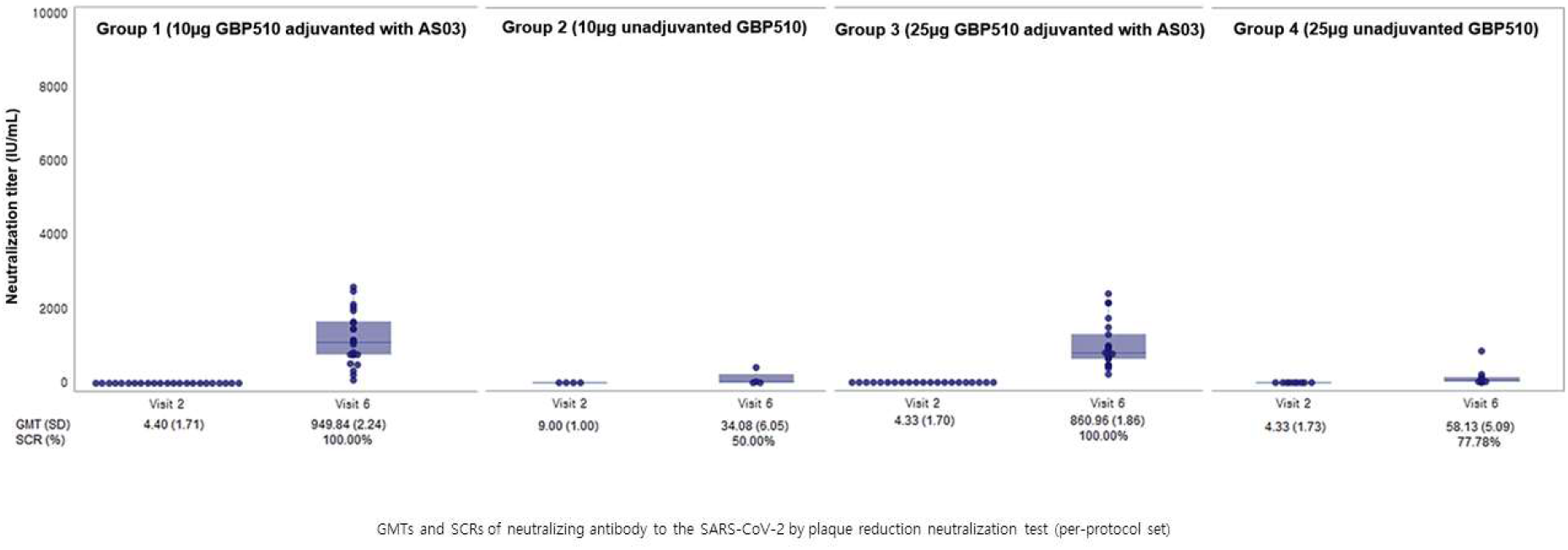
Neutralizing antibody response measured by plaque reduction neutralization test (PRNT). Geometric mean neutralizing antibody titers were compared from baseline (day 0) to day 42 (14 days after the second dose). The serum neutralization titers were defined as the median neutralizing titer (ND_50_). GMT, Geometric mean titer; SD, standard deviation; SCR, seroconversion rate.

Cell-mediated immune responses were evaluated in randomly selected 60 participants (group 1, 19; group 2, 2; group 3, 17; group 4, 7; placebo group, 15) in a blinded manner. GBP510 adjuvanted with AS03 induced stronger CD4+ T-cell responses with higher percentages of IFN-γ, TNF-α, and IL-2 expression compared to unadjuvanted GBP510. However, IL-4 was inconsistent and IL-5 was nearly inexistent across all groups (Figure S2 and Table S15). Thus, data suggest noticeable level of Th1 response in GSBP510 adjuvanted with AS03 however, additional experiments are needed.

## DISCUSSION

The interim analyses (data until 4 weeks after the second dose) of safety and immunogenicity showed that GBP510 adjuvanted with AS03 had an acceptable safety profile and induced high immunogenicity after two-dose vaccination with either a low (10 μg) or high (25 μg) antigen dose. Most AEs were mild and moderate in severity and self-limiting. No vaccination-related SAEs and AESIs were observed. Both low- and high-dose GBP510 adjuvanted with AS03 induced potent immune response with neutralizing antibodies approaching 99% seroconversion. Similar to other COVID-19 vaccines, injection site pain, fatigue, myalgia and headache were the most common solicited AEs.^11-13^ Reactogenicity tended to increase (frequency and intensity) after the second dose as compared to the first-dose^11,13^ and in younger adults as compared to older adults, also in line with what is observed with other COVID-19 vaccines.^23^

Although a threshold of immune correlates of protection against COVID-19 have not yet been established, neutralizing antibodies play an essential role in preventing COVID-19, while cellular immunity is beneficial to limit disease severity. In this trial, we found that two doses of 10 μg or 25 μg GBP510 adjuvanted with AS03 induced high-level of neutralizing antibodies. In addition, immunogenicity of GBP510 adjuvanted with AS03 groups were compared with WHO representative anti-SARS-CoV-2 antibody titres of human convalescent serum (NIBSC code low: 20/140, mid: 20/148, and high: 20/150) calibrated against the WHO International Standard for anti-SARS-CoV-2 immunoglobulin (NIBSC code 20/136), and the result was also higher than the HCS of WHO representative panel.

When comparing the GMTs by PBNA, the titers for 10 μg or 25 μg GBP510 adjuvanted with AS03 on day 42 were 5.7-fold and 6.0-fold higher than the GMT of the WHO representative HCS (low: 44 IU/mL; mid: 210 IU/mL; and high: 1,473 IU/mL; calculated GMT value: 239 IU/mL), respectively (figure 4).^8^ Likewise, GMTs of 10 μg or 25 μg GBP510 adjuvanted with AS03 by PRNT were 4.0-fold and 3.6-fold higher than the GMT of the WHO representative human convalescent serum on day 42 (same GMT value of WHO representative HCS was used for PBNA and PRNT; 239 IU/mL). The GMCs of Anti-SARS-CoV-2-RBD IgG antibody were also 11.0-fold and 13.3-fold higher in 10 μg or 25 μg GBP510 adjuvanted with AS03 recipients on day 42 when compared to that of the WHO representative panel (low: 45 BAU/mL; mid: 205 BAU/mL; and high: 817 BAU/mL; calculated GMC: 196 BAU/mL) (figure 3).^8^ Thus, GBP510 adjuvanted with AS03 is highly immunogenic and may provide high-level protection against COVID-19.

When analyzed by age group, neutralizing antibody titers by PBNA and IgG antibody response by ELISA were relatively lower in older adults (≥ 65 years) compared to the younger, as seen with other COVID-19 vaccines (Table S11 - S14).^10,11^ In addition, when analyzed for all participants, 25 μg GBP510 adjuvanted with AS03, which is currently being tested in the following phase III clinical trials, resulted higher in neutralizing antibody titers by PBNA and IgG antibody response by ELISA compared to that of the 10 μg GBP510 adjuvanted with AS03 at the peak (two weeks after dose 2) and this trend was consistent through four weeks post-vaccination. Safety profile was comparable irrespective of the dosage.

Owing to the recombinant synthetic protein vaccine platform, GBP510 adjuvanted with AS03 is thermostable at 4°C and it can be distributed and stored at refrigerated temperatures; therefore, it has the advantage of being widely usable in developing countries where it is difficult to establish a cold chain for cryogenic distribution and storage (−80 to −20°C). GBP510 uses an immune focused, multi-array of recombinant RBD. RBD not only shares the multiple positive attributes with parental full-length spike protein inducing >6 months antibody persistency and high neutralizing activity, but also has more advantages on high-yielding manufacturing potential and affordable cost.^2^ As reported previously, RBD contains multiple conformational and conserved neutralizing epitopes, so vaccines targeting the RBD will likely induce cross-neutralization against variant viruses, including B.1.1.7 and B.1.617.^3,4,14^ In addition, AS03 might broaden protection with cross-reactive immunogenicity against variant viruses as experienced with AS03-adjuvanted influenza vaccines.^15,16^ PRNT was performed to evaluate the cross-neutralizing antibody immunity against the delta variant virus (B.1.617) for 11 randomly selected 25 μg GBP510 adjuvanted with AS03 recipients. The neutralizing antibody titer against the delta variant showed a 2.4-fold decrease compared to the wild type virus similar to other COVID-19 vaccines (Figure S4).^17,18^ Similar results were also shown with 11 participants limited to the age of 65 years or older who were randomly selected (Figure S4).

AS03-adjuvanted SARS-CoV-2 subunit vaccine induced durable robust neutralizing-antibody response in non-human primates up to 180 days after vaccination.^19^ Depending on the presence or absence of AS03, the immunogenicity of GBP510 showed a 10–30-fold difference in neutralizing antibody titers in this study. AS03 is an adjuvant system containing two biodegradable oils (α-tocopherol and squalene) in an oil-in-water emulsion.^5^ AS03 activates the innate immune response at the injection site and enhance adaptive immune responses (antibody and T-cell) to the vaccine antigen. Unlike MF59, another squalene-based adjuvant, AS03 additionally contains α-tocopherol, which modulates the expression of diverse cytokines, increases antigen loading in monocytes and the recruitment of granulocytes into the draining lymph nodes. Thus, α-tocopherol is an essential component of AS03 for achieving the highest antibody response.^16^ Similar to our results, AS03-adjuvanted influenza vaccines showed a superior immunogenicity compared to unadjuvanted influenza vaccines in a meta-analysis.^20^ In addition, an increased reactogenicity post-dose 2 as compared to post-dose 1 was observed with other AS03-containing COVID-19 vaccines.^21,22^

The limitations of this study include the small sample size, especially for older adults, and short follow-up period. Nevertheless, the sample size provides a meaningful initial assessment of the frequency and intensity of the solicited AEs. As of January 31, 2022 (6 months post-vaccination), no AESI nor SAE related to vaccine have been reported.

In conclusion, GBP510 adjuvanted with AS03 was well-tolerated with an acceptable safety profile, and highly immunogenic. The phase 1/2 trial has been extended to the homologous booster study, which assess the impact of a third dose (25μg GBP510 adjuvanted with AS03) on the durability and cross-protection. Based on the results of this phase 1/2 trial, a phase 3, randomized, observer-blind trial is underway to compare the immunogenicity and safety of 25 μg GBP510 adjuvanted with AS03 to ChAdOx1 in adults 18 years of age and older (initiated August 30, 2021). In addition, a Phase 2, heterologous GBP510 adjuvanted with AS03 booster trial began on January 6, 2022, to assess the merits of this immune-focused antigen as a universal booster to already licensed CoV-2 vaccines.

## Data Availability

All data produced in the present work are contained in the manuscript.

## Contributors

HJC, HK, JHR, SJL, HKP, ACW, LC, DV, NPK, F. Roman, M. A. Ceregido, F. Solmi, and A. Philippot contributed to the conception and design of this study. WSC, JYH, JSL, DSJ, SWK, KHP, JSE, SJJ, JL, KTK, HJC, JWS, YKK, and HJC were the principal investigators of the study site. JYS, WSC, JYH, JSL, DSJ, SWK, KHP, JSE, SJJ, JL, KTK, HJC, JWS, YKK, JYN, WJK and HJC contributed to the acquisition of the clinical and laboratory data. JYS, JYN, WJK, and HJC contributed to the interpretation of the data. JYS and HJC contributed to statistical analysis. JYS and HJC analyzed the data with responsibility for its integrity and prepared the manuscript. All authors reviewed the manuscript for intellectual content and approved the final draft for submission. All authors have full access to all the data in the study and accept responsibility to submit for publication.

## Acknowledgments

This study was supported by the Bill & Melinda Gates Foundation (BMGF) and the Coalition for Epidemic Preparedness Innovations (CEPI).

## Data sharing

Individual participant data will be made available when the trial is complete upon requests directed to the corresponding author. Proposals will be reviewed and approved by the sponsor, investigators, and collaborators on the basis of scientific merit. After approval of a proposal, data can be shared through a secure online platform.

## REFERENCES

1. World Health Organization. Coronavirus disease 2019 (COVID-19): situation report. Available at: https://www.who.int/emergencies/diseases/novel-coronavirus-2019/situation-reports (Accessed : 31 January 2022).

2. Kleanthous H, Silverman JM, Makar KW, Yoon IK, Jackson N, Vaughn DW. Scientific rationale for developing potent RBD-based vaccines targeting COVID-19. NPJ Vaccines 2021; 6(1): 128.

3. Walls AC, Fiala B, Schafer A, et al. Elicitation of Potent Neutralizing Antibody Responses by Designed Protein Nanoparticle Vaccines for SARS-CoV-2. Cell 2020; 183(5): 1367–82 e17.

4. Walls AC, Miranda MC, Schafer A, et al. Elicitation of broadly protective sarbecovirus immunity by receptor-binding domain nanoparticle vaccines. Cell 2021; 184(21): 5432–47 e16.

5. Morel S, Didierlaurent A, Bourguignon P, et al. Adjuvant System AS03 containing alpha-tocopherol modulates innate immune response and leads to improved adaptive immunity. Vaccine 2011; 29(13): 2461–73.

6. Tavares Da Silva F, De Keyser F, Lambert PH, Robinson WH, Westhovens R, Sindic C. Optimal approaches to data collection and analysis of potential immune mediated disorders in clinical trials of new vaccines. Vaccine 2013; 31(14): 1870–6.

7. Center for Biologics Evaluation and Research. Toxicity grading scale for healthy adult and adolescent volunteers enrolled in preventive vaccine clinical trials: guidance for industry.: Food and Drug Administration; 2007.

8. National Institute for Biological Standards and Control (NIBSC). WHO Reference Panel First WHO International Reference Panel for anti-SARS-CoV-2 immunoglubulin NIBSC code: 20/268 Instructions for use. Available at: https://www.nibsc.org/documents/ifu/20-268.pdf (Accessed : 2 January 2022)

9. Noh JY, Kwak JE, Yang JS, et al. Longitudinal Assessment of Antisevere Acute Respiratory Syndrome Coronavirus 2 Immune Responses for Six Months Based on the Clinical Severity of Coronavirus Disease 2019. J Infect Dis 2021; 224(5): 754–63.

10. Ramasamy MN, Minassian AM, Ewer KJ, et al. Safety and immunogenicity of ChAdOx1 nCoV-19 vaccine administered in a prime-boost regimen in young and old adults (COV002): a single-blind, randomised, controlled, phase 2/3 trial. Lancet 2021; 396(10267): 1979–93.

11. Walsh EE, Frenck RW, Jr., Falsey AR, et al. Safety and Immunogenicity of Two RNA-Based Covid-19 Vaccine Candidates. The New England journal of medicine 2020; 383(25): 2439–50.

12. Folegatti PM, Ewer KJ, Aley PK, et al. Safety and immunogenicity of the ChAdOx1 nCoV-19 vaccine against SARS-CoV-2: a preliminary report of a phase 1/2, single-blind, randomised controlled trial. Lancet 2020; 396(10249): 467–78.

13. Keech C, Albert G, Cho I, et al. Phase 1-2 Trial of a SARS-CoV-2 Recombinant Spike Protein Nanoparticle Vaccine. The New England journal of medicine 2020; 383(24): 2320–32.

14. Law JLM, Logan M, Joyce MA, et al. SARS-COV-2 recombinant Receptor-Binding-Domain (RBD) induces neutralizing antibodies against variant strains of SARS-CoV-2 and SARS-CoV-1. Vaccine 2021; 39(40): 5769–79.

15. Cohet C, van der Most R, Bauchau V, et al. Safety of AS03-adjuvanted influenza vaccines: A review of the evidence. Vaccine 2019; 37(23): 3006–21.

16. Garcon N, Vaughn DW, Didierlaurent AM. Development and evaluation of AS03, an Adjuvant System containing alpha-tocopherol and squalene in an oil-in-water emulsion. Expert Rev Vaccines 2012; 11(3): 349–66.

17. Uriu K, Kimura I, Shirakawa K, et al. Neutralization of the SARS-CoV-2 Mu Variant by Convalescent and Vaccine Serum. The New England journal of medicine 2021; 385(25): 2397–9.

18. Madhi SA, Baillie V, Cutland CL, et al. Efficacy of the ChAdOx1 nCoV-19 Covid-19 Vaccine against the B.1.351 Variant. The New England journal of medicine 2021; 384(20): 1885–98.

19. Arunachalam PS, Walls AC, Golden N, et al. Adjuvanting a subunit COVID-19 vaccine to induce protective immunity. Nature 2021; 594(7862): 253–8.

20. Hauser MI, Muscatello DJ, Soh ACY, Dwyer DE, Turner RM. An indirect comparison meta-analysis of AS03 and MF59 adjuvants in pandemic influenza A(H1N1)pdm09 vaccines. Vaccine 2019; 37(31): 4246–55.

21. Sridhar S, Joaquin A, Bonaparte MI, et al. Safety and immunogenicity of an AS03-adjuvanted SARS-CoV-2 recombinant protein vaccine (CoV2 preS dTM) in healthy adults: interim findings from a phase 2, randomised, dose-finding, multicentre study. Lancet Infect Dis 2022.

22. Ward BJ, Gobeil P, Seguin A, et al. Phase 1 randomized trial of a plant-derived virus-like particle vaccine for COVID-19. Nat Med 2021; 27(6): 1071–8.

23. Polack, Fernando P., et al. “Safety and Efficacy of the BNT162B2 Mrna Covid-19 Vaccine: Nejm.” New England Journal of Medicine, 31 Dec. 2020, https://www.nejm.org/doi/full/10.1056/nejmoa2034577.

