## Supplementary Appendix for "Safety and immunogenicity of a SARS-CoV-2 recombinant protein nanoparticle vaccine (GBP510) adjuvanted with AS03: a phase 1/2, randomized, placebo-controlled, observer-blinded trial"

**Table S1.** Adverse event of special interest (AESI) relevant to coronavirus disease 2019 (COVID-19)

| Body System | AESI relevant to COVID-19 |
|---|---|
| Immunologic | Enhanced disease following immunization<br>Anaphylaxis<br>Vasculitis |
| Respiratory | Acute Respiratory Distress Syndrome (ARDS) |
| Cardiac | Acute cardiac injury including:<br>▪ Microangiopathy<br>▪ Heart failure and cardiogenic shock<br>▪ Stress cardiomyopathy<br>▪ Coronary artery disease<br>▪ Arrhythmia<br>▪ Myocarditis, pericarditis |
| Hematologic | Coagulation disorder:<br>▪ Deep vein thrombosis<br>▪ Pulmonary embolus<br>▪ Cerebrovascular stroke<br>▪ Limb ischemia<br>▪ Hemorrhagic disease<br>▪ Thrombocytopenia |
| Renal | Acute kidney injury |
| Gastrointestinal | Liver injury |
| Neurologic | Guillain Barré Syndrome<br>Generalized convulsion<br>Anosmia, ageusia<br>Meningoencephalitis<br>Acute disseminated encephalomyelitis |
| Dermatologic | Chilblain-like lesions<br>Single organ cutaneous vasculitis<br>Erythema multiforme |

**Table S2.** List of potential immune-mediated diseases [6]

| Neuroinflammatory disorders | Musculoskeletal disorders | Skin disorders |
|---|---|---|
| ▪ Cranial nerve neuropathy, including paralysis and paresis (eg, Bell's palsy).<br>▪ Optic neuritis.<br>▪ Multiple sclerosis.<br>▪ Transverse myelitis.<br>▪ Guillain-Barré syndrome, including Miller Fisher syndrome and other variants.<br>▪ Acute disseminated encephalomyelitis, including site specific variants, eg, noninfectious encephalitis, encephalomyelitis, myelitis, myeloradiculoneuritis.<br>▪ Myasthenia gravis, including Lambert-Eaton myasthenic syndrome.<br>▪ Demyelinating peripheral neuropathies including:<br>- Chronic inflammatory demyelinating polyneuropathy.<br>- Multifocal motor neuropathy.<br>- Polyneuropathies associated with monoclonal gammopathy.<br>▪ Narcolepsy. | ▪ Systemic lupus erythematosus and associated conditions.<br>▪ Systemic scleroderma (systemic sclerosis), including:<br>- Diffuse scleroderma.<br>- CREST syndrome.<br>▪ Idiopathic inflammatory myopathies, including:<br>- Dermatomyositis.<br>- Polymyositis.<br>▪ Anti-synthetase syndrome.<br>▪ Rheumatoid arthritis and associated conditions including:<br>- Juvenile idiopathic arthritis.<br>- Still's disease.<br>▪ Polymyalgia rheumatica.<br>▪ Spondyloarthropathies, including:<br>- Ankylosing spondylitis.<br>- Reactive arthritis (Reiter's syndrome).<br>- Undifferentiated spondyloarthritis.<br>- Psoriatic arthritis.<br>- Enteropathic arthritis.<br>▪ Relapsing polychondritis.<br>▪ Mixed connective tissue disorder.<br>▪ Gout. | ▪ Psoriasis.<br>▪ Vitiligo.<br>▪ Erythema nodosum.<br>▪ Autoimmune bullous skin diseases (including pemphigus, pemphigoid, and dermatitis herpetiformis).<br>▪ Lichen planus.<br>▪ Sweet's syndrome.<br>▪ Localized scleroderma (morphea). |
| **Vasculitis** | **Blood disorders** | **Others** |
| ▪ Large vessels vasculitis including:<br>- Giant cell arteritis | ▪ Autoimmune hemolytic anemia.<br>▪ Autoimmune | ▪ Autoimmune glomerulonephritis including: |

| | | |
|---|---|---|
| (temporal arteritis).<br>- Takayasu's arteritis.<br>▪ Medium sized and/or small vessels vasculitis including:<br>- Polyarteritis nodosa.<br>- Kawasaki's disease.<br>- Microscopic polyangiitis.<br>- Wegener's granulomatosis (granulomatosis with polyangiitis).<br>- Churg–Strauss syndrome (allergic granulomatous angiitis or eosinophilic granulomatosis with polyangiitis).<br>- Buerger's disease (thromboangiitis obliterans).<br>- Necrotizing vasculitis (cutaneous or systemic).<br>- Anti-Neutrophil Cytoplasmic Antibody (ANCA) positive vasculitis (type unspecified).<br>- Henoch-Schonlein purpura (IgA vasculitis).<br>- Behcet's syndrome.<br>- Leukocytoclastic vasculitis. | thrombocytopenia.<br>▪ Antiphospholipid syndrome.<br>▪ Pernicious anemia.<br>▪ Autoimmune aplastic anemia.<br>▪ Autoimmune neutropenia.<br>▪ Autoimmune pancytopenia. | - IgA nephropathy.<br>- Glomerulonephritis rapidly progressive.<br>- Membranous glomerulonephritis.<br>- Membranoproliferative glomerulonephritis.<br>- Mesangioproliferative glomerulonephritis.<br>- Tubulointerstitial nephritis and uveitis syndrome.<br>▪ Ocular autoimmune diseases including:<br>- Autoimmune uveitis.<br>- Autoimmune retinitis.<br>▪ Autoimmune myocarditis.<br>▪ Sarcoidosis.<br>▪ Stevens-Johnson syndrome.<br>▪ Sjögren's syndrome.<br>▪ Alopecia areata.<br>▪ Idiopathic pulmonary fibrosis.<br>▪ Goodpasture syndrome.<br>▪ Raynaud's phenomenon. |
| Liver disorders | Gastrointestinal disorders | Endocrine disorders |
| ▪ Autoimmune hepatitis.<br>▪ Primary biliary cirrhosis.<br>▪ Primary sclerosing cholangitis. | ▪ Inflammatory bowel disease, including:<br>- Crohn's disease.<br>- Ulcerative colitis.<br>- Microscopic | ▪ Autoimmune thyroiditis (Hashimoto thyroiditis).<br>▪ Grave's or Basedow's disease.<br>▪ Diabetes mellitus type 1. |

| | | |
|---|---|---|
| ▪ Autoimmune cholangitis. | colitis.<br>- Ulcerative proctitis.<br>▪ Celiac disease.<br>▪ Autoimmune pancreatitis. | ▪ Addison’s disease.<br>▪ Polyglandular autoimmune syndrome.<br>▪ Autoimmune hypophysitis. |

**Table S3. Solicited local adverse events - number of participants experiencing the events (safety set)**

| | 10µg GBP510 with AS03 (N = 101) | 10µg GBP510 (N = 10) | 25µg GBP510 with AS03 (N = 104) | 25µg GBP510 (N = 51) | Placebo (N = 61) |
|---|---|---|---|---|---|
| Adverse Events after any vaccine injection, n (%), | | | | | |
| Injection Site Pain | 89 (88·12) | 5 (50·00) | 96 (92·31) | 34 (66·67) | 13 (21·31) |
| Injection Site Redness | 8 (7·92) | 0 (0·00) | 11 (10·58) | 0 (0·00) | 0 (0·00) |
| Injection Site Swelling | 11 (10·89) | 0 (0·00) | 14 (13·46) | 2 (3·92) | 0 (0·00) |
| Adverse Events after the 1st vaccination, n (%) | | | | | |
| Injection site pain | 83 (82·18) | 2 (20·00) | 89 (85·58) | 25 (49·02) | 8 (13·11) |
| Injection site redness | 2 (1·98) | 0 (0·00) | 3 (2·88) | 0 (0·00) | 0 (0·00) |
| Injection site swelling | 5 (4·95) | 0 (0·00) | 11 (10.58) | 2 (3·92) | 0 (0·00) |
| Adverse Events after the 2nd vaccination, n (%) | | | | | |
| Injection site pain | 79 (78·22) | 4 (40·00) | 89 (85·58) | 24 (47·06) | 8 (13·11) |
| Injection site redness | 7 (6·93) | 0 (0·00) | 10 (9·62) | 0 (0·00) | 0 (0·00) |
| Injection site swelling | 9 (8·91) | 0 (0·00) | 8 (7·69) | 0 (0·00) | 0 (0·00) |

%: n / (No. of participant in Safety Set by group) * 100
95% Confidence Interval for the percentage of participants with AE is calculated by Clopper-Pearson Methods.
n: number of participants

**Table S4. Solicited systemic adverse events - number of participants experiencing the events (safety set)**

| | 10μg GBP510 with AS03 (N = 101) | 10μg GBP510 (N = 10) | 25μg GBP510 with AS03 (N = 104) | 25μg GBP510 (N = 51) | Placebo (N = 61) |
|---|---|---|---|---|---|
| Adverse Events after any vaccine injection, n (%) | | | | | |
| Fever | 10 (9·90) | 0 (0·00) | 18 (17·31) | 0 (0·00) | 1 (1·64) |
| Nausea/vomiting | 18 (17·82) | 2 (20·00) | 24 (23·08) | 3 (5·88) | 6 (9·84) |
| Diarrhea | 18 (17·82) | 2 (20·00) | 17 (16·35) | 9 (17·65) | 5 (8·20) |
| Headache | 54 (53·47) | 4 (40·00) | 59 (56·73) | 16 (31·37) | 21 (34·43) |
| Fatigue | 80 (79·21) | 4 (40·00) | 78 (75·00) | 31 (60·78) | 30 (49·18) |
| Myalgia | 79 (78·22) | 4 (40·00) | 83 (79·81) | 24 (47·06) | 19 (31·15) |
| Arthralgia | 39 (38·61) | 1 (10·00) | 42 (40·38) | 11 (21·57) | 10 (16·39) |
| Chills | 44 (43·56) | 1 (10·00) | 57 (54·81) | 4 (7·84) | 10 (16·39) |
| Adverse events after the first dose, n (%) | | | | | |
| Fever | 0 (0·00) | 0 (0·00) | 2 (1·92) | 0 (0·00) | 1 (1·64) |
| Nausea/vomiting | 6 (5·94) | 1 (10·00) | 9 (8·65) | 1 (1·96) | 3 (4·92) |
| Diarrhea | 9 (8·91) | 2 (20·00) | 11 (10·58) | 5 (9·80) | 4 (6·56) |
| Headache | 24 (23·76) | 3 (30·00) | 26 (25·00) | 10 (19·61) | 12 (19·67) |
| Fatigue | 56 (55·45) | 3 (30·00) | 54 (51·92) | 24 (47·06) | 25 (40·98) |
| Myalgia | 54 (53·47) | 2 (20·00) | 55 (52·88) | 19 (37·25) | 13 (21·31) |
| Arthralgia | 20 (19·80) | 1 (10·00) | 21 (20·19) | 6 (11·76) | 3 (4·92) |
| Chills | 11 (10·89) | 1 (10·00) | 14 (13·46) | 4 (7·84) | 1 (1·64) |
| Adverse events after the second dose, n (%) | | | | | |
| Fever | 10 (9·90) | 0 (0·00) | 17 (16·35) | 0 (0·00) | 0 (0·00) |
| Nausea/vomiting | 14 (13·86) | 1 (10·00) | 19 (18·27) | 3 (5·88) | 3 (4·92) |
| Diarrhea | 12 (11·88) | 0 (0·00) | 9 (8·65) | 6 (11·76) | 2 (3·28) |
| Headache | 46 (45·54) | 2 (20·00) | 52 (50·00) | 10 (19·61) | 13 (21·31) |
| Fatigue | 69 (68·32) | 2 (20·00) | 77 (74·04) | 22 (43·14) | 24 (39·34) |

| | | | | | |
|---|---|---|---|---|---|
| Myalgia | 68 (67·33) | 3 (30·00) | 76 (73·08) | 15 (29·41) | 11 (18·03) |
| Arthralgia | 33 (32·67) | 0 (0·00) | 36 (34·62) | 5 (9·80) | 8 (13·11) |
| Chills | 41 (40·59) | 0 (0·00) | 53 (50·96) | 1 (1·96) | 10 (16·39) |

%: n / (No. of participant in Safety Set by group) * 100
95% Confidence Interval for the percentage of participants with AE is calculated by Clopper-Pearson Methods.
n: number of participants

**Table S5. Hematologic laboratory monitoring for adverse events through 28 days after second-dose vaccination (safety set)**

| Screening | 10µg GBP510 with AS03 (N = 101) | | 10µg GBP510 (N = 10) | | 25µg GBP510 with AS03 (N = 104) | | 25µg GBP510 (N = 51) | | Placebo (N = 61) | |
|---|---|---|---|---|---|---|---|---|---|---|
| | Normal/NCS | CS | Normal/NCS | CS | Normal/NCS | CS | Normal/NCS | CS | Normal/NCS | CS |
| WBC, n (%) | | | | | | | | | | |
| Normal/NCS | 97 (98·98) | 1 (1·02) | 10 (100·00) | 0 (0·00) | 103 (100·00) | 0 (0·00) | 49 (100·00) | 0 (0·00) | 60 (100·00) | 0 (0·00) |
| CS | 0 (0·00) | 0 (0·00) | 0 (0·00) | 0 (0·00) | 0 (0·00) | 0 (0·00) | 0 (0·00) | 0 (0·00) | 0 (0·00) | 0 (0·00) |
| RBC, n (%) | | | | | | | | | | |
| Normal/NCS | 98 (100·00) | 0 (0·00) | 10 (100·00) | 0 (0·00) | 103 (100·00) | 0 (0·00) | 49 (100·00) | 0 (0·00) | 60 (100·00) | 0 (0·00) |
| CS | 0 (0·00) | 0 (0·00) | 0 (0·00) | 0 (0·00) | 0 (0·00) | 0 (0·00) | 0 (0·00) | 0 (0·00) | 0 (0·00) | 0 (0·00) |
| Hemoglobin, n (%) | | | | | | | | | | |
| Normal/NCS | 98 (100·00) | 0 (0·00) | 10 (100·00) | 0 (0·00) | 103 (100·00) | 0 (0·00) | 49 (100·00) | 0 (0·00) | 60 (100·00) | 0 (0·00) |
| CS | 0 (0·00) | 0 (0·00) | 0 (0·00) | 0 (0·00) | 0 (0·00) | 0 (0·00) | 0 (0·00) | 0 (0·00) | 0 (0·00) | 0 (0·00) |
| Hematocrit, n (%) | | | | | | | | | | |
| Normal/NCS | 98 (100·00) | 0 (0·00) | 10 (100·00) | 0 (0·00) | 103 (100·00) | 0 (0·00) | 49 (100·00) | 0 (0·00) | 60 (100·00) | 0 (0·00) |
| CS | 0 (0·00) | 0 (0·00) | 0 (0·00) | 0 (0·00) | 0 (0·00) | 0 (0·00) | 0 (0·00) | 0 (0·00) | 0 (0·00) | 0 (0·00) |
| Platelet, n (%) | | | | | | | | | | |
| Normal/NCS | 98 (100·00) | 0 (0·00) | 10 (100·00) | 0 (0·00) | 103 (100·00) | 0 (0·00) | 49 (100·00) | 0 (0·00) | 60 (100·00) | 0 (0·00) |
| CS | 0 (0·00) | 0 (0·00) | 0 (0·00) | 0 (0·00) | 0 (0·00) | 0 (0·00) | 0 (0·00) | 0 (0·00) | 0 (0·00) | 0 (0·00) |
| Neutrophil, n (%) | | | | | | | | | | |
| Normal/NCS | 98 (100·00) | 0 (0·00) | 10 (100·00) | 0 (0·00) | 103 (100·00) | 0 (0·00) | 49 (100·00) | 0 (0·00) | 60 (100·00) | 0 (0·00) |
| CS | 0 (0·00) | 0 (0·00) | 0 (0·00) | 0 (0·00) | 0 (0·00) | 0 (0·00) | 0 (0·00) | 0 (0·00) | 0 (0·00) | 0 (0·00) |
| Lymphocytes, n (%) | | | | | | | | | | |
| Normal/NCS | 98 (100·00) | 0 (0·00) | 10 (100·00) | 0 (0·00) | 103 (100·00) | 0 (0·00) | 49 (100·00) | 0 (0·00) | 60 (100·00) | 0 (0·00) |
| CS | 0 (0·00) | 0 (0·00) | 0 (0·00) | 0 (0·00) | 0 (0·00) | 0 (0·00) | 0 (0·00) | 0 (0·00) | 0 (0·00) | 0 (0·00) |
| Eosinophils, n (%) | | | | | | | | | | |
| Normal/NCS | 98 (100·00) | 0 (0·00) | 10 (100·00) | 0 (0·00) | 103 (100·00) | 0 (0·00) | 49 (100·00) | 0 (0·00) | 59 (98·33) | 1 (1·67) |
| CS | 0 (0·00) | 0 (0·00) | 0 (0·00) | 0 (0·00) | 0 (0·00) | 0 (0·00) | 0 (0·00) | 0 (0·00) | 0 (0·00) | 0 (0·00) |

NCS, not clinically significant abnormal; CS, clinically significant abnormal

Participants evaluated both screening and visit 7 were analyzed.

%: n / (No. of participants in each group) * 100; it is calculated based on the total number of participants who were available on visit 7.

n: number of participants

### Table S6. Blood biochemistry test monitoring for adverse events through 28 days after second-dose vaccination (safety set)

| Screening | 10μg GBP510 with AS03 (N = 101) | | 10μg GBP510 (N = 10) | | 25μg GBP510 with AS03 (N = 104) | | 25μg GBP510 (N = 51) | | Placebo (N = 61) | |
|---|---|---|---|---|---|---|---|---|---|---|
| | **Normal/NCS** | **CS** | **Normal/NCS** | **CS** | **Normal/NCS** | **CS** | **Normal/NCS** | **CS** | **Normal/NCS** | **CS** |
| Glucose, n (%) | | | | | | | | | | |
| Normal/NCS | 98 (100·00) | 0 (0·00) | 10 (100·00) | 0 (0·00) | 103 (100·00) | 0 (0·00) | 49 (100·00) | 0 (0·00) | 60 (100·00) | 0 (0·00) |
| CS | 0 (0·00) | 0 (0·00) | 0 (0·00) | 0 (0·00) | 0 (0·00) | 0 (0·00) | 0 (0·00) | 0 (0·00) | 0 (0·00) | 0 (0·00) |
| Aspartate transaminase (AST), n (%) | | | | | | | | | | |
| Normal/NCS | 98 (100·00) | 0 (0·00) | 10 (100·00) | 0 (0·00) | 103 (100·00) | 0 (0·00) | 49 (100·00) | 0 (0·00) | 59 (98·33) | 1 (1·67) |
| CS | 0 (0·00) | 0 (0·00) | 0 (0·00) | 0 (0·00) | 0 (0·00) | 0 (0·00) | 0 (0·00) | 0 (0·00) | 0 (0·00) | 0 (0·00) |
| Alanine transferase (ALT), n (%) | | | | | | | | | | |
| Normal/NCS | 97 (98·98) | 1 (1·02) | 10 (100·00) | 0 (0·00) | 103 (100·00) | 0 (0·00) | 49 (100·00) | 0 (0·00) | 59 (98·33) | 1 (1·67) |
| CS | 0 (0·00) | 0 (0·00) | 0 (0·00) | 0 (0·00) | 0 (0·00) | 0 (0·00) | 0 (0·00) | 0 (0·00) | 0 (0·00) | 0 (0·00) |
| Alkaline phosphatase (ALP), n (%) | | | | | | | | | | |
| Normal/NCS | 98 (100·00) | 0 (0·00) | 10 (100·00) | 0 (0·00) | 103 (100·00) | 0 (0·00) | 49 (100·00) | 0 (0·00) | 60 (100·00) | 0 (0·00) |
| CS | 0 (0·00) | 0 (0·00) | 0 (0·00) | 0 (0·00) | 0 (0·00) | 0 (0·00) | 0 (0·00) | 0 (0·00) | 0 (0·00) | 0 (0·00) |
| Total bilirubin, n (%) | | | | | | | | | | |
| Normal/NCS | 98 (100·00) | 0 (0·00) | 10 (100·00) | 0 (0·00) | 103 (100·00) | 0 (0·00) | 49 (100·00) | 0 (0·00) | 60 (100·00) | 0 (0·00) |
| CS | 0 (0·00) | 0 (0·00) | 0 (0·00) | 0 (0·00) | 0 (0·00) | 0 (0·00) | 0 (0·00) | 0 (0·00) | 0 (0·00) | 0 (0·00) |
| Total protein, n (%) | | | | | | | | | | |
| Normal/NCS | 98 (100·00) | 0 (0·00) | 10 (100·00) | 0 (0·00) | 103 (100·00) | 0 (0·00) | 49 (100·00) | 0 (0·00) | 60 (100·00) | 0 (0·00) |
| CS | 0 (0·00) | 0 (0·00) | 0 (0·00) | 0 (0·00) | 0 (0·00) | 0 (0·00) | 0 (0·00) | 0 (0·00) | 0 (0·00) | 0 (0·00) |
| Albumin, n (%) | | | | | | | | | | |
| Normal/NCS | 98 (100·00) | 0 (0·00) | 10 (100·00) | 0 (0·00) | 103 (100·00) | 0 (0·00) | 49 (100·00) | 0 (0·00) | 60 (100·00) | 0 (0·00) |
| CS | 0 (0·00) | 0 (0·00) | 0 (0·00) | 0 (0·00) | 0 (0·00) | 0 (0·00) | 0 (0·00) | 0 (0·00) | 0 (0·00) | 0 (0·00) |
| BUN, n (%) | | | | | | | | | | |
| Normal/NCS | 98 (100·00) | 0 (0·00) | 10 (100·00) | 0 (0·00) | 103 (100·00) | 0 (0·00) | 49 (100·00) | 0 (0·00) | 60 (100·00) | 0 (0·00) |
| CS | 0 (0·00) | 0 (0·00) | 0 (0·00) | 0 (0·00) | 0 (0·00) | 0 (0·00) | 0 (0·00) | 0 (0·00) | 0 (0·00) | 0 (0·00) |
| Creatinine, n (%) | | | | | | | | | | |
| Normal/NCS | 98 (100·00) | 0 (0·00) | 10 (100·00) | 0 (0·00) | 103 (100·00) | 0 (0·00) | 49 (100·00) | 0 (0·00) | 60 (100·00) | 0 (0·00) |
| CS | 0 (0·00) | 0 (0·00) | 0 (0·00) | 0 (0·00) | 0 (0·00) | 0 (0·00) | 0 (0·00) | 0 (0·00) | 0 (0·00) | 0 (0·00) |
| Uric acid, n (%) | | | | | | | | | | |
| Normal/NCS | 98 (100·00) | 0 (0·00) | 10 (100·00) | 0 (0·00) | 103 (100·00) | 0 (0·00) | 49 (100·00) | 0 (0·00) | 59 (98·33) | 1 (1·67) |

| Screening | 10μg GBP510 with AS03 (N = 101) | | 10μg GBP510 (N = 10) | | 25μg GBP510 with AS03 (N = 104) | | 25μg GBP510 (N = 51) | | Placebo (N = 61) | |
|---|---|---|---|---|---|---|---|---|---|---|
| | Normal/NCS | CS | Normal/NCS | CS | Normal/NCS | CS | Normal/NCS | CS | Normal/NCS | CS |
| CS | 0 (0·00) | 0 (0·00) | 0 (0·00) | 0 (0·00) | 0 (0·00) | 0 (0·00) | 0 (0·00) | 0 (0·00) | 0 (0·00) | 0 (0·00) |
| Total cholesterol, n (%) | | | | | | | | | | |
| Normal/NCS | 98 (100·00) | 0 (0·00) | 10 (100·00) | 0 (0·00) | 103 (100·00) | 0 (0·00) | 48 (97·96) | 1 (2·04) | 60 (100·00) | 0 (0·00) |
| CS | 0 (0·00) | 0 (0·00) | 0 (0·00) | 0 (0·00) | 0 (0·00) | 0 (0·00) | 0 (0·00) | 0 (0·00) | 0 (0·00) | 0 (0·00) |
| Triglyceride, n (%) | | | | | | | | | | |
| Normal/NCS | 95 (96·94) | 0 (0·00) | 10 (100·00) | 0 (0·00) | 100 (97·09) | 3 (2·91) | 49 (100·00) | 0 (0·00) | 59 (98·33) | 1 (1·67) |
| CS | 3 (3·06) | 0 (0·00) | 0 (0·00) | 0 (0·00) | 0 (0·00) | 0 (0·00) | 0 (0·00) | 0 (0·00) | 0 (0·00) | 0 (0·00) |
| Calcium, n (%) | | | | | | | | | | |
| Normal/NCS | 98 (100·00) | 0 (0·00) | 10 (100·00) | 0 (0·00) | 103 (100·00) | 0 (0·00) | 49 (100·00) | 0 (0·00) | 60 (100·00) | 0 (0·00) |
| CS | 0 (0·00) | 0 (0·00) | 0 (0·00) | 0 (0·00) | 0 (0·00) | 0 (0·00) | 0 (0·00) | 0 (0·00) | 0 (0·00) | 0 (0·00) |
| Phosphorus, n (%) | | | | | | | | | | |
| Normal/NCS | 98 (100·00) | 0 (0·00) | 10 (100·00) | 0 (0·00) | 103 (100·00) | 0 (0·00) | 49 (100·00) | 0 (0·00) | 60 (100·00) | 0 (0·00) |
| CS | 0 (0·00) | 0 (0·00) | 0 (0·00) | 0 (0·00) | 0 (0·00) | 0 (0·00) | 0 (0·00) | 0 (0·00) | 0 (0·00) | 0 (0·00) |
| Sodium, n (%) | | | | | | | | | | |
| Normal/NCS | 98 (100·00) | 0 (0·00) | 10 (100·00) | 0 (0·00) | 103 (100·00) | 0 (0·00) | 49 (100·00) | 0 (0·00) | 60 (100·00) | 0 (0·00) |
| CS | 0 (0·00) | 0 (0·00) | 0 (0·00) | 0 (0·00) | 0 (0·00) | 0 (0·00) | 0 (0·00) | 0 (0·00) | 0 (0·00) | 0 (0·00) |
| Potassium, n (%) | | | | | | | | | | |
| Normal/NCS | 98 (100·00) | 0 (0·00) | 10 (100·00) | 0 (0·00) | 103 (100·00) | 0 (0·00) | 49 (100·00) | 0 (0·00) | 60 (100·00) | 0 (0·00) |
| CS | 0 (0·00) | 0 (0·00) | 0 (0·00) | 0 (0·00) | 0 (0·00) | 0 (0·00) | 0 (0·00) | 0 (0·00) | 0 (0·00) | 0 (0·00) |

NCS, not clinically significant abnormal; CS, clinically significant abnormal; BUN, blood urea nitrogen

Participants evaluated at both screening and visit 7 were analyzed.

%: n / (No. of participants in each group) * 100; it is calculated based on the total number of participants who were available on visit 7.

n: number of participants

### Table S7. Unsolicited adverse events by category (safety set)

| | 10μg GBP510 with AS03 (N = 101) | 10μg GBP510 (N = 10) | 25μg GBP510 with AS03 (N = 104) | 25μg GBP510 (N = 51) | Placebo (N = 61) |
|---|---|---|---|---|---|
| Adverse events after any vaccine injection | | | | | |
| Total number of AE occurrences (n) | 26 | 3 | 31 | 16 | 16 |
| Maximum severity, No· of AEs (%) | | | | | |
| Grade 1 (mild) | 21 (80·77) | 1 (33·33) | 18 (58·06) | 13 (81·25) | 14 (87·50) |
| Grade 2 (moderate) | 3 (11·54) | 2 (66·67) | 11 (35·48) | 3 (18·75) | 2 (12·50) |
| Grade 3 (severe) | 2 (7·69) | 0 (0·00) | 2 (6·45) | 0 (0·00) | 0 (0·00) |
| Grade 4 (life threatening) | 0 (0·00) | 0 (0·00) | 0 (0·00) | 0 (0·00) | 0 (0·00) |
| Seriousness, number of AEs (%) | | | | | |
| Yes | 0 (0·00) | 0 (0·00) | 1 (3·23) | 2 (12·50) | 0 (0·00) |
| No | 26 (100·00) | 3 (100·00) | 30 (96·77) | 14 (87·50) | 16 (100·00) |
| Outcome, number of AEs (%) | | | | | |
| Recovered/resolved | 21 (80·77) | 3 (100·00) | 29 (93·55) | 12 (75·00) | 15 (93·75) |
| Recovering/resolving | 3 (11·54) | 0 (0·00) | 1 (3·23) | 2 (12·50) | 1 (6·25) |
| Not recovered/not resolved | 2 (7·69) | 0 (0·00) | 1 (3·23) | 1 (6·25) | 0 (0·00) |
| Stabilized | 1 (3·85) | 0 (0·00) | 1 (3·23) | 1 (6·25) | 0 (0·00) |
| Not stabilized | 1 (3·85) | 0 (0·00) | 0 (0·00) | 0 (0·00) | 0 (0·00) |
| Recovered/resolved with sequelae | 0 (0·00) | 0 (0·00) | 0 (0·00) | 1 (6·25) | 0 (0·00) |
| Fatal | 0 (0·00) | 0 (0·00) | 0 (0·00) | 0 (0·00) | 0 (0·00) |
| Unknown | 0 (0·00) | 0 (0·00) | 0 (0·00) | 0 (0·00) | 0 (0·00) |
| Causality, number of AEs (%) | | | | | |
| Related | 7 (26·92) | 0 (0·00) | 9 (29·03) | 4 (25·00) | 2 (12·50) |
| Not-related | 19 (73·08) | 3 (100·00) | 22 (70·97) | 12 (75·00) | 14 (87·50) |
| Action taken with IP, No· of AEs (%) | | | | | |
| Stop vaccination | 0 (0·00) | 0 (0·00) | 0 (0·00) | 4 (25·00) | 0 (0·00) |
| Continue to vaccination | 12 (46·15) | 2 (66·67) | 17 (54·84) | 7 (43·75) | 7 (43·75) |
| Unknown | 0 (0·00) | 0 (0·00) | 0 (0·00) | 0 (0·00) | 0 (0·00) |
| Not applicable | 14 (53·85) | 1 (33·33) | 14 (45·16) | 5 (31·25) | 9 (56·25) |
| Medication (including other treatment), number of AEs (%) | | | | | |
| Yes | 15 (57·69) | 2 (66·67) | 18 (58·06) | 12 (75·00) | 10 (62·50) |
| No | 11 (42·31) | 1 (33·33) | 13 (41·94) | 4 (25·00) | 6 (37·50) |
| Immediate systemic reaction, number of AEs (%) | | | | | |
| Yes | 0 (0·00) | 0 (0·00) | 0 (0·00) | 0 (0·00) | 0 (0·00) |
| No | 26 (100·00) | 3 (100·00) | 31 (100·00) | 16 (100·00) | 16 (100·00) |

| | 10µg GBP510 with AS03 (N = 101) | 10µg GBP510 (N = 10) | 25µg GBP510 with AS03 (N = 104) | 25µg GBP510 (N = 51) | Placebo (N = 61) |
|---|---|---|---|---|---|
| MAAE, number of AEs (%) | | | | | |
| Yes | 12 (46·15) | 2 (66·67) | 9 (29·03) | 8 (50·00) | 5 (31·25) |
| Hospitalization | 0 (0·00) | 0 (0·00) | 1 (3·23) | 1 (6·25) | 0 (0·00) |
| ER visit | 0 (0·00) | 0 (0·00) | 0 (0·00) | 0 (0·00) | 0 (0·00) |
| Other visit to a healthcare professional | 12 (46·15) | 2 (66·67) | 8 (25·81) | 7 (43·75) | 5 (31·25) |
| No | 14 (53·85) | 1 (33·33) | 22 (70·97) | 8 (50·00) | 11 (68·75) |
| AESI, number of AEs (%) | | | | | |
| Yes | 0 (0·00) | 0 (0·00) | 0 (0·00) | 0 (0·00) | 0 (0·00) |
| No | 26 (100·00) | 3 (100·00) | 31 (100·00) | 16 (100·00) | 16 (100·00) |
| Adverse events after the 1st vaccination | | | | | |
| Total number of AE occurrences (n) | 12 | 2 | 17 | 11 | 7 |
| Maximum severity, number of AEs (%) | | | | | |
| Grade 1 (mild) | 11 (91·67) | 1 (50·00) | 9 (52·94) | 9 (81·82) | 6 (85·71) |
| Grade 2 (moderate) | 0 (0·00) | 1 (50·00) | 6 (35·29) | 2 (18·18) | 1 (14·29) |
| Grade 3 (severe) | 1 (8·33) | 0 (0·00) | 2 (11·76) | 0 (0·00) | 0 (0·00) |
| Grade 4 (life threatening) | 0 (0·00) | 0 (0·00) | 0 (0·00) | 0 (0·00) | 0 (0·00) |
| Seriousness, number of AEs (%) | | | | | |
| Yes | 0 (0·00) | 0 (0·00) | 1 (5·88) | 2 (18·18) | 0 (0·00) |
| No | 12 (100·00) | 2 (100·00) | 16 (94·12) | 9 (81·82) | 7 (100·00) |
| Outcome, number of AEs (%) | | | | | |
| Recovered/resolved | 9 (75·00) | 2 (100·00) | 17 (100·00) | 7 (63·64) | 7 (100·00) |
| Recovering/resolving | 2 (16·67) | 0 (0·00) | 0 (0·00) | 2 (18·18) | 0 (0·00) |
| Not recovered/not resolved | 1 (8·33) | 0 (0·00) | 0 (0·00) | 1 (9·09) | 0 (0·00) |
| Stabilized | 0 (0·00) | 0 (0·00) | 0 (0·00) | 1 (9·09) | 0 (0·00) |
| Not stabilized | 1 (8·33) | 0 (0·00) | 0 (0·00) | 0 (0·00) | 0 (0·00) |
| Recovered/resolved with sequelae | 0 (0·00) | 0 (0·00) | 0 (0·00) | 1 (9·09) | 0 (0·00) |
| Fatal | 0 (0·00) | 0 (0·00) | 0 (0·00) | 0 (0·00) | 0 (0·00) |
| Unknown | 0 (0·00) | 0 (0·00) | 0 (0·00) | 0 (0·00) | 0 (0·00) |
| Causality, number of AEs (%) | | | | | |
| Related | 1 (8·33) | 0 (0·00) | 5 (29·41) | 4 (36·36) | 1 (14·29) |
| Not-related | 11 (91·67) | 2 (100·00) | 12 (70·59) | 7 (63·64) | 6 (85·71) |
| Action taken with IP, number of AEs (%) | | | | | |
| Stop vaccination | 0 (0·00) | 0 (0·00) | 0 (0·00) | 4 (36·36) | 0 (0·00) |
| Continue to vaccination | 12 (100·00) | 2 (100·00) | 17 (100·00) | 7 (63·64) | 7 (100·00) |
| Unknown | 0 (0·00) | 0 (0·00) | 0 (0·00) | 0 (0·00) | 0 (0·00) |

| | 10µg GBP510 with AS03 (N = 101) | 10µg GBP510 (N = 10) | 25µg GBP510 with AS03 (N = 104) | 25µg GBP510 (N = 51) | Placebo (N = 61) |
|---|---|---|---|---|---|
| Not applicable | 0 (0·00) | 0 (0·00) | 0 (0·00) | 0 (0·00) | 0 (0·00) |
| Medication (including other treatment), number of AEs (%) | | | | | |
| Yes | 9 (75·00) | 1 (50·00) | 11 (64·71) | 10 (90·91) | 7 (100·00) |
| No | 3 (25·00) | 1 (50·00) | 6 (35·29) | 1 (9·09) | 0 (0·00) |
| Immediate systemic reaction, number of AEs (%) | | | | | |
| Yes | 0 (0·00) | 0 (0·00) | 0 (0·00) | 0 (0·00) | 0 (0·00) |
| No | 12 (100·00) | 2 (100·00) | 17 (100·00) | 11 (100·00) | 7 (100·00) |
| MAAE, number of AEs (%) | | | | | |
| Yes | 5 (41·67) | 2 (100·00) | 6 (35·29) | 7 (63·64) | 2 (28·57) |
| Hospitalization | 0 (0·00) | 0 (0·00) | 1 (5·88) | 1 (9·09) | 0 (0·00) |
| ER visit | 0 (0·00) | 0 (0·00) | 0 (0·00) | 0 (0·00) | 0 (0·00) |
| Other visit to a healthcare professional | 5 (41·67) | 2 (100·00) | 5 (29·41) | 6 (54·55) | 2 (28·57) |
| No | 7 (58·33) | 0 (0·00) | 11 (64·71) | 4 (36·36) | 5 (71·43) |
| AESI, number of AEs (%) | | | | | |
| Yes | 0 (0·00) | 0 (0·00) | 0 (0·00) | 0 (0·00) | 0 (0·00) |
| No | 12 (100·00) | 2 (100·00) | 17 (100·00) | 11 (100·00) | 7 (100·00) |
| Adverse events after the 2nd vaccination | | | | | |
| Total number of AE occurrences (n) | 14 | 1 | 14 | 5 | 9 |
| Maximum severity, number of AEs (%) | | | | | |
| Grade 1 (mild) | 10 (71·43) | 0 (0·00) | 9 (64·29) | 4 (80·00) | 8 (88·89) |
| Grade 2 (moderate) | 3 (21·43) | 1 (100·00) | 5 (35·71) | 1 (20·00) | 1 (11·11) |
| Grade 3 (severe) | 1 (7·14) | 0 (0·00) | 0 (0·00) | 0 (0·00) | 0 (0·00) |
| Grade 4 (life threatening) | 0 (0·00) | 0 (0·00) | 0 (0·00) | 0 (0·00) | 0 (0·00) |
| Seriousness, number of AEs (%) | | | | | |
| Yes | 0 (0·00) | 0 (0·00) | 0 (0·00) | 0 (0·00) | 0 (0·00) |
| No | 14 (100·00) | 1 (100·00) | 14 (100·00) | 5 (100·00) | 9 (100·00) |
| Outcome, number of AEs (%) | | | | | |
| Recovered/resolved | 12 (85·71) | 1 (100·00) | 12 (85·71) | 5 (100·00) | 8 (88·89) |
| Recovering/resolving | 1 (7·14) | 0 (0·00) | 1 (7·14) | 0 (0·00) | 1 (11·11) |
| Not recovered/not resolved | 1 (7·14) | 0 (0·00) | 1 (7·14) | 0 (0·00) | 0 (0·00) |
| Stabilized | 1 (7·14) | 0 (0·00) | 1 (7·14) | 0 (0·00) | 0 (0·00) |
| Not stabilized | 0 (0·00) | 0 (0·00) | 0 (0·00) | 0 (0·00) | 0 (0·00) |
| Recovered/resolved with sequelae | 0 (0·00) | 0 (0·00) | 0 (0·00) | 0 (0·00) | 0 (0·00) |
| Fatal | 0 (0·00) | 0 (0·00) | 0 (0·00) | 0 (0·00) | 0 (0·00) |
| Unknown | 0 (0·00) | 0 (0·00) | 0 (0·00) | 0 (0·00) | 0 (0·00) |

| | 10µg GBP510 with AS03 (N = 101) | 10µg GBP510 (N = 10) | 25µg GBP510 with AS03 (N = 104) | 25µg GBP510 (N = 51) | Placebo (N = 61) |
|---|---|---|---|---|---|
| Causality, number of AEs (%) | | | | | |
| Related | 6 (42·86) | 0 (0·00) | 4 (28·57) | 0 (0·00) | 1 (11·11) |
| Not-related | 8 (57·14) | 1 (100·00) | 10 (71·43) | 5 (100·00) | 8 (88·89) |
| Action taken with IP, number of AEs (%) | | | | | |
| Stop vaccination | 0 (0·00) | 0 (0·00) | 0 (0·00) | 0 (0·00) | 0 (0·00) |
| Continue to vaccination | 0 (0·00) | 0 (0·00) | 0 (0·00) | 0 (0·00) | 0 (0·00) |
| Unknown | 0 (0·00) | 0 (0·00) | 0 (0·00) | 0 (0·00) | 0 (0·00) |
| Not applicable | 14 (100·00) | 1 (100·00) | 14 (100·00) | 5 (100·00) | 9 (100·00) |
| Medication (including other treatment), number of AEs (%) | | | | | |
| Yes | 6 (42·86) | 1 (100·00) | 7 (50·00) | 2 (40·00) | 3 (33·33) |
| No | 8 (57·14) | 0 (0·00) | 7 (50·00) | 3 (60·00) | 6 (66·67) |
| Immediate systemic reaction, number of AEs (%) | | | | | |
| Yes | 0 (0·00) | 0 (0·00) | 0 (0·00) | 0 (0·00) | 0 (0·00) |
| No | 14 (100·00) | 1 (100·00) | 14 (100·00) | 5 (100·00) | 9 (100·00) |
| MAAE, number of AEs (%) | | | | | |
| Yes | 7 (50·00) | 0 (0·00) | 3 (21·43) | 1 (20·00) | 3 (33·33) |
| Hospitalization | 0 (0·00) | 0 (0·00) | 0 (0·00) | 0 (0·00) | 0 (0·00) |
| ER visit | 0 (0·00) | 0 (0·00) | 0 (0·00) | 0 (0·00) | 0 (0·00) |
| Other visit to a healthcare professional | 7 (50·00) | 0 (0·00) | 3 (21·43) | 1 (20·00) | 3 (33·33) |
| No | 7 (50·00) | 1 (100·00) | 11 (78·57) | 4 (80·00) | 6 (66·67) |
| AESI, number of AEs (%) | | | | | |
| Yes | 0 (0·00) | 0 (0·00) | 0 (0·00) | 0 (0·00) | 0 (0·00) |
| No | 14 (100·00) | 1 (100·00) | 14 (100·00) | 5 (100·00) | 9 (100·00) |

AEs, adverse events; SAE, serious adverse events; MAAE, medically attended adverse events; adverse events of special interest (AESIs); MAADRS, medically attended adverse drug reactions; IP, investigational product; ER, emergency room.
%: n / (No. of AEs in Safety Set by group) * 100
n: number of AE

**Table S8. Geometric mean concentrations and seroconversion rates of IgG antibody titer to the SARS-CoV-2-RBD by enzyme-linked immunosorbent assay (per-protocol set)**

| | 10μg GBP510 with AS03 (N = 93) | 10μg GBP510 (N = 10) | 25μg GBP510 with AS03 (N = 96) | 25μg GBP510 (N = 45) | Placebo (N = 58) |
|---|---|---|---|---|---|
| Geometric mean titer and seroconversion rate | | | | | |
| Baseline | | | | | |
| n | 93 | 10 | 96 | 45 | 58 |
| GMC ± standard deviation (BAU/mL) | 15·08 ± 1·69 | 10·95 ± 1·61 | 13·85 ± 1·72 | 16·34 ± 1·66 | 12·51 ± 1·64 |
| 95% CI (lower, upper) | (13·54, 16·80) | (7·78, 15·40) | (12·42, 15·46) | (14·04, 19·03) | (10·97, 14·25) |
| P-value vs. placebo | 0·0311 | 0·4339 | 0·2422 | 0·0084 | |
| P-value vs. GBP510 10μg with AS03 | | 0·0671 | 0·2750 | | |
| P-value vs. GBP510 25μg with AS03 | | | | 0·0862 | |
| Visit 4 (4 weeks post-1st vaccination) | | | | | |
| n | 93 | 10 | 96 | 45 | 58 |
| GMC ± standard deviation (BAU/mL) | 111·85 ± 2·10 | 37·72 ± 2·03 | 129·00 ± 2·09 | 28·63 ± 1·72 | 12·71 ± 1·71 |
| 95% CI (lower, upper) | (96·00, 130·31) | (22·74, 62·56) | (111·05, 149·84) | (24·32, 33·71) | (11·04, 14·63) |
| P-value vs. placebo | <0·0001 | <0·0001 | <0·0001 | <0·0001 | |
| P-value vs. GBP510 10μg with AS03 | | <0·0001 | 0·1871 | | |
| P-value vs. GBP510 25μg with AS03 | | | | <0·0001 | |
| GMFR ± standard deviation | 7·42 ± 2·48 | 3·45 ± 1·91 | 9·31 ± 2·34 | 1·75 ± 1·94 | 1·02 ± 1·90 |
| 95% CI (lower, upper) | (6·15, 8·94) | (2·17, 5·47) | (7·84, 11·06) | (1·44, 2·14) | (0·86, 1·20) |
| P-value vs. placebo | <0·0001 | <0·0001 | <0·0001 | <0·0001 | |
| P-value vs. GBP510 10μg with AS03 | | 0·0110 | 0·0774 | | |
| P-value vs. GBP510 25μg with AS03 | | | | <0·0001 | |
| SCR, n (%) | 68 (73·12) | 4 (40·00) | 79 (82·29) | 5 (11·11) | 0 (0·00) |
| 95% CI (lower, upper) | (62·92, 81·79) | (12·16, 73·76) | (73·17, 89·33) | (3·71, 24·05) | (0·00, 6·16) |
| P-value vs. placebo | <0·0001 | 0·0003 | <0·0001 | 0·0140 | |
| P-value vs. GBP510 10μg with AS03 | | 0·0624 | 0·1294 | | |
| P-value vs. GBP510 25μg with AS03 | | | | <0·0001 | |
| Visit 6 (2 weeks post-2nd vaccination) | | | | | |
| n | 93 | 10 | 96 | 45 | 58 |
| GMC ± standard deviation (BAU/mL) | 2,163·59 ± 1·89 | 155·30 ± 3·05 | 2,599·22 ± 1·86 | 112·40 ± 2·75 | 10·56 ± 1·63 |
| 95% CI (lower, upper) | (1,898·11, 2,466·20) | (70·02, 344·47) | (2,292·62, 2,946·83) | (82·92, 152·37) | (9·28, 12·01) |

| | 10µg GBP510 with AS03 (N = 93) | 10µg GBP510 (N = 10) | 25µg GBP510 with AS03 (N = 96) | 25µg GBP510 (N = 45) | Placebo (N = 58) |
|---|---|---|---|---|---|
| P-value vs. placebo | <0·0001 | <0·0001 | <0·0001 | <0·0001 | |
| P-value vs. GBP510 10µg with AS03 | | <0·0001 | 0·0459 | | |
| P-value vs. GBP510 25µg with AS03 | | | | <0·0001 | |
| GMFR ± standard deviation | 143·47 ± 2·33 | 14·19 ± 2·58 | 187·61 ± 2·09 | 6·88 ± 2·93 | 0·84 ± 1·66 |
| 95% CI (lower, upper) | (120·48, 170·85) | (7·21, 27·92) | (161·61, 217·81) | (4·98, 9·49) | (0·74, 0·96) |
| P-value vs. Placebo | <0·0001 | <0·0001 | <0·0001 | <0·0001 | |
| P-value vs. GBP510 10µg with AS03 | | <0·0001 | 0·0212 | | |
| P-value vs. GBP510 25µg with AS03 | | | | <0·0001 | |
| SCR, n (%) | 93 (100·00) | 10 (100·00) | 96 (100·00) | 29 (64·44) | 0 (0·00) |
| 95% CI (lower, upper) | (96·11, 100·00) | (69·15, 100·00) | (96·23, 100·00) | (48·78, 78·13) | (0·00, 6·16) |
| P-value vs. placebo | <0·0001 | <0·0001 | <0·0001 | <0·0001 | |
| P-value vs. GBP510 10µg with AS03 | | NA | NA | | |
| P-value vs. GBP510 25µg with AS03 | | | | <0·0001 | |
| Visit 7 (4 weeks post-2nd vaccination) | | | | | |
| n | 93 | 10 | 96 | 45 | 58 |
| GMC ± standard deviation (BAU/mL) | 1,517·87 ± 2·09 | 132·64 ± 2·76 | 1,943·44 ± 1·79 | 114·97 ± 2·74 | 12·52 ± 1·71 |
| 95% CI (lower, upper) | (1,304·70, 1,765·87) | (64·23, 273·91) | (1,727·73, 2,186·08) | (84·90, 155·68) | (10·88, 14·42) |
| P-value vs. placebo | <0·0001 | <0·0001 | <0·0001 | <0·0001 | |
| P-value vs. GBP510 10µg with AS03 | | <0·0001 | 0·0113 | | |
| P-value vs. GBP510 25µg with AS03 | | | | <0·0001 | |
| GMFR ± standard deviation | 100·65 ± 2·55 | 12·12 ± 2·37 | 140·28 ± 2·06 | 7·03 ± 2·90 | 1·00 ± 1·93 |
| 95% CI (lower, upper) | (83·03, 122·01) | (6·54, 22·47) | (121·14, 162·44) | (5·11, 9·68) | (0·84, 1·19) |
| P-value vs. placebo | <0·0001 | <0·0001 | <0·0001 | <0·0001 | |
| P-value vs. GBP510 10µg with AS03 | | <0·0001 | 0·0071 | | |
| P-value vs. GBP510 25µg with AS03 | | | | <0·0001 | |
| SCR, n (%) | 92 (98·92) | 10 (100·00) | 96 (100·00) | 28 (62·22) | 1 (1·72) |
| 95% CI (lower, upper) | (94·15, 99·97) | (69·15, 100·00) | (96·23, 100·00) | (46·54, 76·23) | (0·04, 9·24) |
| P-value vs. placebo | <0·0001 | <0·0001 | <0·0001 | <0·0001 | |
| P-value vs. GBP510 10µg with AS03 | | 1·0000 | 0·4921 | | |
| P-value vs. GBP510 25µg with AS03 | | | | <0·0001 | |

GMC, geometric mean concentrations; GMFR, geometric mean fold rise; SCR, seroconversion rate; CI, confidence interval; BAU, binding antibody unit; NA, not applicable

GMFR = GMC (each visit) / GMC (Baseline)

SCR: Percentage of participants with ≥ 4-fold rise from baseline

%: n / (No. of subject who information was collected at each visit by group) * 100

The 95% CI for GMC/GMFR is calculated based on the t-distribution of the log-transformed values for geometric means or geometric mean fold rises, then back transformed to the original scale for presentation.

The 95% CI for SCR is calculated by Clopper-Pearson Methods.

P-value versus placebo: p-value for difference of Treatment groups versus Placebo (continuous values are two sample t-test and categorical values are chi-square test or fisher's exact test)

P-value versus GBP510 10µg with AS03: p-value for difference of each group versus GBP510 10µg with AS03 (continuous values are two sample t-test and categorical values are chi-square test or fisher's exact test)

P-value versus GBP510 25µg with AS03: p-value for difference of each group versus GBP510 25µg with AS03 (continuous values are two sample t-test and categorical values are chi-square test or fisher's exact test)

n: number of participants

**Table S9. Geometric mean titers and seroconversion rates of neutralizing antibody to the SARS-CoV-2 by pseudovirus-based neutralization assay (per-protocol set)**

| | 10μg GBP510 with AS03 (N = 93) | 10μg GBP510 (N = 10) | 25μg GBP510 with AS03 (N = 96) | 25μg GBP510 (N = 45) | Placebo (N = 58) |
|---|---|---|---|---|---|
| Geometric mean titer and seroconversion rate | | | | | |
| Baseline | | | | | |
| n | 93 | 10 | 96 | 45 | 58 |
| GMT ± standard deviation (IU/mL) | 18·05 ± 1·28 | 21·01 ± 1·58 | 19·49 ± 1·43 | 19·05 ± 1·48 | 19·19 ± 1·51 |
| 95% CI (lower, upper) | (17·16, 18·99) | (15·16, 29·12) | (18·13, 20·95) | (16·94, 21·43) | (17·21, 21·39) |
| P-value vs. placebo | 0·3126 | 0·5302 | 0·8043 | 0·9290 | |
| P-value vs. GBP510 10μg with AS03 | | 0·3266 | 0·0869 | | |
| P-value vs. GBP510 25μg with AS03 | | | | 0·7321 | |
| Visit 4 (4 weeks post-1st vaccination) | | | | | |
| n | 93 | 10 | 96 | 45 | 58 |
| GMT± standard deviation (IU/mL) | 39·97 ± 2·73 | 32·79 ± 2·45 | 40·94 ± 2·77 | 21·23 ± 1·65 | 19·71 ± 1·58 |
| 95% CI (lower, upper) | (32·51, 49·14) | (17·28, 62·23) | (33·31, 50·32) | (18·28, 24·65) | (17·48, 22·22) |
| P-value vs. placebo | <0·0001 | 0·1095 | <0·0001 | 0·4327 | |
| P-value vs. GBP510 10μg with AS03 | | 0·5509 | 0·8705 | | |
| P-value vs. GBP510 25μg with AS03 | | | | <0·0001 | |
| GMFR ± standard deviation | 2·21 ± 2·78 | 1·56 ± 2·42 | 2·10 ± 2·92 | 1·11 ± 1·63 | 1·03 ± 1·91 |
| 95% CI (lower, upper) | (1·79, 2·73) | (0·83, 2·94) | (1·69, 2·61) | (0·96, 1·29) | (0·87, 1·22) |
| P-value vs. placebo | <0·0001 | 0·0776 | <0·0001 | 0·4822 | |
| P-value vs. GBP510 10μg with AS03 | | 0·3011 | 0·7307 | | |
| P-value vs. GBP510 25μg with AS03 | | | | <0·0001 | |
| SCR, n (%) | 28 (30·11) | 1 (10·00) | 27 (28·13) | 2 (4·44) | 4 (6·90) |
| 95% CI (lower, upper) | (21·03, 40·50) | (0·25, 44·50) | (19·42, 38·22) | (0·54, 15·15) | (1·91, 16·73) |
| P-value vs. placebo | 0·0007 | 0·5604 | 0·0015 | 0·6938 | |
| P-value vs. GBP510 10μg with AS03 | | 0·2754 | 0·7642 | | |
| P-value vs. GBP510 25μg with AS03 | | | | 0·0012 | |
| Visit 6 (2 weeks post-2nd vaccination) | | | | | |
| n | 93 | 10 | 96 | 45 | 58 |
| GMT ± standard deviation (IU/mL) | 1,369·02 ± 2·60 | 83·47 ± 4·47 | 1,431·45 ± 2·69 | 63·85 ± 3·31 | 19·24 ± 1·47 |
| 95% CI (lower, upper) | (1,124·49, 1,666·73) | (28·62, 243·48) | (1,171·07, 1,749·73) | (44·55, 91·50) | (17·37, 21·31) |

| | 10µg GBP510 with AS03 (N = 93) | 10µg GBP510 (N = 10) | 25µg GBP510 with AS03 (N = 96) | 25µg GBP510 (N = 45) | Placebo (N = 58) |
|---|---|---|---|---|---|
| P-value vs. placebo | <0·0001 | 0·0127 | <0·0001 | <0·0001 | |
| P-value vs. GBP510 10µg with AS03 | | 0·0002 | 0·7533 | | |
| P-value vs. GBP510 25µg with AS03 | | | | <0·0001 | |
| GMFR ± Standard deviation | 75·83 ± 2·68 | 3·97 ± 4·09 | 73·44 ± 2·88 | 3·35 ± 3·44 | 1·00 ± 1·64 |
| 95% CI (lower, upper) | (61·91, 92·88) | (1·45, 10·88) | (59·30, 90·97) | (2·31, 4·86) | (0·88, 1·14) |
| P-value vs. placebo | <0·0001 | 0·0129 | <0·0001 | <0·0001 | |
| P-value vs. GBP510 10µg with AS03 | | <0·0001 | 0·8300 | | |
| P-value vs. GBP510 25µg with AS03 | | | | <0·0001 | |
| SCR, n (%) | 93 (100·00) | 5 (50·00) | 95 (98·96) | 23 (51·11) | 1 (1·72) |
| 95% CI (lower, upper) | (96·11, 100·00) | (18·71, 81·29) | (94·33, 99·97) | (35·77, 66·30) | (0·04, 9·24) |
| P-value vs. placebo | <0·0001 | 0·0001 | <0·0001 | <0·0001 | |
| P-value vs. GBP510 10µg with AS03 | | <0·0001 | 1·0000 | | |
| P-value vs. GBP510 25µg with AS03 | | | | <0·0001 | |
| Visit 7 (4 weeks post-2nd vaccination) | | | | | |
| n | 93 | 10 | 96 | 45 | 58 |
| GMT ± standard deviation (IU/mL) | 838·79 ± 2·74 | 68·52 ± 3·85 | 1,107·32 ± 2·61 | 51·87 ± 3·36 | 19·29 ± 1·49 |
| 95% CI (lower, upper) | (681·68, 1,032·11) | (26·12, 179·79) | (911·43, 1,345·31) | (36·05, 74·63) | (17·38, 21·41) |
| P-value vs. placebo | <0·0001 | 0·0157 | <0·0001 | <0·0001 | |
| P-value vs. GBP510 10µg with AS03 | | <0·0001 | 0·0539 | | |
| P-value vs. GBP510 25µg with AS03 | | | | <0·0001 | |
| GMFR ± Standard deviation | 46·46 ± 2·78 | 3·26 ± 4·15 | 56·81 ± 2·95 | 2·72 ± 3·72 | 1·01 ± 1·72 |
| 95% CI (lower, upper) | (37·62, 57·37) | (1·18, 9·02) | (45·64, 70·72) | (1·84, 4·04) | (0·87, 1·16) |
| P-value vs. placebo | <0·0001 | 0·0284 | <0·0001 | <0·0001 | |
| P-value vs. GBP510 10µg with AS03 | | <0·0001 | 0·1909 | | |
| P-value vs. GBP510 25µg with AS03 | | | | <0·0001 | |
| SCR, n (%) | 91 (97·85) | 5 (50·00) | 95 (98·96) | 19 (42·22) | 2 (3·45) |
| 95% CI (lower, upper) | (92·45, 99·74) | (18·71, 81·29) | (94·33, 99·97) | (27·66, 57·85) | (0·42, 11·91) |
| P-value vs. placebo | <0·0001 | 0·0004 | <0·0001 | <0·0001 | |
| P-value vs. GBP510 10µg with AS03 | | <0·0001 | 0·6171 | | |
| P-value vs. GBP510 25µg with AS03 | | | | <0·0001 | |

GMT, geometric mean titer; GMFR, geometric mean fold rise; SCR, seroconversion rate; CI, confidence interval
GMFR = GMT (each visit) / GMT (Baseline)
SCR: Percentage of participants with ≥ 4-fold rise from baseline
%: n / (No· of participant who information was collected at each visit by group) * 100
The 95% CI for GMT/GMFR is calculated based on the t-distribution of the log-transformed values for geometric means or geometric mean fold rises, then back transformed to the original scale for presentation.
The 95% CI for SCR is calculated by Clopper-Pearson Methods.
P-value versus placebo: p-value for difference of Treatment groups versus Placebo (continuous values are two sample t-test and categorical values are chi-square test or fisher's exact test)
P-value versus GBP510 10µg with AS03: p-value for difference of each group versus GBP510 10µg with AS03 (continuous values are two sample t-test and categorical values are chi-square test or fisher's exact test)
P-value versus GBP510 25µg with AS03: p-value for difference of each group versus GBP510 25µg with AS03 (continuous values are two sample t-test and categorical values are chi-square test or fisher's exact test)

**Table S10. Geometric mean titers and seroconversion rates of neutralizing antibody to the SARS-CoV-2 by plaque reduction neutralization test (per-protocol set)**

| | 10µg GBP510 with AS03 (N = 93) | 10µg GBP510 (N = 10) | 25µg GBP510 with AS03 (N = 96) | 25µg GBP510 (N = 45) | Placebo (N = 58) |
|---|---|---|---|---|---|
| Geometric mean titer and seroconversion rate | | | | | |
| Baseline | | | | | |
| N | 23 | 4 | 21 | 9 | 19 |
| GMT ± Standard deviation (IU/mL) | 4·40±1·71 | 9·00±1·00 | 4·33±1·70 | 4·33±1·73 | 4·76±1·75 |
| 95% CI (lower, upper) | (3·49, 5·54) | (9·00, 9·00) | (3·40, 5·51) | (2·84, 6·60) | (3·64, 6·23) |
| P-value vs. Placebo | 0·6367 | <0·0001 | 0·5787 | 0·6713 | |
| P-value vs. GBP510 10µg with AS03 | | <0·0001 | 0·9216 | | |
| P-value vs. GBP510 25µg with AS03 | | | | 1·0000 | |
| Visit 6 (2 weeks post-2nd vaccination) | | | | | |
| N | 23 | 4 | 21 | 9 | 19 |
| GMT ± Standard deviation (IU/mL) | 949·84±2·24 | 34·08±6·05 | 860·96±1·86 | 58·13±5·09 | 4·76±1·75 |
| 95% CI (lower, upper) | (670·40, 1,345·77) | (1·94, 597·40) | (649·60, 1,141·10) | (16·64, 203·13) | (3·64, 6·23) |
| P-value vs. Placebo | <0·0001 | 0·1156 | <0·0001 | 0·0016 | |
| P-value vs. GBP510 10µg with AS03 | | 0·0320 | 0·6548 | | |
| P-value vs. GBP510 25µg with AS03 | | | | 0·0009 | |
| GMFR ± Standard deviation | 216·06±2·04 | 3·79±6·05 | 198·99±1·83 | 13·44±4·50 | 1·00±1·00 |
| 95% CI (lower, upper) | (158·81, 293·95) | (0·22, 66·38) | (151·17, 261·93) | (4·23, 42·71) | (1·00, 1·00) |
| P-value vs. Placebo | <0·0001 | 0·2356 | <0·0001 | 0·0008 | |
| P-value vs. GBP510 10µg with AS03 | | 0·0190 | 0·6827 | | |
| P-value vs. GBP510 25µg with AS03 | | | | 0·0005 | |
| SCR, n (%) | 23 (100·00) | 2 (50·00) | 21 (100·00) | 7 (77·78) | 0 (0·00) |
| 95% CI (lower, upper) | (85·18, 100·00) | (6·76, 93·24) | (83·89, 100·00) | (39·99, 97·19) | (0·00, 17·65) |
| P-value vs. Placebo | <0·0001 | 0·0237 | <0·0001 | <0·0001 | |
| P-value vs. GBP510 10µg with AS03 | | 0·0171 | NA | | |
| P-value vs. GBP510 25µg with AS03 | | | | 0·0828 | |

GMT, geometric mean titer; GMFR, geomtetric mean fold rise; SCR, seroconversion rate; CI, confidence interval
GMFR = GMT (each visit) / GMT (Baseline)
SCR: Percentage of participants with ≥ 4-fold rise from baseline
%: n / (No. of subject who information was collected at each visit by group) * 100

The 95% CI for GMT/GMFR is calculated based on the t-distribution of the log-transformed values for geometric means or geometric mean fold rises, then back transformed to the original scale for presentation·
The 95% CI for SCR is calculated by Clopper-Pearson Methods·
P-value vs Placebo: P-value for difference of Treatment groups versus Placebo (continuous values are two sample t-test and categorical values are chi-square test or fisher's exact test·)
P-value vs GBP510 10μg with AS03: P-value for difference of each group versus GBP510 10μg with AS03 (continuous values are two sample t-test and categorical values are chi-square test or fisher's exact test·)
P-value vs GBP510 25μg with AS03: P-value for difference of each group versus GBP510 25μg with AS03 (continuous values are two sample t-test and categorical values are chi-square test or fisher's exact test·)
n: number of participants

**Table S11. Geometric mean concentration and seroconversion rate of IgG antibody to the SARS-CoV-2-RBD in adults aged 19-64 years: analysis by enzyme-linked immunosorbent assay (per-protocol set)**

| | **10μg GBP510 with AS03 (N=81)** | **10μg GBP510 (N=10)** | **25μg GBP510 with AS03 (N=85)** | **25μg GBP510 (N=41)** | **Placebo (N=53)** |
|---|---|---|---|---|---|
| Geometric mean titer and seroconversion rate | | | | | |
| Baseline | | | | | |
| n | 81 | 10 | 85 | 41 | 53 |
| GMC±standard deviation (BAU/mL) | 14·72±1·72 | 10·95±1·61 | 13·18±1·67 | 16·24±1·70 | 12·20±1·63 |
| 95% CI (lower, upper) | (13·06, 16·59) | (7·78, 15·40) | (11·81, 14·72) | (13·4, 19·20) | (10·66, 13·97) |
| P-value vs. Placebo | 0·0441 | 0·5220 | 0·3820 | 0·0081 | |
| P-value vs. 10μg GBP510 with AS03 | | 0·1020 | 0·1799 | | |
| P-value vs. 25μg GBP510 with AS03 | | | | 0·0360 | |
| Visit 4 (4 weeks post-1st vaccination) | | | | | |
| n | 81 | 10 | 85 | 41 | 53 |
| GMC±standard deviation (BAU/mL) | 121·33±1·94 | 37·72±2·03 | 135·87±2·03 | 29·65±1·72 | 12·66±1·71 |
| 95% CI (lower, upper) | (104·75, 140·53) | (22·74, 62·56) | (116·57, 158·37) | (24·97, 35·20) | (10·92, 14·67) |
| P-value vs. Placebo | <0·0001 | <0·0001 | <0·0001 | <0·0001 | |
| P-value vs. 10μg GBP510 with AS03 | | <0·0001 | 0·2912 | | |
| P-value vs. 25μg GBP510 with AS03 | | | | <0·0001 | |
| GMFR±standard deviation | 8·24±2·33 | 3·45±1·91 | 10·31±2·23 | 1·83±1·94 | 1·04±1·88 |
| 95% CI (lower, upper) | (6·84, 9·94) | (2·17, 5·47) | (8·67, 12·26) | (1·48, 2·25) | (0·87, 1·23) |
| P-value vs. Placebo | <0·0001 | <0·0001 | <0·0001 | <0·0001 | |
| P-value vs. 10μg GBP510 with AS03 | | 0·0023 | 0·0835 | | |
| P-value vs. 25μg GBP510 with AS03 | | | | <0·0001 | |
| SCR, n (%) | 63 (77·78) | 4 (40·00) | 75 (88·24) | 5 (12·20) | 0 (0·00) |
| 95% CI (lower, upper) | (67·17, 86·27) | (12·16, 73·76) | (79·43, 94·21) | (4·08, 26·20) | (0·00, 6·72) |
| P-value vs. Placebo | <0·0001 | 0·0004 | <0·0001 | 0·0137 | |
| P-value vs. 10μg GBP510 with AS03 | | 0·0189 | 0·0721 | | |
| P-value vs 25μg GBP510 with AS03 | | | | <0·0001 | |

| | 10μg GBP510 with AS03 (N=81) | 10μg GBP510 (N=10) | 25μg GBP510 with AS03 (N=85) | 25μg GBP510 (N=41) | Placebo (N=53) |
|---|---|---|---|---|---|
| Visit 6 (2 weeks post-2nd vaccination) | | | | | |
| n | 81 | 10 | 85 | 41 | 53 |
| GMC±standard deviation (BAU/mL) | 2,368·72±1·72 | 155·30±3·05 | 2,777·08±1·71 | 114·64±2·69 | 10·37±1·62 |
| 95% CI (lower, upper) | (2,100·72, 2,670·91) | (70·02, 344·47) | (2,475·11, 3,115·90) | (83·84, 156·75) | (9·07, 11·85) |
| P-value vs. Placebo | <0·0001 | <0·0001 | <0·0001 | <0·0001 | |
| P-value vs. 10μg GBP510 with AS03 | | <0·0001 | 0·0588 | | |
| P-value vs. 25μg GBP510 with AS03 | | | | <0·0001 | |
| GMFR±standard deviation | 160·95±2·15 | 14·19±2·58 | 210·65±1·85 | 7·06±2·86 | 0·85±1·67 |
| 95% CI (lower, upper) | (135·85, 190·70) | (7·21, 27·92) | (184·50, 240·50) | (5·07, 9·83) | (0·74, 0·98) |
| P-value vs. Placebo | <0·0001 | <0·0001 | <0·0001 | <0·0001 | |
| P-value vs. 10μg GBP510 with AS03 | | <0·0001 | 0·0140 | | |
| P-value vs. 25μg GBP510 with AS03 | | | | <0·0001 | |
| SCR, n (%) | 81 (100·00) | 10 (100·00) | 85 (100·00) | 27 (65·85) | 0 (0·00) |
| 95% CI (lower, upper) | (95·55, 100·00) | (69·15, 100·00) | (95·75, 100·00) | (49·41, 79·92) | (0·00, 6·72) |
| P-value vs. Placebo | <0·0001 | <0·0001 | <0·0001 | <0·0001 | |
| P-value vs. 10μg GBP510 with AS03 | | NA | NA | | |
| P-value vs. 25μg GBP510 with AS03 | | | | <0·0001 | |
| Visit 7 (4 weeks post-2nd vaccination) | | | | | |
| n | 81 | 10 | 85 | 41 | 53 |
| GMC±standard deviation (BAU/mL) | 1,628·59±2·02 | 132·64±2·76 | 2,073·65±1·61 | 119·45±2·71 | 12·51±1·71 |
| 95% CI (lower, upper) | (1,393·34, 1,903·56) | (64·23, 273·91) | (1,870·12, 2,299·33) | (87·24, 163·56) | (10·78, 14·51) |
| P-value vs. Placebo | <0·0001 | <0·0001 | <0·0001 | <0·0001 | |
| P-value vs. 10μg GBP510 with AS03 | | <0·0001 | 0·0112 | | |
| P-value vs. 25μg GBP510 with AS03 | | | | <0·0001 | |
| GMFR±standard deviation | 110·66±2·48 | 12·12±2·37 | 157·29±1·83 | 7·35±2·85 | 1·03±1·96 |
| 95% CI (lower, upper) | (90·50, 135·32) | (6·54, 22·47) | (138·08, 179·18) | (5·29, 10·23) | (0·85, 1·23) |
| P-value vs. Placebo | <0·0001 | <0·0001 | <0·0001 | <0·0001 | |
| P-value vs. 10μg GBP510 with AS03 | | <0·0001 | 0·0041 | | |
| P-value vs. 25μg GBP510 with AS03 | | | | <0·0001 | |

| | 10µg GBP510 with AS03 (N=81) | 10µg GBP510 (N=10) | 25µg GBP510 with AS03 (N=85) | 25µg GBP510 (N=41) | Placebo (N=53) |
|---|---|---|---|---|---|
| SCR, n (%) | 80 (98·77) | 10 (100·00) | 85 (100·00) | 26 (63·41) | 1 (1·89) |
| 95% CI (lower, upper) | (93·31, 99·97) | (69·15, 100·00) | (95·75, 100·00) | (46·94, 77·88) | (0·05, 10·07) |
| P-value vs. Placebo | <0·0001 | <0·0001 | <0·0001 | <0·0001 | |
| P-value vs. 10µg GBP510 with AS03 | | 1·0000 | 0·4880 | | |
| P-value vs. 25µg GBP510 with AS03 | | | | <0·0001 | |

GMC (Geometric Mean Concentration); GMFR, geometric mean fold rise; SCR, seroconversion rate; CI, confidence interval; BAU, binding antibody unit; NA, not applicable
GMFR (Geomtetric Mean Fold Rise) = GMC (each visit) / GMC (Baseline)
SCR (Seroconversion rate): Percentage of participants with ≥ 4-fold rise from baseline
CI (Confidence Interval)
%: n / (No· of subject who information was collected at each visit by group) * 100
The 95% CI for GMC/GMFR is calculated based on the t-distribution of the log-transformed values for geometric means or geometric mean fold rises, then back transformed to the original scale for presentation.
The 95% CI for SCR is calculated by Clopper-Pearson Methods.
P-value vs Placebo: P-value for difference of Treatment groups versus Placebo (continuous values are two sample t-test and categorical values are chi-square test or fisher's exact test.)
P-value vs GBP510 10ug with AS03: P-value for difference of each group versus GBP510 10ug with AS03 (continuous values are two sample t-test and categorical values are chi-square test or fisher's exact test.)
P-value vs GBP510 25ug with AS03: P-value for difference of each group versus GBP510 25ug with AS03 (continuous values are two sample t-test and categorical values are chi-square test or fisher's exact test.)
n = number of participants

**Table S12. Geometric mean titers and seroconversion rates of neutralizing antibody to the SARS-CoV-2 in adult aged 19-64 years: analysis by pseudovirus-based neutralization assay (per-protocol set)**

| | 10µg GBP510 with AS03 (N = 81) | 10µg GBP510 (N = 10) | 25µg GBP510 with AS03 (N = 85) | 25µg GBP510 (N = 41) | Placebo (N = 53) |
|---|---|---|---|---|---|
| Geometric mean titer and seroconversion rate | | | | | |
| Baseline | | | | | |
| n | 81 | 10 | 85 | 41 | 53 |
| GMT ± standard deviation (IU/mL) | 18·22±1·30 | 21·01±1·58 | 19·68±1·45 | 18·47±1·38 | 19·41±1·54 |
| 95% CI (lower, upper) | (17·19, 19·30) | (15·16, 29·12) | (18·16, 21·32) | (16·70, 20·43) | (17·24, 21·86) |
| P-value vs. placebo | 0·3394 | 0·5992 | 0·8426 | 0·5397 | |
| P-value vs. GBP510 10µg with AS03 | | 0·3562 | 0·1229 | | |
| P-value vs. GBP510 25µg with AS03 | | | | 0·3516 | |
| Visit 4 (4 weeks post-1st vaccination) | | | | | |
| N | 81 | 10 | 85 | 41 | 53 |
| GMT± standard deviation (IU/mL) | 38·87±2·69 | 32·79±2·45 | 43·19±2·79 | 21·03±1·64 | 19·98±1·61 |
| 95% CI (lower, upper) | (31·22, 48·38) | (17·28, 62·23) | (34·62, 53·87) | (18·01, 24·57) | (17·53, 22·78) |
| P-value vs. placebo | <0·0001 | 0·1191 | <0·0001 | 0·6107 | |
| P-value vs. GBP510 10µg with AS03 | | 0·6067 | 0·5017 | | |
| P-value vs. GBP510 25µg with AS03 | | | | <0·0001 | |
| GMFR ± standard deviation | 2·13±2·75 | 1·56±2·42 | 2·19±2·95 | 1·14±1·65 | 1·03±1·96 |
| 95% CI (lower, upper) | (1·71, 2·67) | (0·83, 2·94) | (1·74, 2·77) | (0·97, 1·33) | (0·85, 1·24) |
| P-value vs. placebo | <0·0001 | 0·0939 | <0·0001 | 0·4262 | |
| P-value vs. GBP510 10µg with AS03 | | 0·3527 | 0·8626 | | |
| P-value vs. GBP510 25µg with AS03 | | | | <0·0001 | |
| SCR, n (%) | 23 (28·40) | 1 (10·00) | 25 (29·41) | 2 (4·88) | 4 (7·55) |
| 95% CI (lower, upper) | (18·93, 39·50) | (0·25, 44·50) | (20·02, 40·29) | (0·60, 16·53) | (2·09, 18·21) |
| P-value vs. placebo | 0·0033 | 1·0000 | 0·0022 | 0·6933 | |
| P-value vs. GBP510 10µg with AS03 | | 0·2810 | 0·8852 | | |
| P-value vs. GBP510 25µg with AS03 | | | | 0·0017 | |
| Visit 6 (2 weeks post-2nd vaccination) | | | | | |
| N | 81 | 10 | 85 | 41 | 53 |
| GMT ± standard deviation (IU/mL) | 1,536·69±2·43 | 83·47±4·47 | 1,622·50±2·42 | 62·04±3·22 | 19·46±1·50 |
| 95% CI (lower, upper) | (1,262·76, 1,870·03) | (28·62, 243·48) | (1,340·93, 1,963·20) | (42·88, 89·77) | (17·41, 21·76) |

| | **10µg GBP510 with AS03 (N = 81)** | **10µg GBP510 (N = 10)** | **25µg GBP510 with AS03 (N = 85)** | **25µg GBP510 (N = 41)** | **Placebo (N = 53)** |
|---|---|---|---|---|---|
| P-value vs. placebo | <0·0001 | 0·0132 | <0·0001 | <0·0001 | |
| P-value vs. GBP510 10µg with AS03 | | 0·0001 | 0·6933 | | |
| P-value vs. GBP510 25µg with AS03 | | | | <0·0001 | |
| GMFR ± Standard deviation | 84·36±2·54 | 3·97±4·09 | 82·46±2·61 | 3·36±3·35 | 1·00±1·68 |
| 95% CI (lower, upper) | (68·64, 103·68) | (1·45, 10·88) | (67·05, 101·41) | (2·29, 4·92) | (0·87, 1·16) |
| P-value vs. placebo | <0·0001 | 0·0129 | <0·0001 | <0·0001 | |
| P-value vs. GBP510 10µg with AS03 | | <0·0001 | 0·8767 | | |
| P-value vs. GBP510 25µg with AS03 | | | | <0·0001 | |
| SCR, n (%) | 81 (100·00) | 5 (50·00) | 85 (100·00) | 22 (53·66) | 1 (1·89) |
| 95% CI (lower, upper) | (95·55, 100·00) | (18·71, 81·29) | (95·75, 100·00) | (37·42, 69·34) | (0·05, 10·07) |
| P-value vs. placebo | <0·0001 | 0·0002 | <0·0001 | <0·0001 | |
| P-value vs. GBP510 10µg with AS03 | | <0·0001 | NA | | |
| P-value vs. GBP510 25µg with AS03 | | | | <0·0001 | |
| Visit 7 (4 weeks post-2nd vaccination) | | | | | |
| N | 81 | 10 | 85 | 41 | 53 |
| GMT ± standard deviation (IU/mL) | 916·92±2·64 | 68·52±3·85 | 1,222·02±2·42 | 52·99±3·36 | 19·52±1·51 |
| 95% CI (lower, upper) | (740·07, 1,136·03) | (26·12, 179·79) | (1,009·72, 1,478·94) | (36·14, 77·69) | (17·42, 21·88) |
| P-value vs. placebo | <0·0001 | 0·0165 | <0·0001 | <0·0001 | |
| P-value vs. GBP510 10µg with AS03 | | <0·0001 | 0·0476 | | |
| P-value vs. GBP510 25µg with AS03 | | | | <0·0001 | |
| GMFR ± Standard deviation | 50·34±2·71 | 3·26±4·15 | 62·10±2·77 | 2·87±3·54 | 1·01±1·77 |
| 95% CI (lower, upper) | (40·39, 62·73) | (1·18, 9·02) | (49·86, 77·36) | (1·93, 4·27) | (0·86, 1·18) |
| P-value vs. placebo | <0·0001 | 0·0285 | <0·0001 | <0·0001 | |
| P-value vs. GBP510 10µg with AS03 | | <0·0001 | 0·1811 | | |
| P-value vs· GBP510 25µg with AS03 | | | | <0·0001 | |
| SCR, n (%) | 79 (97·53) | 5 (50·00) | 85 (100·00) | 18 (43·90) | 2 (3·77) |
| 95% CI (lower, upper) | (91·36, 99·70) | (18·71, 81·29) | (95·75, 100·00) | (28·47, 60·25) | (0·46, 12·98) |
| P-value vs. placebo | <0·0001 | 0·0006 | <0·0001 | <0·0001 | |
| P-value vs. GBP510 10µg with AS03 | | 0·0001 | 0·2366 | | |
| P-value vs. GBP510 25µg with AS03 | | | | <0·0001 | |

GMT, geometric mean titer; GMFR, geometric mean fold rise; SCR, seroconversion rate; CI, confidence interval

GMFR = GMT (each visit) / GMT (Baseline)

SCR: Percentage of participants with ≥ 4-fold rise from baseline

%: n / (No· of participant who information was collected at each visit by group) * 100

The 95% CI for GMT/GMFR is calculated based on the t-distribution of the log-transformed values for geometric means or geometric mean fold rises, then back transformed to the original scale for presentation.

The 95% CI for SCR is calculated by Clopper-Pearson Methods.

P-value versus placebo: p-value for difference of Treatment groups versus Placebo (continuous values are two sample t-test and categorical values are chi-square test or fisher's exact test)

P-value versus GBP510 10μg with AS03: p-value for difference of each group versus GBP510 10μg with AS03 (continuous values are two sample t-test and categorical values are chi-square test or fisher's exact test)

P-value versus GBP510 25μg with AS03: p-value for difference of each group versus GBP510 25μg with AS03 (continuous values are two sample t-test and categorical values are chi-square test or fisher's exact test)

**Table S13. Geometric mean concentration and seroconversion rate of IgG antibody to the SARS-CoV-2-RBD in adults aged ≥65 years: analysis by enzyme-linked immunosorbent assay (per-protocol set)**

| | 10µg GBP510 with AS03 (N=12) | 10µg GBP510 (N=0) | 25µg GBP510 with AS03 (N=11) | 25µg GBP510 (N=4) | Placebo (N=5) |
|---|---|---|---|---|---|
| Geometric mean titer and seroconversion rate | | | | | |
| Baseline | | | | | |
| n | 12 | 0 | 11 | 4 | 5 |
| GMC±standard deviation (BAU/mL) | 17·78±1·45 | | 20·33±1·86 | 17·42±1·11 | 16·24±1·71 |
| 95% CI (lower, upper) | (14·02, 22·55) | | (13·39, 30·86) | (14·69, 20·67) | (8·34, 31·61) |
| P-value vs. Placebo | 0·6933 | | 0·4977 | 0·7875 | |
| P-value vs. 10µg GBP510 with AS03 | | | 0·5337 | | |
| P-value vs. 25µg GBP510 with AS03 | | | | 0·4449 | |
| Visit 4 (4 weeks post-1st vaccination) | | | | | |
| N | 12 | 0 | 11 | 4 | 5 |
| GMC±standard deviation (BAU/mL) | 64·57±2·74 | | 86·36±2·38 | 20·05±1·56 | 13·24±1·80 |
| 95% CI (lower, upper) | (34·03, 122·49) | | (48·22, 154·67) | (9·89, 40·65) | (6·39, 27·44) |
| P-value vs. Placebo | 0·0053 | | 0·0007 | 0·2815 | |
| P-value vs. 10µg GBP510 with AS03 | | | 0·4684 | | |
| P-value vs. 25µg GBP510 with AS03 | | | | 0·0075 | |
| GMFR±standard deviation | 3·63±2·79 | | 4·25±2·30 | 1·15±1·64 | 0·82±2·30 |
| 95% CI (lower, upper) | (1·89, 6·98) | | (2·43, 7·44) | (0·52, 2·53) | (0·29, 2·30) |
| P-value vs. Placebo | 0·0118 | | 0·0025 | 0·4918 | |
| P-value vs. 10µg GBP510 with AS03 | | | 0·6933 | | |
| P-value vs. 25µg GBP510 with AS03 | | | | 0·0122 | |
| SCR, n (%) | 5 (41·67) | 0 (-) | 4 (36·36) | 0 (0·00) | 0 (0·00) |
| 95% CI (lower, upper) | (15·17, 72·33) | | (10·93, 69·21) | (0·00, 60·24) | (0·00, 52·18) |
| P-value vs. Placebo | 0·2445 | | 0·2445 | NA | |
| P-value vs. 10µg GBP510 with AS03 | | | 1·0000 | | |
| P-value vs 25µg GBP510 with AS03 | | | | 0·5165 f | |

| | 10μg GBP510 with AS03 (N=12) | 10μg GBP510 (N=0) | 25μg GBP510 with AS03 (N=11) | 25μg GBP510 (N=4) | Placebo (N=5) |
|---|---|---|---|---|---|
| Visit 6 (2 weeks post-2nd vaccination) | | | | | |
| n | 12 | 0 | 11 | 4 | 5 |
| GMC±standard deviation (BAU/mL) | 1,173·91±2·40 | | 1,558·54±2·62 | 91·89±3·93 | 12·82±1·79 |
| 95% CI (lower, upper) | (673·04, 2,047·53) | | (815·94, 2,976·98) | (10·39, 812·45) | (6·21, 26·45) |
| P-value vs. Placebo | <0·0001 | | <0·0001 | 0·0218 | |
| P-value vs. 10μg GBP510 with AS03 | | | 0·4679 | | |
| P-value vs. 25μg GBP510 with AS03 | | | | 0·0006 | |
| GMFR±standard deviation | 66·03±2·69 | | 76·67±2·70 | 5·27±4·24 | 0·79±1·63 |
| 95% CI (lower, upper) | (35·20, 123·86) | | (39·34, 149·41) | (0·53, 52·50) | (0·43, 1·45) |
| P-value vs. Placebo | <0·0001 | | <0·0001 | 0·0269 | |
| P-value vs. 10μg GBP510 with AS03 | | | 0·7217 | | |
| P-value vs. 25μg GBP510 with AS03 | | | | 0·0012 | |
| SCR, n (%) | 12 (100·00) | 0 (-) | 11 (100·00) | 2 (50·00) | 0 (0·00) |
| 95% CI (lower, upper) | (73·54, 100·00) | | (71·51, 100·00) | (6·76, 93·24) | (0·00, 52·18) |
| P-value vs. Placebo | 0·0002 | | 0·0002 | 0·1667 | |
| P-value vs. 10μg GBP510 with AS03 | | | NA | | |
| P-value vs. 25μg GBP510 with AS03 | | | | 0·0571 | |
| Visit 7 (4 weeks post-2nd vaccination) | | | | | |
| n | 12 | 0 | 11 | 4 | 5 |
| GMC±standard deviation (BAU/mL) | 943·74±2·19 | | 1,177·45±2·67 | 77·68±3·39 | 12·70±1·74 |
| 95% CI (lower, upper) | (573·91, 1,551·88) | | (608·78, 2,277·31) | (11·13, 542·43) | (6·40, 25·19) |
| P-value vs. Placebo | <0·0001 | | <0·0001 | 0·0201 | |
| P-value vs. 10μg GBP510 with AS03 | | | 0·5548 | | |
| P-value vs. 25μg GBP510 with AS03 | | | | 0·0006 | |
| GMFR±standard deviation | 53·08±2·41 | | 57·92±2·64 | 4·46±3·70 | 0·78±1·50 |
| 95% CI (lower, upper) | (30·38, 92·74) | | (30·18, 111·16) | (0·56, 35·68) | (0·47, 1·29) |
| P-value vs. Placebo | <0·0001 | | <0·0001 | 0·0717 | |
| P-value vs. 10μg GBP510 with AS03 | | | 0·8230 | | |
| P-value vs 25μg GBP510 with AS03 | | | | 0·0011 | |

| | 10µg GBP510 with AS03 (N=12) | 10µg GBP510 (N=0) | 25µg GBP510 with AS03 (N=11) | 25µg GBP510 (N=4) | Placebo (N=5) |
|---|---|---|---|---|---|
| SCR, n (%) | 12 (100·00) | 0 (-) | 11 (100·00) | 2 (50·00) | 0 (0·00) |
| 95% CI (lower, upper) | (73·54, 100·00) | | (71·51, 100·00) | (6·76, 93·24) | (0·00, 52·18) |
| P-value vs. Placebo | 0·0002 | | 0·0002 | 0·1667 | |
| P-value vs. 10µg GBP510 with AS03 | | | NA | | |
| P-value vs. 25µg GBP510 with AS03 | | | | 0·0571 | |

GMC (Geometric Mean Concentration); GMFR, geometric mean fold rise; SCR, seroconversion rate; CI, confidence interval; BAU, binding antibody unit; NA, not applicable
GMFR (Geomtetric Mean Fold Rise) = GMC (each visit) / GMC (Baseline)
SCR (Seroconversion rate): Percentage of participants with ≥ 4-fold rise from baseline
CI (Confidence Interval)
%: n / (No. of subject who information was collected at each visit by group) * 100
The 95% CI for GMC/GMFR is calculated based on the t-distribution of the log-transformed values for geometric means or geometric mean fold rises, then back transformed to the original scale for presentation.
The 95% CI for SCR is calculated by Clopper-Pearson Methods.
P-value vs Placebo: P-value for difference of Treatment groups versus Placebo (continuous values are two sample t-test and categorical values are chi-square test or fisher's exact test.)
P-value vs GBP510 10ug with AS03: P-value for difference of each group versus GBP510 10ug with AS03 (continuous values are two sample t-test and categorical values are chi-square test or fisher's exact test.)
P-value vs GBP510 25ug with AS03: P-value for difference of each group versus GBP510 25ug with AS03 (continuous values are two sample t-test and categorical values are chi-square test or fisher's exact test.)
n = number of participants

**Table S14. Geometric mean titers and seroconversion rates of neutralizing antibody to the SARS-CoV-2 in adult aged ≥65 years: analysis by pseudovirus-based neutralization assay (per-protocol set)**

| | 10μg GBP510 with AS03 (N = 12) | 10μg GBP510 (N = 0) | 25μg GBP510 with AS03 (N = 11) | 25μg GBP510 (N = 4) | Placebo (N = 5) |
|---|---|---|---|---|---|
| Geometric mean titer and seroconversion rate | | | | | |
| Baseline | | | | | |
| n | 12 | 0 | 11 | 4 | 5 |
| GMT ± standard deviation (IU/mL) | 17·00±1·00 | | 18·11±1·23 | 26·14±2·36 | 17·00±1·00 |
| 95% CI (lower, upper) | (17·00, 17·00) | | (15·73, 20·83) | (6·65, 102·76) | (17·00, 17·00) |
| P-value vs. placebo | NA | | 0·3409 | 0·3910 t | |
| P-value vs. GBP510 10μg with AS03 | | | 0·3409 | | |
| P-value vs. GBP510 25μg with AS03 | | | | 0·4581 | |
| Visit 4 (4 weeks post-1st vaccination) | | | | | |
| n | 12 | 0 | 11 | 4 | 5 |
| GMT± standard deviation (IU/mL) | 48·31±3·04 | | 27·12±2·47 | 23·30±1·88 | 17·00±1·00 |
| 95% CI (lower, upper) | (23·82, 97·97) | | (14·76, 49·81) | (8·54, 63·55) | (17·00, 17·00) |
| P-value vs. placebo | 0·0077 | | 0·1178 | 0·3910 | |
| P-value vs. GBP510 10μg with AS03 | | | 0·1891 | | |
| P-value vs. GBP510 25μg with AS03 | | | | 0·7646 | |
| GMFR ± standard deviation | 2·84±3·04 | | 1·50±2·62 | 0·89±1·26 | 1·00±1·00 |
| 95% CI (lower, upper) | (1·40, 5·76) | | (0·78, 2·86) | (0·62, 1·28) | (1·00, 1·00) |
| P-value vs. placebo | 0·0077 | | 0·1944 | 0·3910 t | |
| P-value vs. GBP510 10μg with AS03 | | | 0·1566 | | |
| P-value vs. GBP510 25μg with AS03 | | | | 0·1218 t | |
| SCR, n (%) | 5 (41·67) | 0 (-) | 2 (18·18) | 0 (0·00) | 0 (0·00) |
| 95% CI (lower, upper) | (15·17, 72·33) | | (2·28, 51·78) | (0·00, 60·24) | (0·00, 52·18) |
| P-value vs. placebo | 0·2445 | | 1·0000 | NA | |
| P-value vs. GBP510 10μg with AS03 | | | 0·3707 | | |
| P-value vs. GBP510 25μg with AS03 | | | | 1·0000 | |
| Visit 6 (2 weeks post-2nd vaccination) | | | | | |
| n | 12 | 0 | 11 | 4 | 5 |
| GMT ± standard deviation (IU/mL) | 627·68±2·91 | | 543·68±3·55 | 85·71±5·08 | 17·00±1·00 |
| 95% CI (lower, upper) | (318·55, 1,236·78) | | (232·19, 1,273·06) | (6·45, 1,138·93) | (17·00, 17·00) |

| | 10µg GBP510 with AS03 (N = 12) | 10µg GBP510 (N = 0) | 25µg GBP510 with AS03 (N = 11) | 25µg GBP510 (N = 4) | Placebo (N = 5) |
|---|---|---|---|---|---|
| P-value vs. placebo | <0·0001 | | <0·0001 | 0·1406 | |
| P-value vs. GBP510 10µg with AS03 | | | 0·7709 | | |
| P-value vs· GBP510 25µg with AS03 | | | | 0·0365 | |
| GMFR ± Standard deviation | 36·92±2·91 | | 30·03±3·93 | 3·28±5·56 | 1·00±1·00 |
| 95% CI (lower, upper) | (18·74, 72·75) | | (11·97, 75·33) | (0·21, 50·23) | (1·00, 1·00) |
| P-value vs. placebo | <0·0001 | | <0·0001 t | 0·2601 | |
| P-value vs. GBP510 10µg with AS03 | | | 0·6891 t | | |
| P-value vs. GBP510 25µg with AS03 | | | | 0·0218 | |
| SCR, n (%) | 12 (100·00) | 0 (-) | 10 (90·91) | 1 (25·00) | 0 (0·00) |
| 95% CI (lower, upper) | (73·54, 100·00) | | (58·72, 99·77) | (0·63, 80·59) | (0·00, 52·18) |
| P-value vs. placebo | 0·0002 | | 0·0014 | 0·4444 | |
| P-value vs. GBP510 10µg with AS03 | | | 0·4783 | | |
| P-value vs. GBP510 25µg with AS03 | | | | 0·0330 | |
| Visit 7 (4 weeks post-2nd vaccination) | | | | | |
| n | 12 | 0 | 11 | 4 | 5 |
| GMT ± standard deviation (IU/mL) | 459·81±2·99 | | 517·02±3·37 | 41·70±3·88 | 17·00±1·00 |
| 95% CI (lower, upper) | (229·26, 922·22) | | (228·65, 1,169·08) | (4·82, 361·16) | (17·00, 17·00) |
| P-value vs. placebo | <0·0001 | | <0·0001 | 0·2777 | |
| P-value vs. GBP510 10µg with AS03 | | | 0·8100 | | |
| P-value vs. GBP510 25µg with AS03 | | | | 0·0043 | |
| GMFR ± Standard deviation | 27·05±2·99 | | 28·56±3·83 | 1·60±6·69 | 1·00±1·00 |
| 95% CI (lower, upper) | (13·49, 54·25) | | (11·58, 70·40) | (0·08, 32·80) | (1·00, 1·00) |
| P-value vs. placebo | <0·0001 | | <0·0001 | 0·6566 | |
| P-value vs. GBP510 10µg with AS03 | | | 0·9161 | | |
| P-value vs. GBP510 25µg with AS03 | | | | 0·0056 | |
| SCR, n (%) | 12 (100·00) | 0 (-) | 10 (90·91) | 1 (25·00) | 0 (0·00) |
| 95% CI (lower, upper) | (73·54, 100·00) | | (58·72, 99·77) | (0·63, 80·59) | (0·00, 52·18) |
| P-value vs. placebo | 0·0002 | | 0·0014 | 0·4444 | |
| P-value vs. GBP510 10µg with AS03 | | | 0·4783 | | |
| P-value vs. GBP510 25µg with AS03 | | | | 0·0330 | |

GMT, geometric mean titer; GMFR, geometric mean fold rise; SCR, seroconversion rate; CI, confidence interval
GMFR = GMT (each visit) / GMT (Baseline)
SCR: Percentage of participants with ≥ 4-fold rise from baseline
%: n / (No· of participant who information was collected at each visit by group) * 100
The 95% CI for GMT/GMFR is calculated based on the t-distribution of the log-transformed values for geometric means or geometric mean fold rises, then back transformed to the original scale for presentation.
The 95% CI for SCR is calculated by Clopper-Pearson Methods.
P-value versus placebo: p-value for difference of Treatment groups versus Placebo (continuous values are two sample t-test and categorical values are chi-square test or fisher's exact test)
P-value versus GBP510 10μg with AS03: p-value for difference of each group versus GBP510 10μg with AS03 (continuous values are two sample t-test and categorical values are chi-square test or fisher's exact test)
P-value versus GBP510 25μg with AS03: p-value for difference of each group versus GBP510 25μg with AS03 (continuous values are two sample t-test and categorical values are chi-square test or fisher's exact test)

**Table S15. Cell-mediated response for CD4+ T cells expressing cytokines using intracellular cytokine staining (per-protocol set)**

| | 10μg GBP510 with AS03 (N=93) | 10μg GBP510 (N=10) | 25μg GBP510 with AS03 (N=96) | 25μg GBP510 (N=45) | Placebo (N=58) |
|---|---|---|---|---|---|
| IFN-γ | | | | | |
| Baseline (day 0) | | | | | |
| n | 19 | 2 | 17 | 7 | 15 |
| Median | 0.00 | 0.01 | 0.01 | 0.01 | 0.00 |
| IQR (Q1, Q3) | (0.00, 0.04) | (0.00, 0.01) | (0.00, 0.02) | (0.00, 0.03) | (0.00, 0.01) |
| Visit 4 (day 28) | | | | | |
| n | 19 | 2 | 17 | 7 | 15 |
| Median | 0.01 | 0.01 | 0.03 | 0.01 | 0.01 |
| IQR (Q1, Q3) | (0.00, 0.04) | (0.00, 0.01) | (0.01, 0.06) | (0.00, 0.07) | (0.00, 0.03) |
| Visit 6 (day 42) | | | | | |
| n | 19 | 2 | 17 | 7 | 15 |
| Median | 0.06 | 0.00 | 0.06 | 0.02 | 0.02 |
| IQR (Q1, Q3) | (0.02, 0.11) | (0.00, 0.01) | (0.01, 0.12) | (0.01, 0.04) | (0.00, 0.04) |
| TNFα | | | | | |
| Baseline (day 0) | | | | | |
| n | 19 | 2 | 17 | 7 | 15 |
| Median | 0.06 | 0.06 | 0.05 | 0.03 | 0.02 |
| IQR (Q1, Q3) | (0.02, 0.20) | (0.04, 0.09) | (0.03, 0.08) | (0.00, 0.07) | (0.01, 0.06) |
| Visit 4 (day 28) | | | | | |
| n | 19 | 2 | 17 | 7 | 15 |
| Median | 0.13 | 0.03 | 0.09 | 0.13 | 0.09 |
| IQR (Q1, Q3) | (0.06, 0.30) | (0.00, 0.06) | (0.03, 0.35) | (0.06, 0.32) | (0.04, 0.26) |
| Visit 6 (day 42) | | | | | |
| n | 19 | 2 | 17 | 7 | 15 |
| Median | 0.24 | 0.18 | 0.27 | 0.25 | 0.12 |
| IQR (Q1, Q3) | (0.19, 0.59) | (0.02, 0.34) | (0.10, 0.52) | (0.04, 0.32) | (0.05, 0.49) |
| IL-2 | | | | | |
| Baseline (day 0) | | | | | |
| n | 19 | 2 | 17 | 7 | 15 |

| | 10µg GBP510 with AS03 (N=93) | 10µg GBP510 (N=10) | 25µg GBP510 with AS03 (N=96) | 25µg GBP510 (N=45) | Placebo (N=58) |
|---|---|---|---|---|---|
| Median | 0.01 | 0.01 | 0.01 | 0.01 | 0.01 |
| IQR (Q1, Q3) | (0.01, 0.03) | (0.01, 0.01) | (0.00, 0.02) | (0.00, 0.02) | (0.00, 0.02) |
| Visit 4 (day 28) | | | | | |
| n | 19 | 2 | 17 | 7 | 15 |
| Median | 0.03 | 0.02 | 0.03 | 0.02 | 0.02 |
| IQR (Q1, Q3) | (0.01, 0.04) | (0.00, 0.03) | (0.01, 0.05) | (0.00, 0.04) | (0.00, 0.04) |
| Visit 6 (day 42) | | | | | |
| n | 19 | 2 | 17 | 7 | 15 |
| Median | 0.11 | 0.02 | 0.10 | 0.04 | 0.01 |
| IQR (Q1, Q3) | (0.07, 0.13) | (0.01, 0.03) | (0.04, 0.15) | (0.01, 0.04) | (0.00, 0.06) |
| IL-4 | | | | | |
| Baseline (day 0) | | | | | |
| n | 19 | 2 | 17 | 7 | 15 |
| Median | 0.01 | 0.04 | 0.01 | 0.00 | 0.02 |
| IQR (Q1, Q3) | (0.00, 0.03) | (0.00, 0.08) | (0.00, 0.10) | (0.00, 0.04) | (0.00, 0.04) |
| Visit 4 (day 28) | | | | | |
| n | 19 | 2 | 17 | 7 | 15 |
| Median | 0.00 | 0.00 | 0.02 | 0.02 | 0.01 |
| IQR (Q1, Q3) | (0.00, 0.02) | (0.00, 0.00) | (0.01, 0.04) | (0.00, 0.10) | (0.00, 0.12) |
| Visit 6 (day 42) | | | | | |
| n | 19 | 2 | 17 | 7 | 15 |
| Median | 0.00 | 0.00 | 0.00 | 0.00 | 0.01 |
| IQR (Q1, Q3) | (0.00, 0.06) | (0.00, 0.00) | (0.00, 0.05) | (0.00, 0.04) | (0.00, 0.04) |
| IL-5 | | | | | |
| Baseline (day 0) | | | | | |
| n | 19 | 2 | 17 | 7 | 15 |
| Median | 0.00 | 0.00 | 0.00 | 0.00 | 0.00 |
| IQR (Q1, Q3) | (0.00, 0.01) | (0.00, 0.00) | (0.00, 0.00) | (0.00, 0.00) | (0.00, 0.00) |
| Visit 4 (day 28) | | | | | |
| n | 19 | 2 | 17 | 7 | 15 |
| Median | 0.00 | 0.00 | 0.00 | 0.00 | 0.00 |
| IQR (Q1, Q3) | (0.00, 0.00) | (0.00, 0.00) | (0.00, 0.01) | (0.00, 0.01) | (0.00, 0.01) |

| | 10µg GBP510 with AS03<br>(N=93) | 10µg GBP510<br>(N=10) | 25µg GBP510 with AS03<br>(N=96) | 25µg GBP510<br>(N=45) | Placebo<br>(N=58) |
|---|---|---|---|---|---|
| Visit 6 (day 42) | | | | | |
| n | 19 | 2 | 17 | 7 | 15 |
| Median | 0.00 | 0.00 | 0.00 | 0.00 | 0.00 |
| IQR (Q1, Q3) | (0.00, 0.00) | (0.00, 0.00) | (0.00, 0.01) | (0.00, 0.01) | (0.00, 0.01) |

N= Total number of participants in each vaccine group; n = number of participants in each visit; IQR=Interquartile range; Q1=Quartile 3; Q1=Quartile 3.
RBD-specific CD4+ T cells producing the indicated cytokine as a fraction of total cytokine-producing RBD-specific CD4+ T cells.

**Figure S1. Clinical trial scheme.**

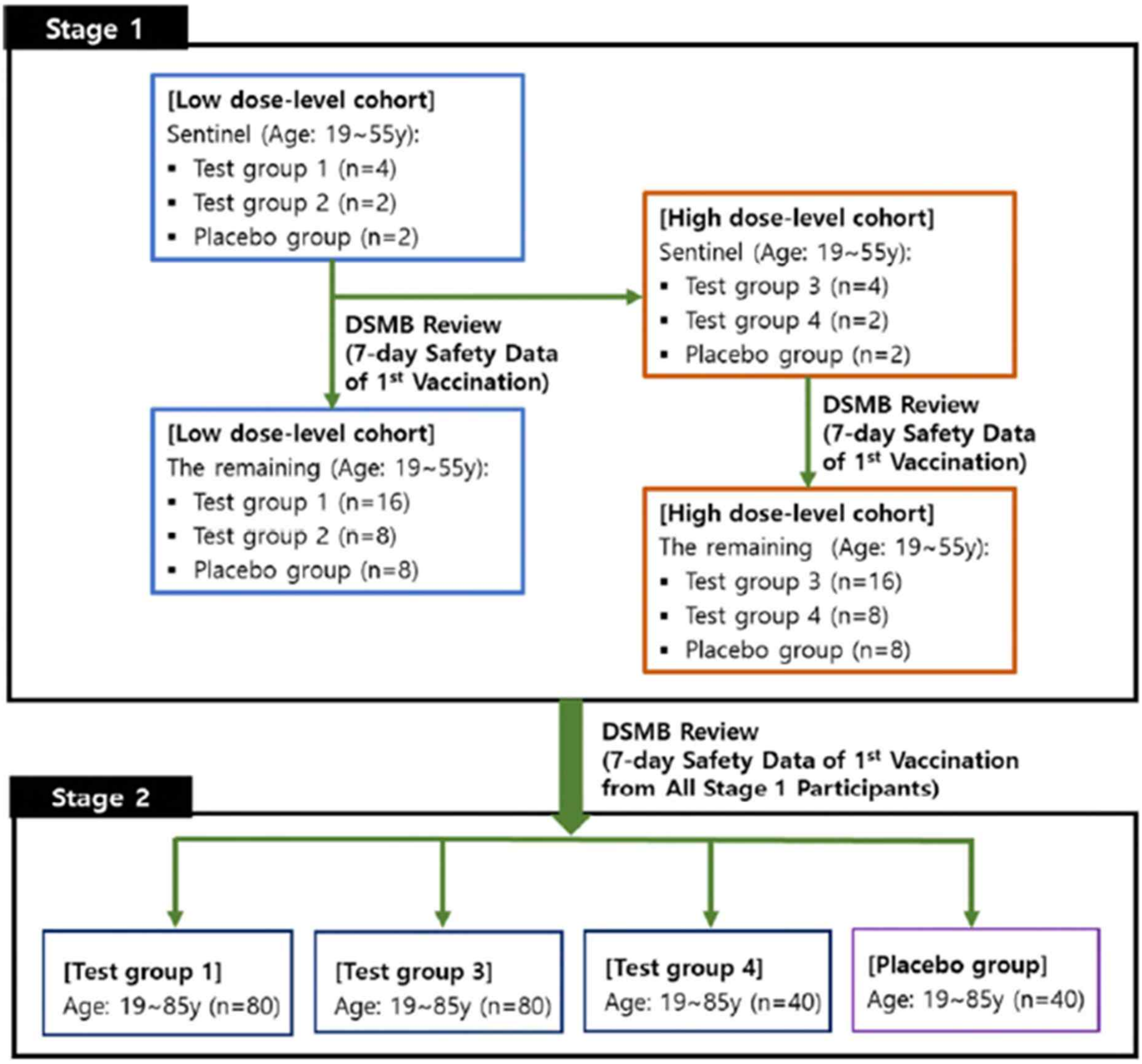

**Figure S2. Boxplot for the IFN-γ (A), TNF-α (B), IL-2 (C), IL-4 (D), IL-5 (E) of cell-mediated immune responses for CD4+ T-cells expressing cytokines using intracellular cytokine staining at baseline (day 0), visit 4 (day 28) and visit 6 (day 42).**

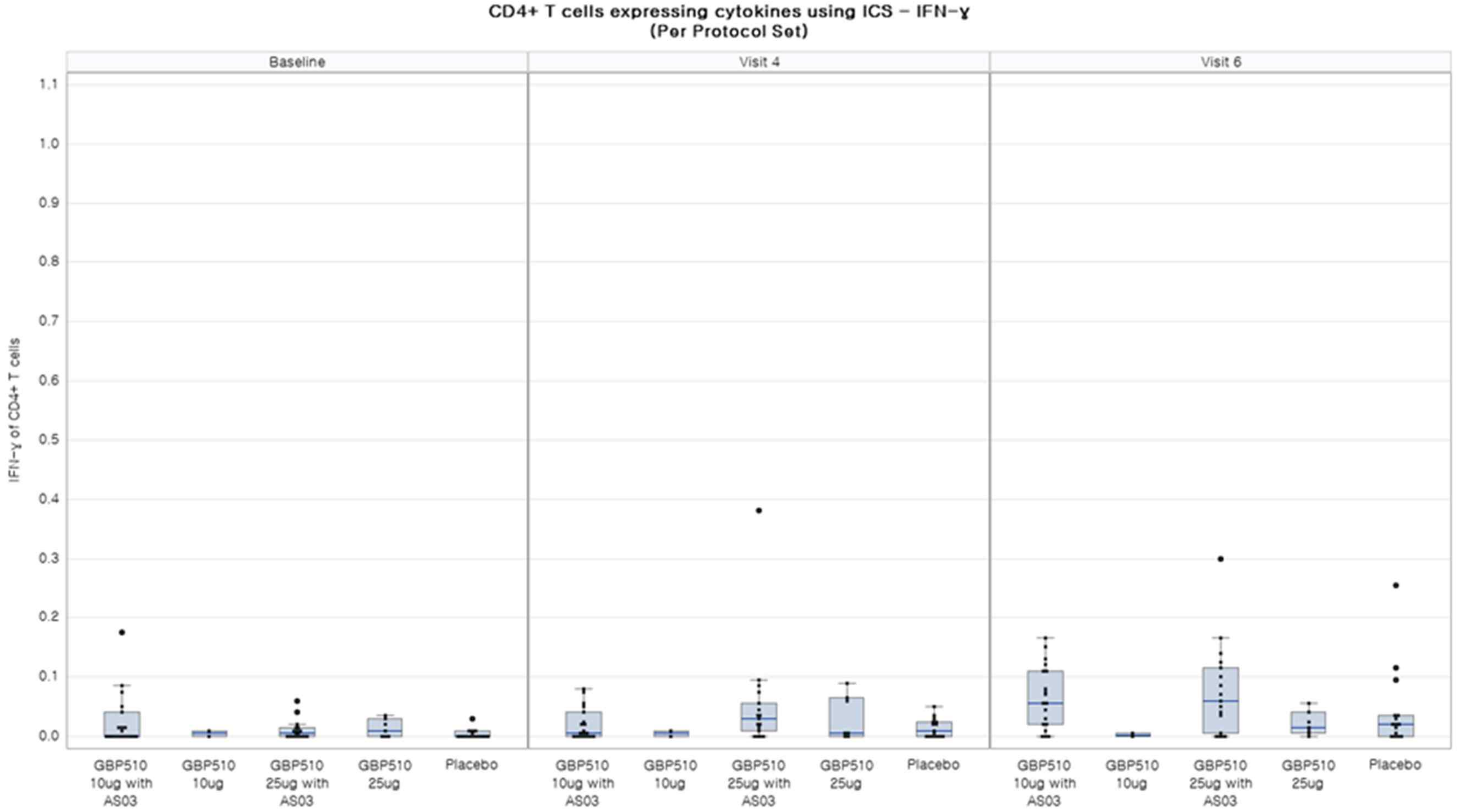

**(A)**

(B)

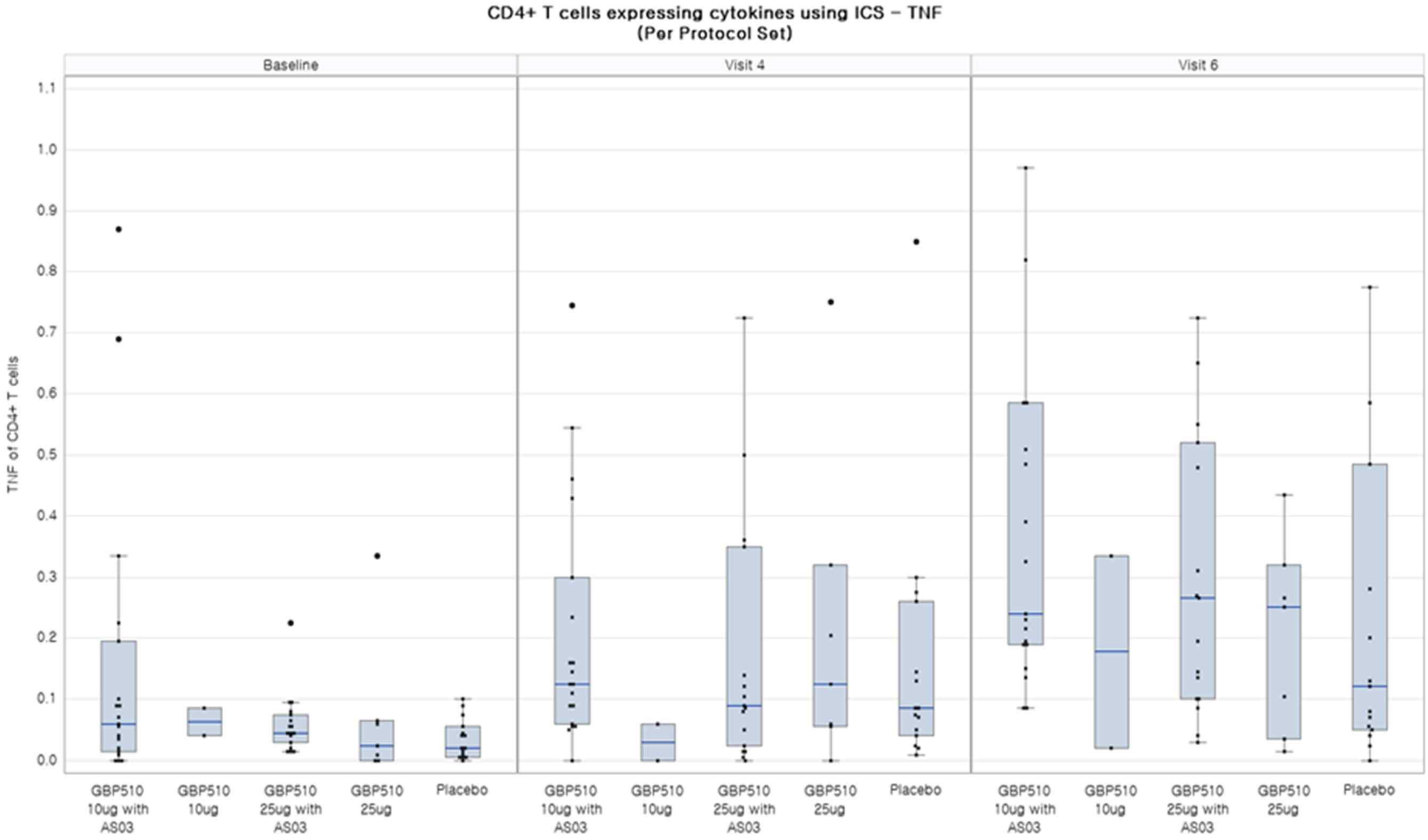
CD4+ T cells expressing cytokines using ICS – TNF
(Per Protocol Set)
Baseline
Visit 4
Visit 6
TNF of CD4+ T cells
1.1
1.0
0.9
0.8
0.7
0.6
0.5
0.4
0.3
0.2
0.1
0.0
GBP510 10ug with AS03
GBP510 10ug
GBP510 25ug with AS03
GBP510 25ug
Placebo

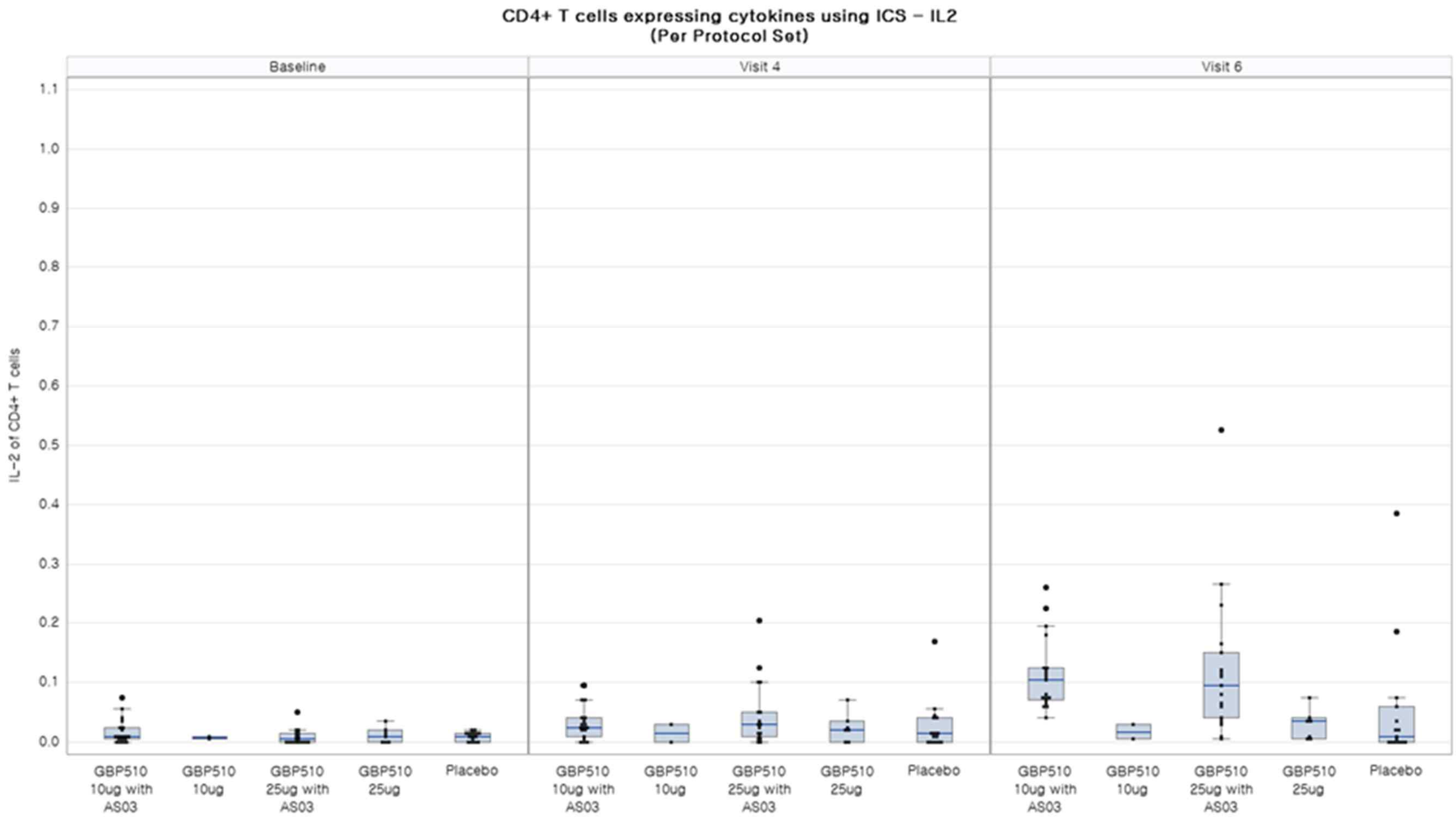
CD4+ T cells expressing cytokines using ICS - IL2
(Per Protocol Set)
Baseline
Visit 4
Visit 6
IL-2 of CD4+ T cells
1.1
1.0
0.9
0.8
0.7
0.6
0.5
0.4
0.3
0.2
0.1
0.0
GBP510 10ug with AS03
GBP510 10ug
GBP510 25ug with AS03
GBP510 25ug
Placebo

(C)

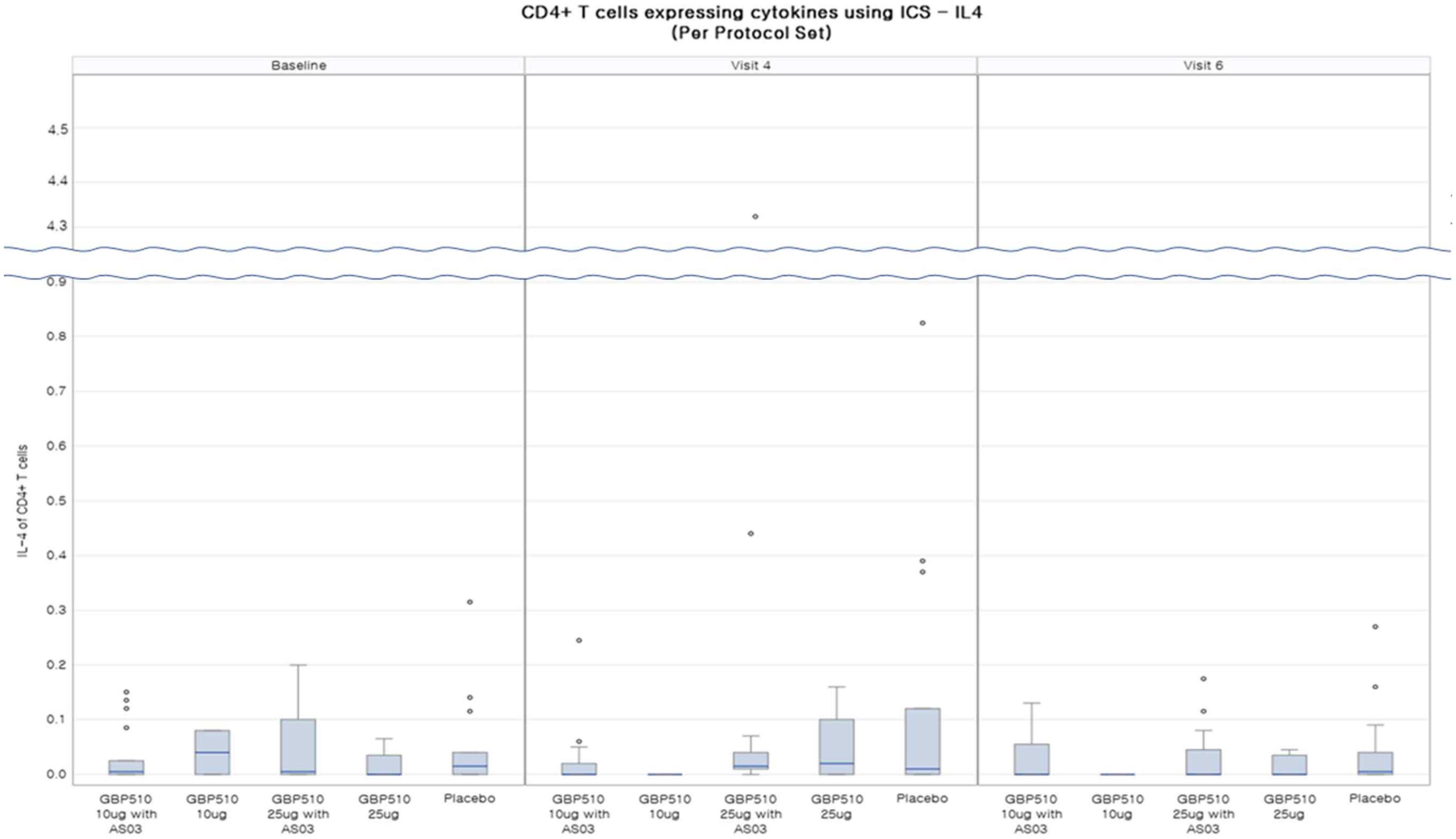
CD4+ T cells expressing cytokines using ICS – IL4
(Per Protocol Set)
Baseline
Visit 4
Visit 6
IL-4 of CD4+ T cells
4.5
4.4
4.3
0.9
0.8
0.7
0.6
0.5
0.4
0.3
0.2
0.1
0.0
GBP510 10ug with AS03
GBP510 10ug
GBP510 25ug with AS03
GBP510 25ug
Placebo

(D)

(E)

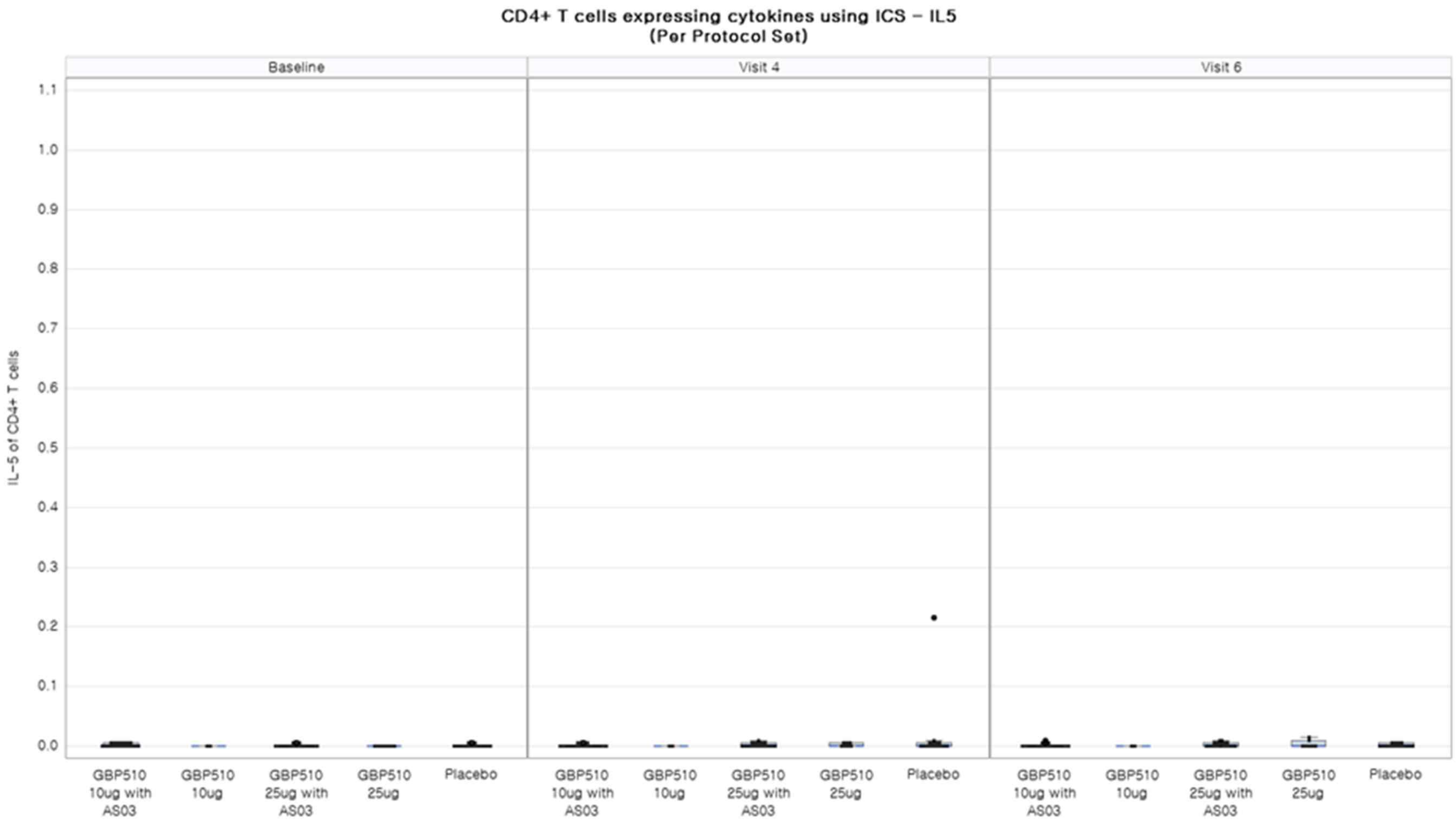
CD4+ T cells expressing cytokines using ICS – IL5
(Per Protocol Set)
Baseline
Visit 4
Visit 6
IL-5 of CD4+ T cells
1.1
1.0
0.9
0.8
0.7
0.6
0.5
0.4
0.3
0.2
0.1
0.0
GBP510 10ug with AS03
GBP510 10ug
GBP510 25ug with AS03
GBP510 25ug
Placebo

**Figure S3. Age-stratified analysis of solicited local (A) and systemic (B) adverse events within 7 days after first-dose and second-dose (proportion of participants with adverse events).**

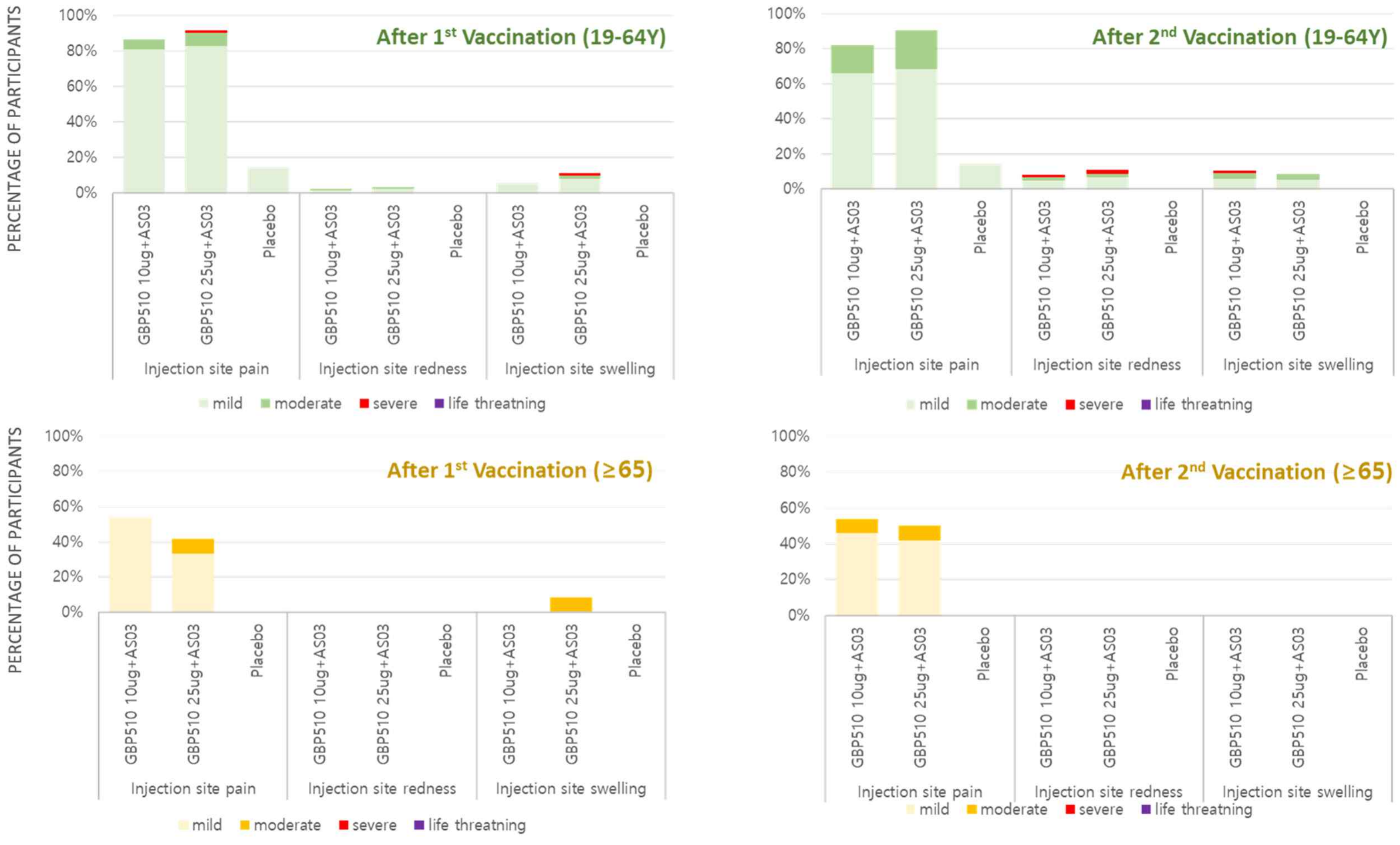

(A)

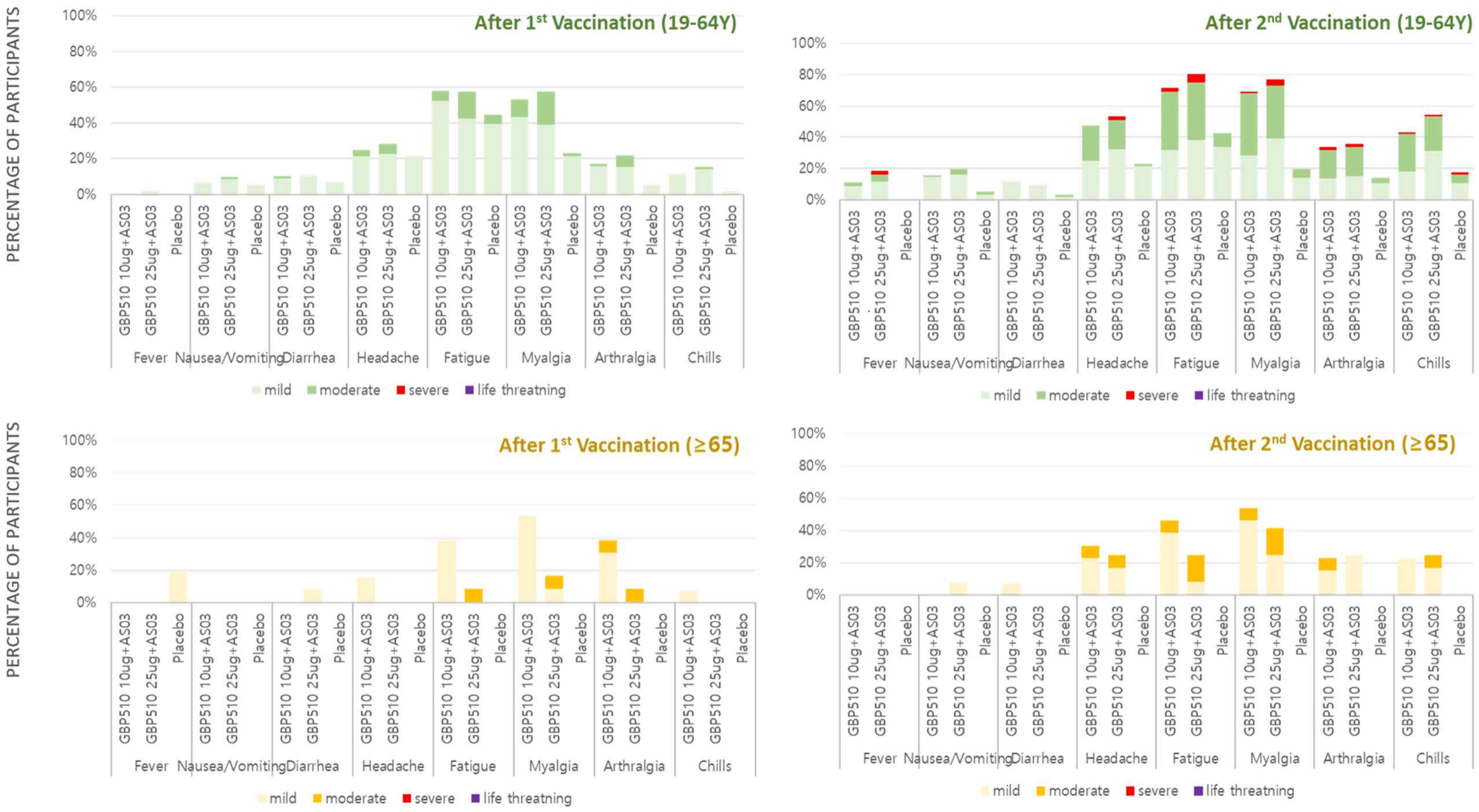
After 1st Vaccination (19-64Y)
After 2nd Vaccination (19-64Y)
After 1st Vaccination (≥65)
After 2nd Vaccination (≥65)
PERCENTAGE OF PARTICIPANTS
100%
80%
60%
40%
20%
0%
GBP510 10ug+AS03
GBP510 25ug+AS03
Placebo
Fever
Nausea/Vomiting
Diarrhea
Headache
Fatigue
Myalgia
Arthralgia
Chills
mild
moderate
severe
life threatning

(B)

**Figure S4. Cross-reactive immunogenicity against the delta variant virus (B·1·617) in 25 µg GBP510 adjuvanted with AS03 recipients, assessed by the plaque reduction neutralization test (PRNT). Data from randomly selected 11 participants aged 19–85 years (A). Data from randomly selected 11 participants aged ≥ 65 years.**

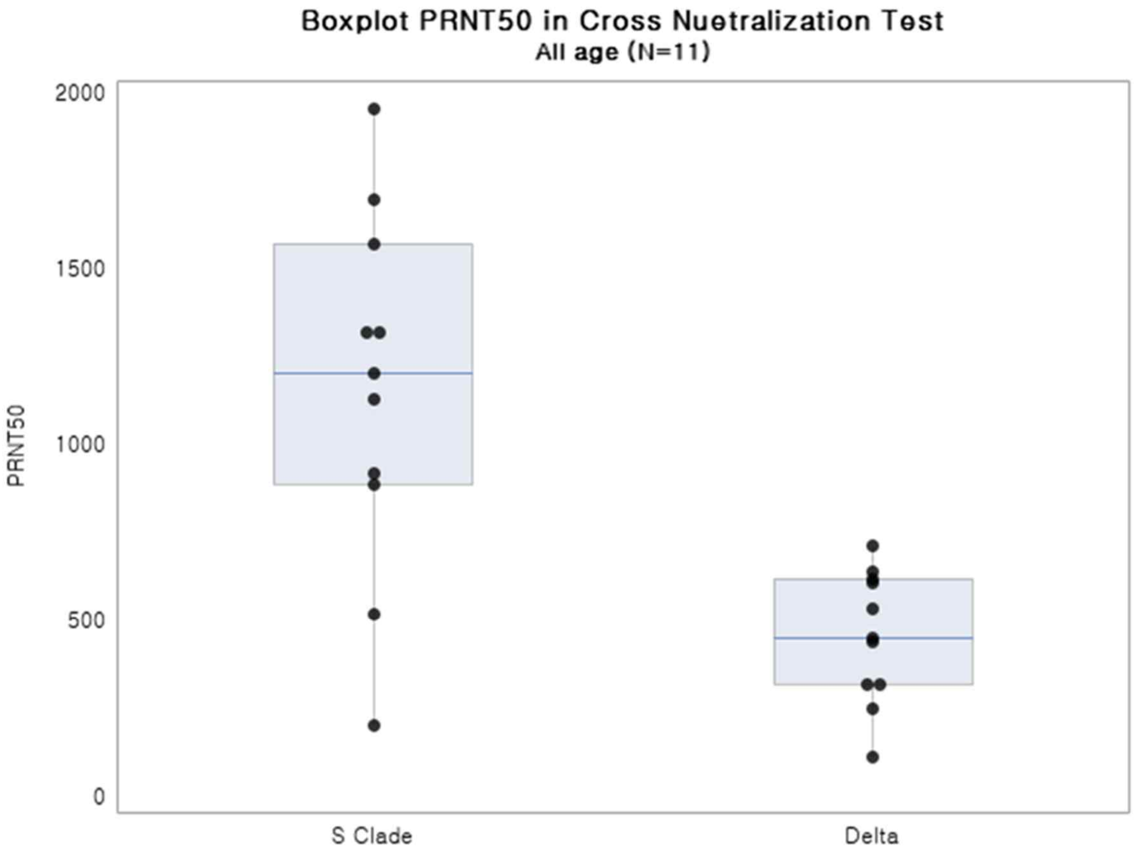

(A)

| GMT (SD) | 1009.70 (1.90) | 412.44 (1.70) |
|---|---|---|

(B)

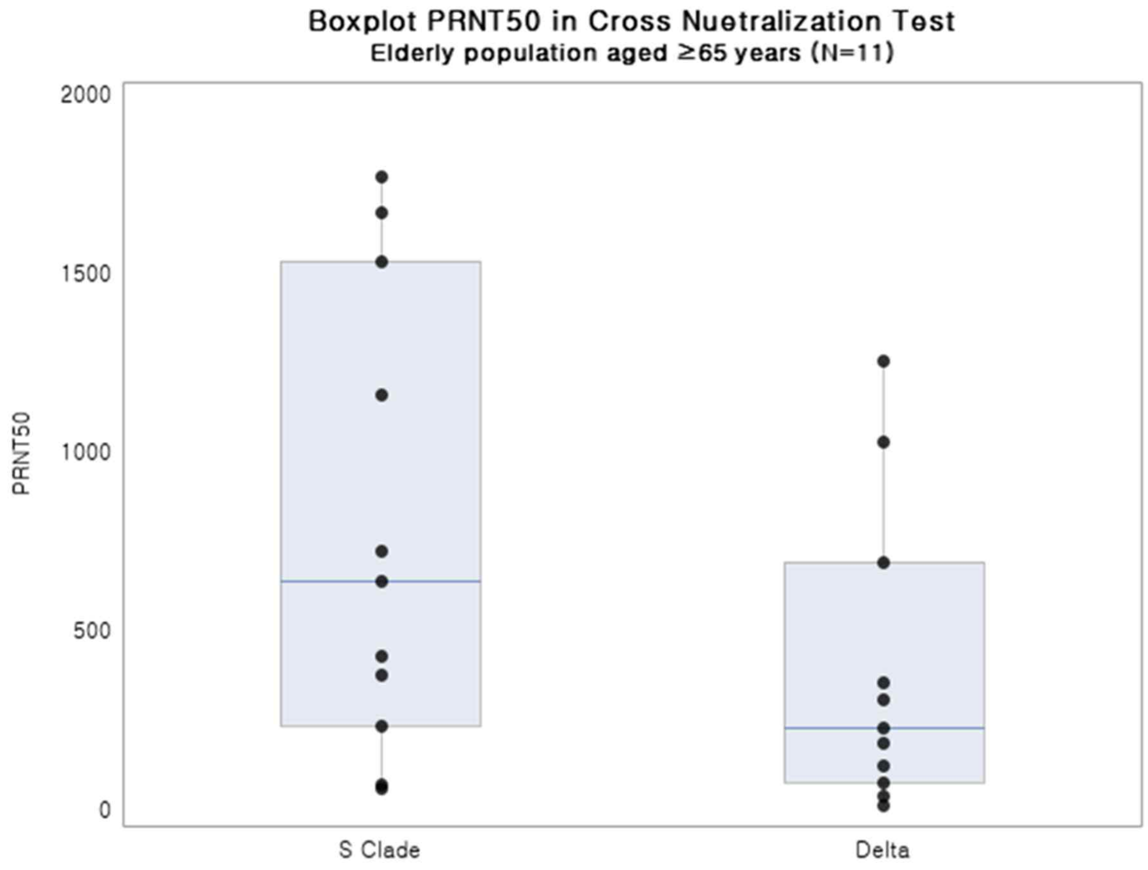

| GMT (SD) | 495.59 (3.26) | 206.59 (3.84) |
|---|---|---|
